# Adaptive combination of interventions required to reach population immunity due to stochastic community dynamics and limited vaccination

**DOI:** 10.1101/2020.12.16.20248301

**Authors:** Björn Goldenbogen, Stephan O Adler, Oliver Bodeit, Judith AH Wodke, Ximena Escalera-Fanjul, Aviv Korman, Maria Krantz, Lasse Bonn, Rafael Morán-Torres, Johanna EL Haffner, Maxim Karnetzki, Ivo Maintz, Lisa Mallis, Hannah Prawitz, Patrick S Segelitz, Martin Seeger, Rune Linding, Edda Klipp

## Abstract

Reaching population immunity against COVID-19 is proving difficult even in countries with high vaccination levels. We demonstrate that this in part is due to heterogeneity and stochasticity resulting from community-specific human-human interaction and infection networks. We address this challenge by community-specific simulation of adaptive strategies. Analyzing the predicted effect of vaccination *into* an ongoing COVID-19 outbreak, we find that adaptive combinations of targeted vaccination and non-pharmaceutical interventions (NPIs) are required to reach population immunity. Importantly, the threshold for population immunity is not a unique number but strategy and community dependent. Furthermore, the dynamics of COVID-19 outbreaks is highly community-specific: in some communities vaccinating highly interactive people diminishes the risk for an infection wave, while vaccinating the elderly reduces fatalities when vaccinations are low due to supply or hesitancy. Similarly, while risk groups should be vaccinated first to minimize fatalities, optimality branching is observed with increasing population immunity. Bimodality emerges as the infection network gains complexity over time, which entails that NPIs generally need to be longer and stricter. Thus, we analyze and quantify the requirement for NPIs dependent on the chosen vaccination strategy. We validate our simulation platform on real-world epidemiological data and demonstrate that it can predict pathways to population immunity for diverse communities world-wide challenged by limited vaccination.

---

In response to the COVID-19 pandemic major efforts have been carried out to provide models and data driven support for public health and government decision making^1–12^. These have predominantly focused on individual countries^13–17^ whilst others have aimed to integrate worldwide high-resolution demographic and mobility data to simulate the disease^18–20^. However, even in countries with strong public health governance there is often a discrepancy between what is required for specific, effective and fast decision making and what models can actually offer^7,21,22^. This is in part due to the fact that many models are based on population-wide assertions and not human individuals^23,24^. In contrast, virus outbreaks are non-linear, stochastic, network-based and localized. The propagation between human individuals depends on the specific geospatial and demographical context as well as contact patterns between groups of individuals^25,26^. Consequently, the spread of SARS-CoV-2 in the population is a complex system and must be analyzed accordingly.

Thus, here we explore whether essential aspects of the system’s behavior (e.g. stochasticity and bimodality) may be missed if the system is assumed to be homogeneous or modelled deterministically. We hypothesize that different types of epidemiological models may be useful at different phases of a pandemic. For example, in phases with high infection numbers a homogenous mixing model may well predict relevant effects, whilst in early or late phases of a pandemic or with new variants emerging it may be vital to capture stochasticity and structural heterogeneity within the system.

These possibilities are perhaps most important to consider in the pursuit of population immunity for COVID-19. Even in countries like Israel with relatively high vaccine uptake^27^ reaching population immunity is proving elusive; or in the case of the Seychelles, a nation among the fastest to vaccinate its population (>70% received at least one vaccine dose by May 12^th^ 2021), which has subsequently experienced a surge in infection cases^28^. These challenges are further emboldened by the relatively high attack-rate that has been demonstrated for SARS-CoV-2^29^, and the fact that many countries around the world are faced with limited vaccine supply or vaccine hesitancy.

Thus, there is an urgent need to explore a new paradigm for epidemiological governance based on community-specific spatio-temporal adaptive combinations of NPIs and targeted vaccination. To keep vulnerable citizens, heterogeneous and diverse societies safe, e.g. from new variants, while allowing economic and social activity to resume, governments may have to continuously adapt and combine NPIs and vaccination strategies in order to reach and maintain population immunity and avoid fatalities. Therefore, it is an open question whether adaptive approaches may be required in order for societies to reach and maintain population immunity in an increasingly NPI fatigued world.

Here, we present a globally applicable platform to analyze the effect of adaptive population immunity strategies in different communities. Based on precision simulation of individualized, real-world, spatio-temporal SARS-CoV-2 transmission networks the methodology can determine adaptive combinations and optimality in intervention strategies. By enabling prediction of the effects of combining vaccination strategies with NPIs in an adaptive and context specific manner we demonstrate how it is possible to reach and maintain population immunity. The platform is readily applicable to diverse communities world-wide and offers the ability to identify key aspects of COVID-19 outbreaks that epidemiological homogenous mixing models cannot detect.

## Results

### Heterogeneous model of human-human interaction networks enables precision-simulation of COVID-19 outbreaks

Human-human interaction networks (HHIN) are formed by physical proximity between individuals in time and space and depend on the typical or exceptional behavior of humans (**Fig. 1**). The spreading of respiratory diseases, such as COVID-19, can be described as a sub-network of infection HHIN (iHHIN) within a HHIN consisting of infection emitters and receivers. As a result, these networks are stochastic and evolve over time in a non-linear manner and their analysis requires models that can both capture this complexity and human behavior. We developed a detailed agent-based geospatial model^30^, where each agent represents a human individual within a real-world community (**Fig. 1, Online Methods, Supplementary Material, Supplementary Figs. 1-32, Supplementary Tables 1-26**). We complemented a classical SIR model with the clinically described stages of SARS-CoV-2 infection^31^ and COVID-19 (**Fig. 1f, Online Methods**) and incorporated georeferenced information^32,33^, demographic data^32,33^ and realistic weekly schedules (**Fig. 5**). This enables the model to reflect the current state of the pandemic and to simulate realistic scenarios within concrete human populations; the respective HHIN and iHHIN can be reconstructed from the simulations (**Fig. 1a**). We first simulated a baseline scenario, i.e. the spread of infection from one/few individuals through the population without any NPIs. Then, we simulated various NPIs such as full lockdown, closure and reopening of selected locations, non-compliance with interventions as well as the effect of different values of infectivity (where lowering mimics social distancing or mask wearing, while new strains can lead to an increase). Here, we applied the approach to communities in Germany, Israel, United Kingdom and Sweden, including the German town Gangelt, which witnessed one of the first outbreaks and has been thoroughly monitored during the pandemic^34^.

**Fig. 1:**
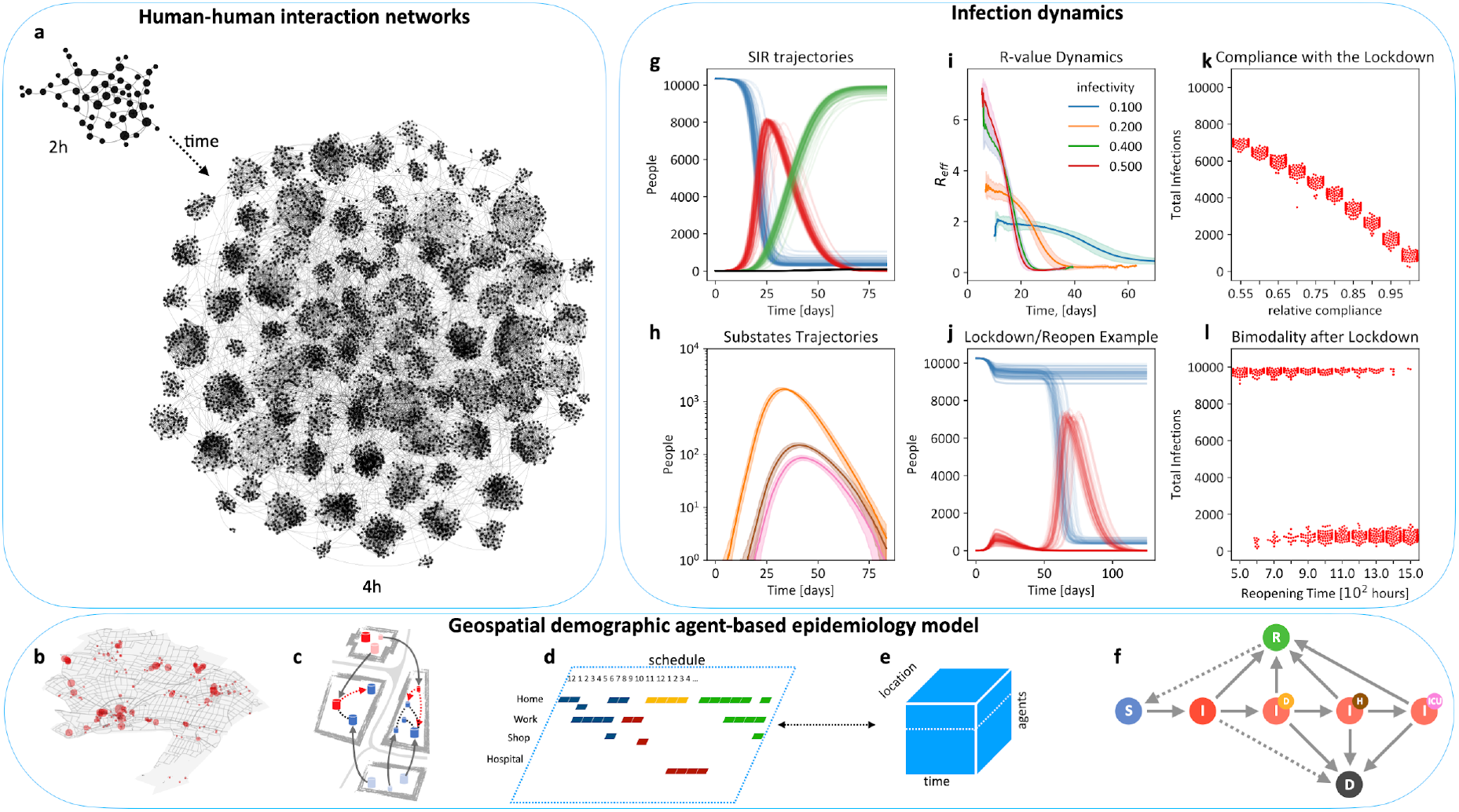
The nonlinear network effects of SARS-Cov-2 outbreaks are quantified with non-homogenous spatio-temporal models of individual human behavior. **a**, Human-human interactions create dynamic stochastic networks in space and time. **b**, The model uses data from real-world communities with annotated buildings, demographics, and statistics on daily occupations (**<u>Movie 1</u>**). **c**, Individuals move between locations to meet other individuals, enabling infection transmission. **d**, Schedules define typical behavior and where-abouts of individuals per hour. **e**, Individuals, locations, and time span a multidimensional space for stochastic simulations. **f**, An individual’s health status can be susceptible - S, infected - I, recovered - R, deceased - D, and infection sub-states a- or presymptomatic (plain I), diagnosed (I^d^), hospitalized (I^d^_H_), in intensive care (I^d^_ICU_). **g**, Simulation of uncontrolled baseline scenario for a town (here the German town Gangelt with 10.351 individuals), starting from 4 infected individuals; dynamics of states S, I, R, and D over approx. 12 weeks (100 replicates, colors as in **f**). **h**, Dynamics of I, I^d^, I^d^_H_, and I^d^_ICU_ (colors as in **f**). **i**, The R-value as emergent model property for different values of the model parameter infectivity, which may change with NPIs such as mask wearing or social distancing. **j**, Simulated performance of an NPI: lockdown (8 days after first infection) and reopening (after 5 weeks). **k**, Compliance (in % of population) with the lockdown leads to less total infection. **l**, Bimodality: whether lockdown and reopening lead to high or low infection numbers depends on the reopening time. For a range of reopening times, high/low I are obtained in a certain ratio, indicating uncertainty in the outcome. Parameter values: infectivity *k*_*I*_ = 0.3 (if not stated otherwise in **i**), interaction frequency *μ* = 2.

### Community-specificity of SARS-CoV-2 outbreak dynamics determined by heterogeneous groups of emitters and receivers

We set out to determine how interaction and infection networks could provide the means to quantify routes of SARS-CoV-2 spreading and thus serve as a decision basis for NPIs. The HHIN encompass three different classes of interactions, namely those that: (i) cannot lead to transmission of infection (e.g. between two S or between two I), (ii) can potentially result in transmission (interaction between S and I without successful transmission), and (iii) result in transmission of infection from I to S, which defines the iHHIN (**Supplementary Fig. S18**).

Thus, we simulated the German community Gangelt (G) and the Israeli community Zikhron Ya’akov (ZY) to analyze the dynamic networks and to find community-specific differences in the baseline scenario without NPIs (**Fig. 2)**. The two communities have different demographics (e.g. more children in ZY), a different geography and composition of locations (e.g. more schools in ZY, **Fig. 2a**), and while the average household size in Germany is 1,9 it is 3,2 in Israel.

**Fig. 2:**
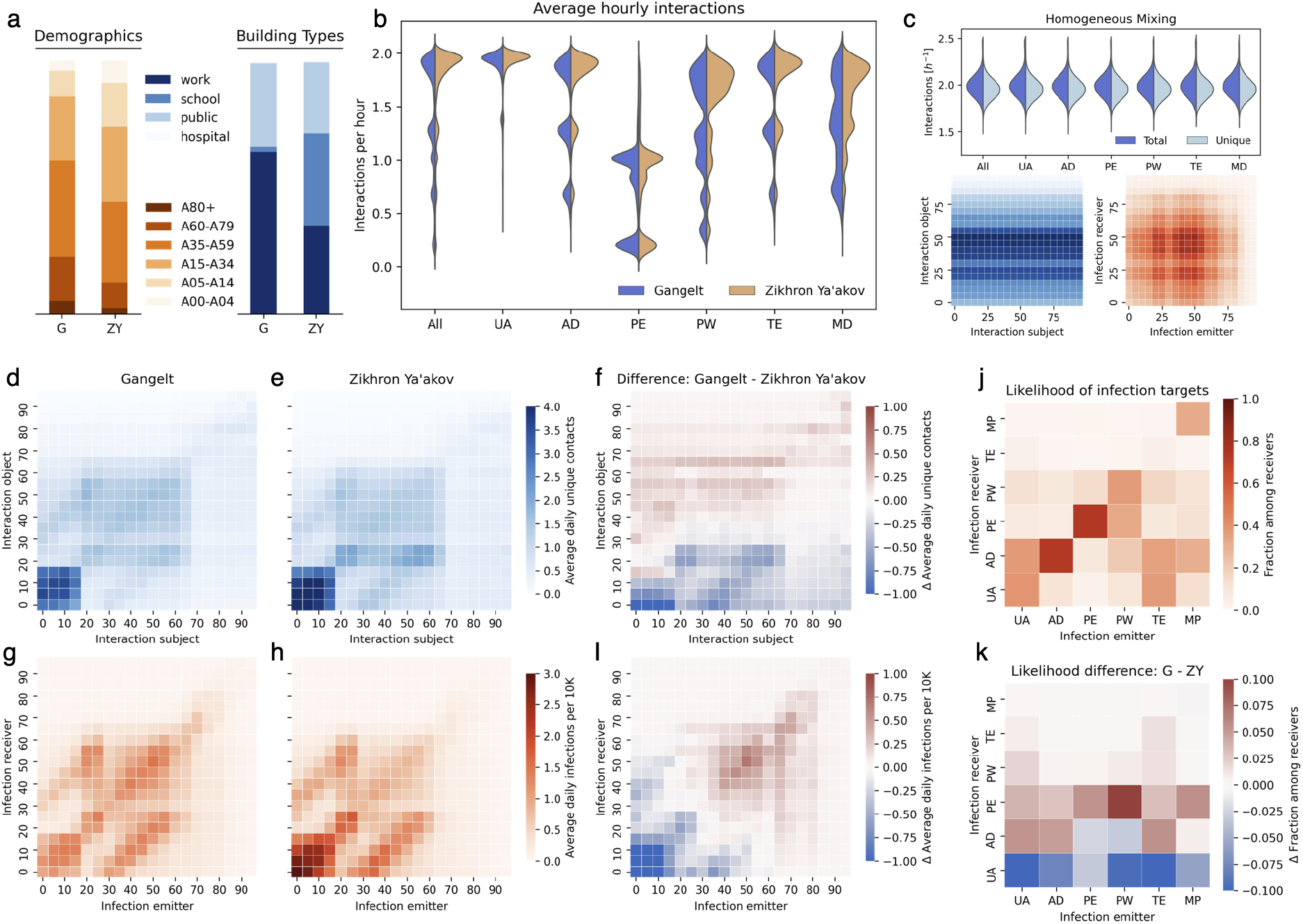
Geolocation and demographics alter the interaction networks and the routes of SARS-CoV-2 spreading, exemplified for Gangelt (DE) and Zikhron Ya’akov (IL). Simulations of Gangelt (G, N=10.296 agents) and Zikhron Ya’akov (ZY, N=17.764 agents) predict different spread of infections despite similar initial conditions and parameters, because both differ w.r.t. demographics and number/types of geolocations. **a**, Distribution of building types (excluding homes for visibility) and of age groups in G and ZY. **b**, Distribution of average hourly interactions for different population groups. Unique interactions are given in **Supplementary Figure S29. c**, Results of a homogenous mixing model (neglecting daily routines) for comparison of total and unique interactions and interaction and infection patterns between age cohorts (demographics correspond to Gangelt). **d, e, f**, Patterns of average daily interactions for **d** G and **e** ZY between age groups exhibiting superposition between intra-household interactions (diagonal patterns) and location type specific interactions (square patterns) **f**, The difference between **d** and **e** reflects the increased interactions of children in ZY **g, h, i**, Patterns of daily infections between age groups per 10.000 individuals for the baseline scenario in **g** G and **h** ZY displaying a strong contribution of intra-household infections. **i** Difference between **g** and **h** showing the shift of infections towards younger cohorts in ZY. **j, k**, Likelihoods to transmit the infection from emitters to receivers sorted by occupation for **j** G and **k** the difference between G and ZY. UA - Underaged, AD - adults (age groups 20-65 excluding PW, MP, and TE), PW - public workers, MP - medical professionals, TE - teachers, PE - pensioners. Parameter values: infectivity *k*_*I*_ = 0.3, interaction frequency *μ* = 2.

The HHIN and iHHIN are age- and occupation dependent. The distribution of interactions per occupation reveals that underaged in general have more interactions than others, even than public workers (**Fig. 2b**). Comparing G and ZY, we find more small clusters in G and a (slightly) stronger accumulation around a value of two interactions per hour in ZY (note that 2 per hour is the chosen value of the global interaction rate). **Figure 2c** provides interaction and infection patterns for a homogeneous mixing scenario as comparison to appreciate of the location- and demographics-based effects.

Analysis of age-specific interaction patterns reveals strong interactions within households, indicated by the relative strength of interactions within and between adjacent age cohorts (partnerships) and parent-child related age-cohorts (**Fig. 2d and e**, center and off-center diagonals). Underaged individuals, as well as the working population, show stronger interactions with members of the same group, as apparent from rectangular interaction patterns representing schools and workplaces. This resembles real-world interaction patterns^8,38^, but strongly differs from predictions for homogeneous mixing assumptions with same age distribution (**Fig. 2c, Supplementary material**). **Fig 2f** highlights the differences between G and ZH indicating that both working adults and children have more contacts in G. The latter is due to the fact that G has only two schools, thus a higher number of students per school than ZY.

The infection transmission in the baseline scenario reflects the interaction patterns yielding high infection transmission numbers within households and within the group of underaged, medium infection transmission between working adults and lower infection rates among pensioners, when sorted by age (**Fig. 2g,h**). This holds in general for both communities. In contrast, the infection patterns (like the interactions) in homogeneous mixing are solely determined by the prevalence of different age-cohorts and not by social structure (**Fig. 2c). Fig 2i** elucidates that infection transmission among underaged but also between underaged and their parents is clearly stronger in ZH than G, which can be explained by the higher proportion of underaged individuals in the Israeli demographic structure.

Sorting infections according to occupation uncovers that underaged most likely infect other underaged and adults, while adults predominantly emit to other adults, and pensioners mostly infect each other. However, public workers emit towards other public workers and pensioners and, hence, create an infection hub between the groups (**Fig. 2j** for Gangelt). **Fig. 2k** shows that in ZY compared to G the larger ratio of underaged makes underaged more likely as receiver of an infection, except for medical personnel as emitter due to the lower likelihood of hospital admission for the underaged.

We also find that underaged are clearly overrepresented as emitters, followed by adults, while pensioners are underrepresented. Public workers are slightly underrepresented as emitters, while teachers and medical professionals belong to the average. These interaction and infection patterns can change significantly when NPIs are applied (**Supplementary material, section 4.5**).

The stochasticity within the iHHIN can be recognized from the impact of an individual infection event, which may either not give rise to further infection events or further grow the network. We observed that 70% of infections originate from only 20% of the infected population and that 70% of infected do not spread the infection further. The iHHNI exhibits emergent patterns, which help to understand infection spread and provide a basis to efficiently interrupt infection transmission, as discussed below.

### Outcomes from interventions can be like flipping a coin

We hypothesized that the stochastic nature of infections and non-linearity that pursues may create basins of attractions or attractor states that are highly context dependent. This would mean that even with identical initial conditions the same community may with a certain likelihood end up with opposite outcomes following NPIs.

This possibility is perhaps best illustrated in the case of airliners: The NPIs on most flights are identical (negative tests, masks, distributed seating, air filtering etc). As a result, most flights neither contribute significantly to new outbreaks nor see significant transmission onboard -- but not all flights. There are several spectacular known incidents where due to the stochastic nature of who sits next to whom and even within the controlled environment of an airliner major transmission networks emerge as a result of the flight^35^. This unique phenomenon in a complex system is known as bimodality^36^ and the corresponding distributions are poorly characterized by traditional statistical metrics such as mean and variance ^37^.

Our model predicts bimodality in multiple situations: For example, a strict lockdown with reopening after a given period *T* can lead to either termination of an ongoing outbreak or - the opposite - a subsequent wave of infection within a certain window of *T* (**Fig. 1j**). Likewise, selective reopening of either schools, public or workplaces after lockdown can result in bimodal behavior of the system (**Fig. 1j** and **Supplementary Fig. S12-S15** for variation of reopening time points). Also, the dynamics after introduction of a few initial infections can either cease or lead to an outbreak (**Supplementary Fig. S17**). The bimodality, however, also implies that early lifting of NPIs does not necessarily lead to a second outbreak, which can be observed across the different simulated communities (**Supplementary Fig. 12**). Bimodality demonstrates that effective NPIs require strict execution (stringency) and careful temporal control (timing), because the likelihood of a following wave decreases with increasing length of the lockdown. Thus, even in an ideal situation with 100% covering sentinel and surveillance data available in real-time it would not be possible to predict the optimal strategy without a model that captures both stochasticity and non-linearity of the transmission network. Moreover, this challenge may scale differently across communities with, e.g. different numbers of inhabitants and social structure (**Supplementary Fig. S31**). Therefore, community and location-specificity of the epidemiological model deployed is essential for accurate prediction of adaptive and optimal strategies.

### Quantifying the effect of targeted immunization strategies

Hitherto, NPIs alone have not been sufficient to stop the COVID-19 pandemic. We hypothesize that this, at least partially, may be due to the observed bimodality and community-specificity. Therefore, we set out to determine how strategic targeting of vaccination may be required to reach population immunity.

Given that effective vaccines are now available, but not for everybody at the same time, we can use the model that has been trained for different communities and for different NPI scenarios to analyze the effect of pharmaceutical interventions. To this end, it is critical to define which specific objective applies when searching for optimal targeted immunization strategies for its communities^39^. Here, we analyze four alternatives: (i) minimize the number of fatalities, (ii) reduce the number of infections (attack rate), (iii) reduce the number of hospitalizations or individuals requiring intensive-care treatment to prevent a collapse of the health system, (iv) robustness to future outbreaks.

We simulated seven vaccination strategies (**Fig. 3, Supplementary Figure S25-S27**), where individuals were either sorted by descending *age*, by descending *interaction* frequency, according to their infection time in previous unaltered simulations (*forecasted*) or not sorted at all (*random*). Additionally, we defined a *combined* strategy, where individuals above age 60 were prioritized and the rest were chosen from the most interactive individuals. The strategies were compared with respect to the attack rate (ratio of infected individuals to all, **Fig. 3a**, the proportion of fatalities (**Fig. 3b)**, the likelihood of an emerging infection wave, which also provides a measure for the robustness of strategy against infection waves (**Fig. 3c**), and the maximum number of individuals simultaneously requiring an ICU (**Fig. 3d**) for different fractions of vaccinated individuals. Here, we discuss this for an infectivity of 0.15, corresponding to an R_0_-value of 2.7, which is in the reported range of 2.3 - 3.4 for Germany; variation of R_0_-values would modify the quantitative, but not the qualitative outcomes (**Supplementary Figure S26**).

**Fig. 3:**
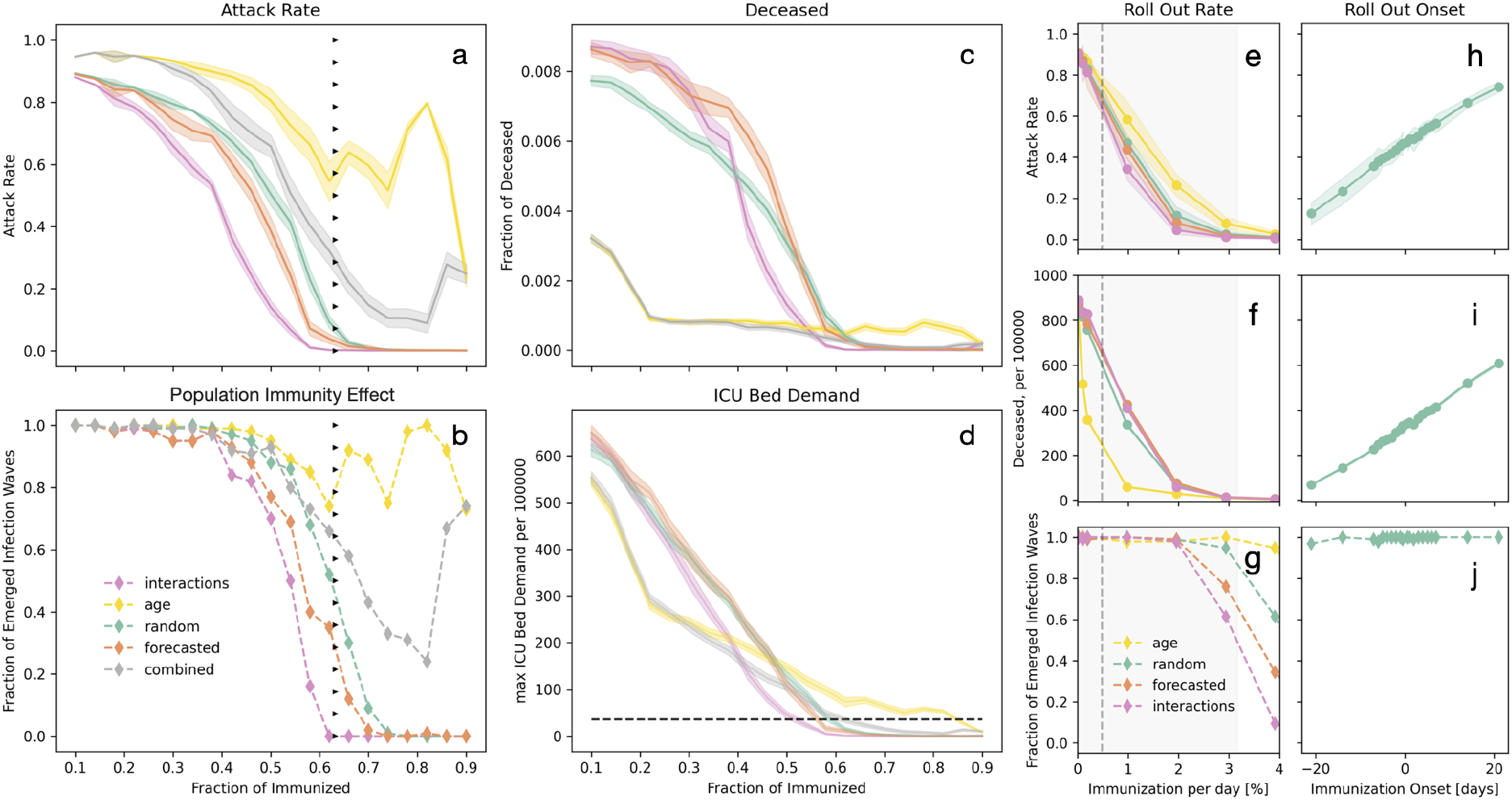
Vaccination strategies alter attack rate, population immunity threshold, death toll and burden on the health care system. Compared vaccination strategies are: *random* - individuals are selected randomly, *forecasted* - individuals are selected according to their infection order in a previous simulation, *interaction* - individuals are selected by descending interaction frequency, *age* - individuals are selected by descending age, and *combined* - individuals are first selected by descending age (>60), the rest was ordered by their interaction frequency. **a-d** Impact of a newly introduced infection on a partially immunized population (10%-90%): a fraction of the population selected by the named criteria was set to be immunized, then four new infections were introduced. Population effect: **a**, Attack rate, **b**, Fraction of simulation runs, which exhibited an infection wave (l> 80 subsequent infections) after the vaccination. For comparison, triangles indicate the population immunity threshold calculated from the estimated R_0_ of 2.71 for the chosen infectivity of 0.15, which coincides with our random strategy. **c**, Death toll: fraction of deceased relative to susceptible at start, **d**, Burden on health care system: Maximum number of individuals simultaneously requiring an ICU treatment. The black line indicates the capacity of ICU beds (assuming the German number of roughly 30.000 per 82 million inhabitants). **a, c, d**). Lines and shaded areas represent mean values and confidence intervals, respectively (CI=95%, N=100). **e-g**: Effect of roll out rate on **e** attack rate, **f** death toll, and **g** population immunity, assuming immunization starts concomitantly with introduction of infections. Average European roll out speed (dashed line) and min max range (shaded area) from 01/02/2021 to 20/06/2021. **h-j**: Onset of immunization: Effect of shift between introduction of infection and start of immunization at 1%/day on **h** attack rate, **i** death toll and **j** population immunity for the strategy *random*. All simulations are based on the German municipality Gangelt with 10.351 agents. Parameter values: infectivity *k*_*I*_ = 0.15, interaction frequency *μ* = 2.

For high vaccination levels (here more than about 60% of the population is immunized), the strategy to vaccinate the *most interactive* individuals first is most effective for all four objectives. For lower vaccination levels, we identify a clear tradeoff between different strategies depending on the objective, i.e. attenuation of the infection wave, preventing fatalities, or avoiding ICU overload. Aiming for a reduced number of infections, it is most effective to prioritize the *most interactive* individuals, as it reduces the probability for an emerging infection wave and, thus, increases systemic robustness. It outperforms vaccination by forecasting of infected individuals in a pre-simulated baseline scenario. Random vaccination underperforms compared to the other two strategies, in agreement with^7,40^, but outperforms *sorted-by-age* vaccination. While most strategies reduce the fraction of infection down to zero at 90% vaccination or lower, this is not achieved by *age-sorted* vaccination. The reason is that young individuals always keep interacting, leading to high connectivity among the remaining susceptibles. In general individuals of the same cohort form sub-networks that remain unperturbed by vaccination of other individuals who do not belong to that same age group.

To reduce fatalities, the age-sorted strategy is very effective at low vaccination levels and is only outperformed by the “combined” strategy (**Supplementary Fig. 24**). However, for the high vaccination levels at which other strategies display population immunity, those strategies surpass both strategies, vaccination by age (first the interactives, then the forecasted and random strategies), since this strategy is not able to suppress deaths completely before 100% population immunity is reached. The “combined” strategy integrates the two strategies that either best reduce infections (i.e. by interaction) or death toll (by age), however it outperforms neither.

The simulations also clearly reveal a problem of strategies that focus on vaccination only (**Fig. 3d**): with the objective to reduce the occupancy of ICUs, vaccination by age (or the combined strategy) performs best at less than ∼40% but vaccination by interactivity is best above this level. However, below ∼50% vaccination, none of these strategies is able to prevent overload of ICU capacity, without additional NPIs. Importantly, our model has not implicitly included an increased death rate if ICUs are overloaded. Hence, the death toll would be even higher than predicted if ICU demands cannot be met. While the ICU capacities may vary in different locations, the problem remains that ICU demand and capacity differ widely for all strategies at stages of partial vaccination. This implies NPIs remain to be considered to accompany the vaccination process in order to prevent the collapse of the healthcare system.

For the random strategy the attack rate approaches zero and the likelihood for a subsequent infection wave is halved at the herd (population) immunity threshold (HIT) calculated for the corresponding R_0_, showing that our approach is comparable to the established methods^41^ for random vaccination and that different vaccination strategies can drastically alter the outcome.

In summary, as long as we cannot ensure vaccination of about 60% of the population, it is not possible to serve all objectives to reduce deaths, ICU demand and infection levels at the same time equally well with vaccinations only. Above this value, vaccination of individuals sorted by their interactivity shows to be most successful for all three objectives.

We observe bimodality also for vaccination, i.e. systematic vaccination of the population can lead to either termination or recurrence of outbreaks with a certain likelihood - depending on strategy in the range of between 60% and 90% people vaccinated (**Fig. 3b**). This means that in these ranges it depends essentially on luck whether a community will experience another wave. During these “windows” - either periods of time or sets of NPIs or ratios of vaccinated individuals -, decision makers have to adapt and **(i)** use precision simulations to see whether a community is in such a window or if the situation is controlled, and **(ii)** combine vaccinations with NPIs in order to at the same time keep the outbreak down and ease the conditions for the population by preventing unnecessary lockdowns or school closures.

As a result, there is not a single number for a vaccination percentage <100% ensuring population immunity. Instead, this number depends entirely on the chosen strategy. Importantly, it also depends both on the heterogeneity of the population and on the specific virus, its virulence and infectiousness. In particular, lower infectivity will decrease the required vaccination coverage to achieve population immunity (**Supplementary Fig. 25**). Note that the attack rate and the population immunity threshold (**Fig. 3a**) are different concepts, which was also shown for SARS-CoV-2 spreading in Brazil, where >70% of a population has been infected during an uncontrolled outbreak, a fraction above the reported theoretical population immunity threshold of below 66% for R_0_ < 3 ^29^. Our model predicts that depending on the vaccination strategy, the attack rate can be smaller or even larger than the population immunity threshold.

Since vaccination is not mandatory in most countries, high levels of vaccination of the population can only be reached if people comply. Lack of commitment of the population would be partially comparable to lack of compliance to NPIs (**Fig. 1k**): if 25% of the population refuse vaccination (irrespective of the reason), the effects are similar to non-sufficient dose numbers with the same strategy-dependent effects on infection spreading, ICU overload, or fatalities.

### Reaching and maintaining population immunity requires adaptive combinations of NPIs and vaccination strategies

To demonstrate the capabilities of the model in the light of real-world data, we compared the simulated course of infection between early January 2021 and end of May 2021 in the municipality Gangelt to data from the enclosing district Heinsberg and Germany (**Fig. 4**). Here, we considered **(i)** the incidence value in the area in early January 2021 according to RKI data, **(ii)** the appearance of the β(UK)-variant and its roughly linear relative increase in the infected population, and **(iii)** the increasing immunization rate (2nd vaccine dose) in Germany. We modelled vaccination according to *age* as described above since this best matches the situation in Germany during that period. Under these conditions, the simulations again exhibit bimodality (subsiding infections in about 40% of the simulations, a new wave of infections in April in the other simulations). The dynamics of the five-day rolling average of reported daily new cases (**Fig. 4 b, c**) matches the rolling average of the simulated newly diagnosed individuals and the relative occurrence in different age cohorts **(Fig. 4g)** in simulations showing a new infection wave. Moreover, even the value, tendency, and weekly oscillation of the derived R-value (**Fig. 4d**) are in agreement with the reported estimates for R, too.

**Fig. 4:**
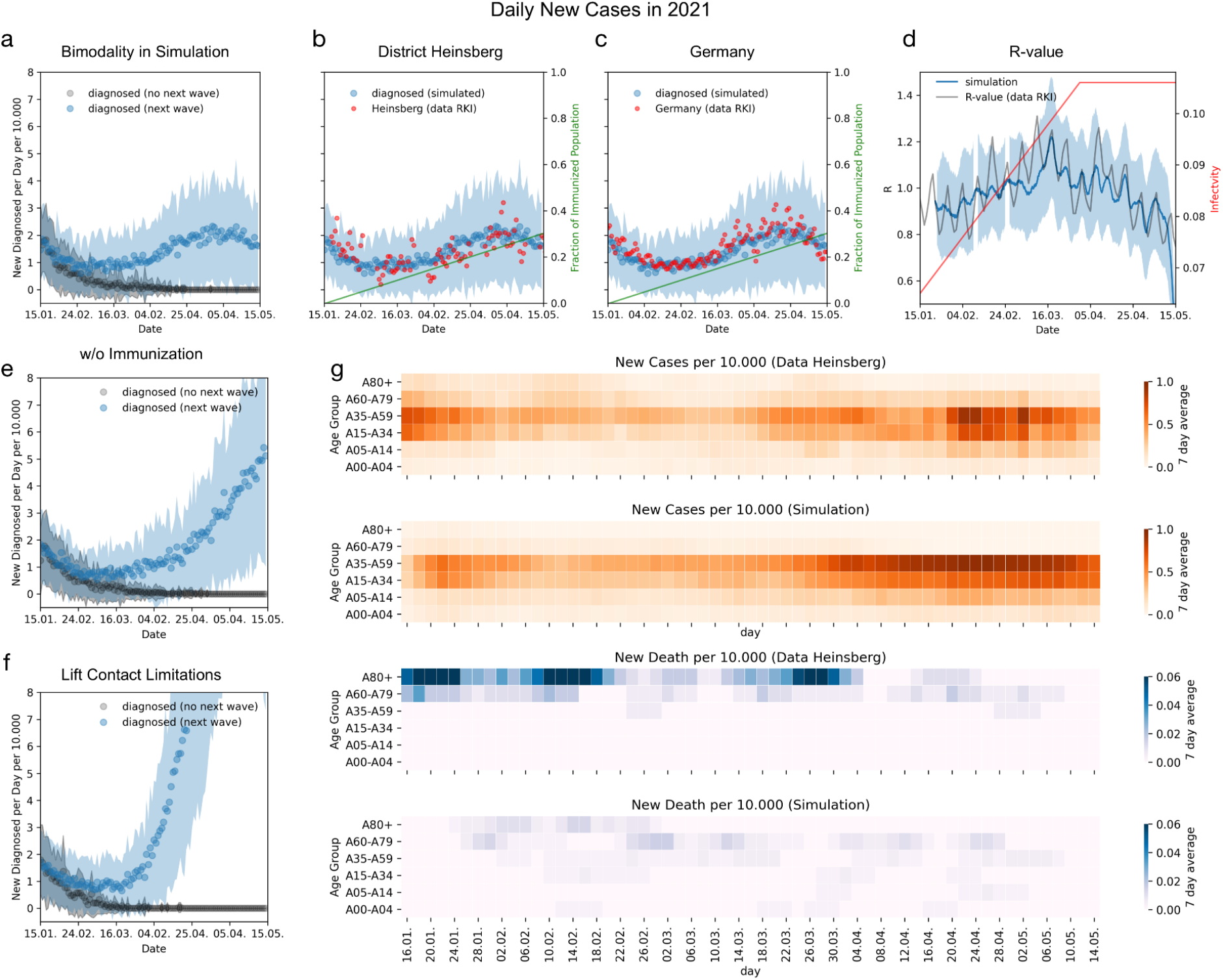
Comparing adaptive interventions with real-world epidemiological data. Simulations demonstrate the combination of NPIs and vaccinations as well as the predictive power of the model reproducing the infection dynamics in Gangelt from 01/15/2021 to 05/15/2021 in comparison to data from the enclosing district Heinsberg and all of Germany. Simulations started with 117 infected individuals (corresponding to the incidence numbers on 01/15/2021) and drastically reduced contacts (by 75% at public places, by 75% at schools, and by 40% at workplaces) representing the applied NPI (lockdown). The increase in prevalence of the β-variant from 10% to 90% during that period was modeled by a linearly increasing infectivity until day 75, assuming a 50% higher infectivity of the β-variant. To reflect vaccination individuals were immunized by decreasing age (0.25% per day starting with the oldest). **a** Simulated daily new cases of (diagnosed) individuals, exhibiting bimodality by showing either a new infection wave (blue; mean +/- standard deviation; next wave here for 54 out of 96 simulations) or extinction of infection (black, mean +/- SD, 42 out of 96 simulations) after the initial decline in cases. **b/c** Comparison of the (non-extinct) simulations from **a** to data for Heinsberg (b) or Germany (c) (red dots: 4-day-sliding window per 10000 inhabitants). Green line indicates the fraction of immunized individuals. **d** Comparison of calculated (blue, mean +/- SD) and reported R-values (gray line) for Germany, exhibiting weekly oscillations. Red line indicates the development of simulated infectivity. **e/f** Tests of the effect of **e** immunization and **f** contact limitations on the epidemic progress still exhibited bimodal outcomes. Simulations without vaccination (**e**) show infection waves in 50 out of 96 cases and simulations with lifted NPI (**f**) in 66 cases of 96, respectively. **g** Comparison of simulated spread of detected new cases and fatalities within age cohorts with data obtained for the Heinsberg. Parameter values: infectivity *k*_*I*_ = 0.065, interaction frequency *μ* = 2.

Note that a few aspects not considered here can have influence on the real-world trajectory. The assumed roll-out strategy *age* is an idealized version of the strategy applied in Germany, leading to strong reduction of infections in the cohorts 60+. Data show that also vulnerable individuals of other age groups and health care persons were vaccinated. Consequently, we obtained an underestimation of the death numbers since the 60+ cohort has a significantly increased fatality rate. This, in turn, demonstrates the effectiveness of *age* strategy to reduce fatalities. Furthermore, fatalities occurred in the data during the simulated period (**Fig. 4g**) may also be the consequence of prior infection events and thus not captured by the simulation.

The increasing rate of testing, the potential seasonal dynamics of the virus^42^, and the exchange of infected individuals with other communities may also play a role. Since no extinction of local infection waves was observed in the various German districts in this period (no systematic data for individual municipalities), exchange of infected individuals may have stabilized the infection wave.

Using the model with infectivity adjusted to the real course of infection in the period Jan-May 2021, we show the effect of individual NPI by not including either vaccination (**Fig 4 e**) or NPIs (**Fig 4f**). It is obvious that both NPI and increasing numbers of vaccinations were relevant in keeping the infection wave under control, where lifting of NPI had a stronger effect in the considered period.

## Discussion

Vaccination of the human population against COVID-19 is an immense logistical challenge that necessitates careful prioritization in order to swiftly reach maximal suppression of the disease and also save lives^43^. Limits on production and distribution renders it absolutely necessary to prioritize and structure the vaccine deployment^39,44,45^. The European Commission concluded that “the successful deployment and a sufficient uptake of such vaccines is equally important” rendering it critical to be able to “monitor the performance of the vaccination strategies”^46^. Thus, the challenge for the scientific community is to develop platforms that can provide precise, context specific and adaptive predictions of the quantitative effect and requirements of such strategies in close to real-time. Here, we have responded to this challenge by developing a data-driven geospatial, temporal, network-based model of integrated individual human behavior. This made it possible to quantify non-linear effects of NPIs and compare the impact of targeted immunization strategies. Remarkably, the heterogeneous model offers insight into the bimodal behavior of SARS-CoV-2 infection dynamics and demonstrates that effective interventions require strict execution (stringency) and careful temporal control (timing). We demonstrate that COVID-19 infection networks (iHHIN) are sparse and small compared to the overall population interaction network (HHIN). Therefore, models based on homogenous mixing or averaging statistical models are likely to be of limited use^23,24^ especially during phases of low to moderate infection or emergence of new variants, since they fail to capture heterogeneity, nonlinearity, and complexity emerging from stochastic and sparse events influenced by individual human behavior ^15–17,47^.

Our work shows there is a tradeoff between different strategies for low levels of vaccination: vaccination by age minimizes fatalities, while vaccination by interactivity reduces infection events. However, at high vaccination coverage, vaccination by interaction prevails. It is important to note that the vaccination level giving rise to population immunity is not a unique number but depends on the chosen vaccination strategy. These conclusions depend on the demographic structure and the heterogeneity in the interactions and it can be assumed that the stronger the heterogeneity the better vaccination by interaction will perform.

Targeted immunization “into” an ongoing outbreak may become relevant as we enter the fall of 2021. This approach likely performs differently to other strategies, since infection spreading might already reach the vulnerable subgroups and spread further in these sub-networks. Vaccination of the population is a process in time, especially in the global context. But locally, significant vaccination coverage may be achieved fast in some countries or regions. The optimal vaccination strategy depends on the supply of vaccines, the demographic structure, local behavioral costumes, and the capacity to realize a specific strategy. In future, forecasting of the effect of vaccination shall be combined with prior simulation of the ongoing surge of infections and the effect of hitherto applied NPIs to precisely model the situation in specific communities at the time when vaccines become available.

We propose that our work suggests that in order to reach a post-Covid world, communities and governments world-wide will have to deploy adaptive and real-time based simulation support for decision making. This is the only path that can ensure continued education, economic and research activity, healthcare and other society functions during new outbreaks by minimizing the catastrophic socio-economic impact of lockdowns, travel bans, and civil non-compliance. To fully achieve this, new paradigms for modeling of infection networks that capture the nonlinear complexity and stochasticity will be important, beyond what we have demonstrated here.

### Online Methods

The detailed concept of geospatial demographic heterogeneous agent-based model, the estimation of parameters for the stochastic transitions between states, and additional simulation results are represented in the **Supplementary materials**.

### Model Design - Locations, Agents, Health states

To reduce complexity of the HHIN, each human individual (agent) is associated with a specific physical location at each time point. These *locations* are specific for the community such as homes, workplaces, schools, hospitals, and public places^48^ (**Fig. 1b**). Since transmissions outdoors or in public transport are of less importance^49^, these places are not explicitly included, although movement between locations is included. The entire *population* is initialized with demographic census data resulting in representative age distributions and household compositions. An individual is defined by its household H ICU membership, age, weekly schedule, and health status. The weekly schedule (hourly resolution, discriminating between weekdays and weekends) specifies the individual’s presence in different locations (**Fig. 1c-e**). Schedules change with health states and with NPIs. The health states for individuals are defined as: susceptible (S), infected (I), recovered (R) or deceased (D). Infected individuals (I) can obtain sub-states specifying their condition as pre- or asymptomatic (plain I), diagnosed (I^d^), hospitalized (I^d^), or being in an ICU (I^d^) (**Fig. 1f**).

### Model Dynamics - HHIN and Infection Transmission

When two agents are present at the same location at the same hour (co-location network), they can interact with a probability that depends on the number of present agents and the parameter interactivity *µ*, which yields the human-human interaction network (HHIN). If two agents interact and one of them is infected and the other one susceptible, then the infection can be transmitted with a probability that depends on the time-dependent infectiousness of the emitter and on the parameter infectivity *k*_*I*_ (tunable according to NPI such as mask wearing or distance keeping), establishing the iHHIN.

### Data Integration and Model Initialization

All data are taken from publicly available databases^32,33^ (**Figure 5**). As a basis, we considered location information and information about agents. Model initialization ensures that we simulate the dynamics in a community with a number of agents that is about the same as the number of inhabitants of that community, representing the statistical information w.r.t. to age, household composition, and occupations. For the infection process, we integrated information from the German Robert Koch Institute (RKI) factsheets^50,51^ and from recent publications^31,34^ to derive hourly conditional probabilities for transitions between the health states. The model has only two parameters that can be freely chosen or adjusted to observations, i.e. the infectivity *k*_*I*_ and the global interaction frequency *μ*. (Detailed description see **Supplementary Material**).

**Fig. 5:**
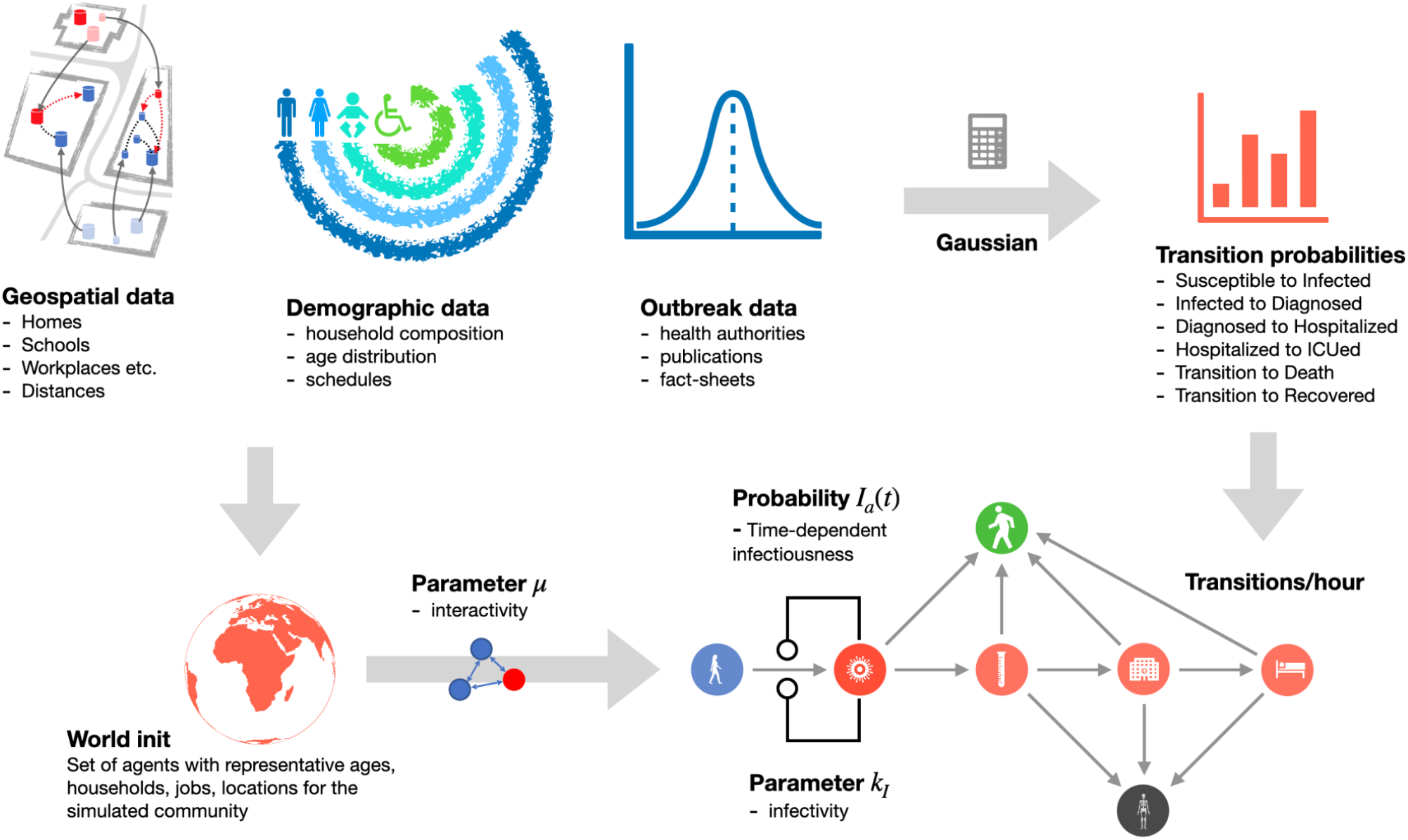
Data flow-diagram. The data required to simulate a community includes geospatial information about types and position of locations such as homes, workplaces, schools, public places and hospitals. Demographic data is used to derive attributes of the agents such as household affiliation, age and schedule. Both data types together are used to initialize the model (World init). Information about the outbreak is used to collect health state transition probabilities (cumulative probabilities for transitions, e.g. from being diagnosed to getting hospitalized). The cumulative probabilities are transformed with Gaussian probability mass function into age-dependent, time-dependent conditional probabilities for the respective transition (e.g. the likelihood of an agent belonging to a specific age group getting into hospital at a specific hour after being diagnosed if that agent has not already recovered or got hospitalized before). From the outbreak data, we also derived the time-dependent infectiousness *I*_*a*_(*t*) of infected agents (in short, it peaks on day 2 after infection before symptoms onset on day 3) with hourly resolution. The model has two global parameters that can be varied and adjusted to data, i.e. the infectivity *k*_*I*_ and the global interaction frequency *μ. μ* determines how likely it is that two agents being at the same location at the same time interact. If they interact, the product of the agent-specific *I*_*a*_(*t*) and the global *kI* determine infection transmission.

### Model Simulation

During simulations, individuals move between locations according to their schedules in an hourly resolution. The individuals’ presence at locations, interactions with other individuals, health states, and infection transmissions are recorded at each time step. This makes the stochastic HHIN and iHHIN traceable and amenable to theoretical analysis and implementation of different interventions such as (time or location-specific) lockdowns and vaccination. In order to evaluate the effect of NPI and vaccinations, we simulated a baseline scenario representing an uncontrolled outbreak (**Fig. 1g,h**). The history of infection events defines the basic reproduction number (R-value), and is, thus, an emergent property of our model (**Fig. 1i**). To implement NPIs, we modify the schedules (e.g. stay at home instead of visits to the workplace or public places during lockdown) (**Fig. 1j**), which alters the course of infection. Also the level of compliance in the population with interventions influences their effect, e.g. on infection numbers in a manner consistent with reality (**Fig. 1k, Supplementary Fig. 10**). Importantly, simulations of specific interventions reveal bimodality, i.e. lead to qualitatively different outcomes for repeated simulations with identical initial conditions (the infection ceases in some simulations, while generating a strong second wave in others, **Fig. 1l**).

## Data Availability

All data used are included in the supplementary material.

## Code Availability

The code is available at our GitLab repository (https://tbp-klipp.science/GERDA-model/) which also contains a manual for the application of the method to other municipalities including the required types of data.

## ACKNOWLEDGMENTS

We wish the populations of Gangelt, Heinsberg, Epping, Vaxholm, and Zhikron Ya’akov all the best and would like to point out that the model remains an abstraction and it is neither a repetition of history nor can it be traced back to real individuals, thus safe-guarding the privacy of the public.

## Author contributions

Conceived the project: RL, EK. Conceptual work on the model: BG, SOA, OB, JAHW, XEF, RL, EK. Programming: BG, SOA, OB, JAHW, AK, LB, JELH, MK^a^, RMT, MS. Analysis and computational experiments: BG, SOA, OB, JAHW, XEF, AK, MK^a^, RMT, MS, EK. Data collection: XEF, JELH, MK^b^, LM, HP, PSS. Wrote the paper: BG, OB, JAHW, XEF, MK^b^, MS, RL, EK. All authors agreed on the final version of the manuscript.

## Data and materials availability

All data is available in the main text or the supplementary materials.” The code is available at https://tbp-klipp.science/GERDA/code/

## SUPPLEMENTARY MATERIALS

- Materials and Methods
- Figures 1-26.
- Tables 1-26.
- Movie 1.

