## Supplementary Material for "Adaptive combination of interventions required to reach population immunity due to stochastic community dynamics and limited vaccination"

Supplementary Material

1 Introduction

2 Model Design

2.1 Locations

2.1.1 Location Types

2.2 Agents

2.2.1 Agent Properties

2.2.2 Agent Operations - Schedules and Occupation

2.2.3 Motion Between Locations According to Schedules
2.2.4 Agent-agent Interactions
2.2.5 Infection Transmission
2.2.6 Health States
2.2.7 Health State Transitions
3 Model Initialization and Data Integration
3.1 Location Initialization
3.1.1 Principles of Location Initialization
3.1.2 Data used for Location Initialization
3.2 Household Initialization
3.2.1 Principles of Household/Population Initialization
3.2.2 Data for Household Initialization
Algorithm
3.3 Agent Initialization
3.3.1 Principles of Agent Initialization
3.3.2 Schedule Generation for Agents
3.4 Health State Transitions
3.4.1 Principles
3.4.2 Input Data for Health State Transition Rates
3.4.3 Probability Calculations
3.4.3.1 Age-independent state transition probabilities
3.4.3.2 Age-dependent state transition probabilities
3.4.3.3 Time-Dependent Conditional State Transitions
3.5 Infectiousness of an infected agent
3.6 Additional Parameters
4 Simulation and Analysis of GERDA
4.1 Simulation time course and outputs
4.1.1 Simulation object at final time step

4.2 Simulation of specific scenarios

4.3 Sensitivity analysis

4.4 Network analysis

4.4.1 Interaction networks

4.4.2 Infection networks

4.4.3 Overrepresentation and underrepresentation of specific groups in the infection process

4.5 Immunization Scans

4.6 Comparison to Homogeneous Mixing Approach

4.7 The Movie

5 References

### 1 Introduction

Severe acute respiratory syndrome coronavirus 2 (SARS-CoV-2) has been globally affecting societies and economies for more than a year and classical strategies have not been successful to prevent further spreading of the virus. To support evidence-based decision making during this ongoing crisis, the scientific community has made great efforts to analyze the available data (selected reviews: (*1*–*5*)), to predict virus propagation and the influence of different societal aspects, such as age or contact levels (*6*–*8*), to analyze the underlying networks of outbreaks (*9*), and to provide scientific evidence for the efficacy of different mitigation measures, testing or vaccination strategies (selected reviews: (*10*–*12*)). With the Global Epidemic and Mobility (GLEaM) framework there is a stochastic computational scheme that integrates worldwide high-resolution demographic and mobility data to simulate disease spread on the global scale (*13*). Still, in a recent comment, Bouffanais and Lim pinpointed the discrepancy between the detail levels of available data and mathematical models versus the requested and required information to effectively prevent uncontrollable virus spreading (*14*).

Classical SIR-models (*15*, *16*) are commonly based on population-wide assertions, thereby omitting the variability of the infection transition process between individuals. Traditional agent-based models (*17*) capture stochasticity in agent behavior, but are highly limited in size by computing capacities. Others have attempted to overcome these limitations: Already in 2005, Ferguson et al. (*18*) published an agent-based model considering statistical distributions of transmission events in households, building types and alike, while Chao and coworkers considered data on personal contacts back in 2009 (*19*). More recently, (*20*) simulated infection transmission throughout Bogotá representing the city's population by 1000 agents and (*21*) created a simple model to analyze the effect of traveling between cities in China, but without realistic individual agents. Quilty et al. (*22*) predicted quarantine and test strategies. Still, they all attempt to describe the infection process on population level, thus relying on a coarse-grained representation by a limited number of agents. Alternatively, (*23*) developed an ODE model that describes increased social distancing by decreasing the basic reproduction number (R value). (*24*) modeled presymptomatic and non-diagnosed transmissions, (*7*) provided valuable age specific data on contact patterns in the Chinese population, and (*8*) assessed the effect of alternative working schedules. (*25*, *26*) estimated the rate of detection of symptomatic cases of COVID-19 in France after lockdown and suggested school reopening scenarios using data coupled with mathematical transmission models. Mistry et al. (*27*) derived contact matrices to model virus spread and show that sub-national heterogeneities in human mixing patterns, however they employed ODE whereby theneglected the derived detailed household structures again. Also (*22*) derived contact patterns of UK adults before and during lockdowns from questionnaires.

While certainly all of those models administer important information for decision processes during a pandemic, they still focus on population dynamics, i.e. the macroscopic view, and consider only specific aspects of the virus transmission process. In contrast, virus outbreaks are fundamentally non-linear, stochastic, and highly network-based. Additionally, the propagation between human individuals depends on the specific geospatial and demographical context. Consequently, quantitative assessment of non-linear integrative effects might be key to developing suitable intervention and vaccination strategies (*6*, *14*, *28*). To efficiently predict virus propagation on a more specific, not necessarily global level, we require heterogeneous models that take into account not only age cohorts and social activity levels but many other types of heterogeneities such as households (of different sizes) or different types of locations, like schools or workplaces (*6*).

We developed a new integrative concept for modeling of virus spreading that focuses on individual virus transmission events in a defined community based on real-world data. To this end, we complement an expanded SIR model with information on heterogeneity in the population. The resulting GEoReferenced Demographic Agent-based (GERDA) model allows simulation of a multitude of consecutive and comparative scenarios providing locally specific information on infection hubs and a variety of population subgroups. Showing bimodal simulation outcomes for the same scenario proves GERDA's capability to capture the stochasticity of the virus transmission process. Comparative simulations allowed us to evaluate the efficacy of different vaccination strategies. Finally, the ability to adapt simulation scenarios to the current situation in real-time, the potential applicability of the approach to any virus, and the capability to deal with various levels of detail for the different input data, render this approach highly valuable in the current but also in future pandemics.The model entities, parameters, and other important symbols used throughout this document are summarized in **Table S1**.

| **Symbol** | **Name** | **Detail** | **Property/Dependency** | **Type** | **Reference** |
|---|---|---|---|---|---|
| MW | Modeled world | Reflects real-world community/town, composed of locations and agents | Custom | Model object | |
| loc | Location | Container for agents with geographical coordinates, reflects, real-world building or open space | workplaces, schools, public places, hospitals, morgue, and households or mixing location | Model entity | OpenStreetMap (OSM) |
| | Agent | Reflects human individual | agent ID, home location, age, schedules, health state | Model entity | **https://ergebnisse.zensus2011.de** |
| $n_{loc}$ | Agents per location | Number of agents present at a specific location at the same time | Location type, location size, time, schedules | Time dependent simulation output | |
| μ | Global interaction frequency | Mean interaction frequency of agents | | Input parameter | Wallinga et al., 2006 |

| | | | | | |
|---|---|---|---|---|---|
| $P^{int}_{ab}$ | Interaction probability | Interaction probability of two agents a and b per time step at specific location | M, $n_{loc}$, $x_a$ , $x_b$ and μ | Emerging parameter | |
| M | Interaction matrix | Matrix coefficients specify the probability with which any pair of agents may interact at a given location and time ($x_i$ $x_j$) | Interactivity, schedule, location | Emerging parameter | |
| $k_I$ | Global infection | Reflects the overall infectivity | User dependent | Input parameter | |
| $P^{inf}_{ab}$ | Infection probability | Probability of an infected agent to transmit the virus to a susceptible interaction partner | Age, infected agent time of infection | Input parameter | He et al., 2020 |
| $X_{ij}$ | State transition | Infected agents undergo various health state transitions reflecting disease progression until they either recover or die. $X_{ij}$, $S_i \rightarrow S_j$ with $S_i$, $S_j$ being health (sub-)states | Si health (sub-)states, state transition probabilities $P^{X}_{ijt}$ | Model entity | |
| $P^{c}_{ij}$ | Cumulative transition probability | Transition probability, with i, j being health (sub-)states. | | Input Parameter | RKI Factsheet; Karagiannidis et al., 2020 |
| $P^{X}_{ijt}$, $P^{X}_{ijT}$ | Transition probability | Time and age dependent gaussian distributed transition probabilities with i, j being health (sub-)states and time t | | Input Parameter | |
| $p^{X}_{ijt}$, $p^{X}_{ijT}$ | Conditional transition probability | Time and age dependent conditional transition probabilities with i, j being health (sub-)states and time t | | Input Parameter | |
| | Infectivity distribution | Poisson distribution reflecting infection time dependent differences in infectivity | Time of infection | Input Parameter | He, et al., 2020 |
| t, T | Time | Discrete time step (per hour) | | | Mossong et al., 2008 |
| S or $S_1$ | Susceptible | Default state. Agents have never been subject to the infection and neither are immune to it | Simulation, state transition probabilities | Health state | |
| I or $S_2$ | Infected | Comprise presymptomatic, asymptomatic and symptomatic infected individual | Simulation, state transition probabilities | Health state | |
| $I^d$ or $S_3$ | Diagnosed | Recognized infections including the cases with or without symptoms, irrespective of how they were identified. | Simulation, state transition probabilities | Health (sub)state | |
| $I^d_H$ or $S_4$ | Hospitalized | Cases of moderate severity, hospitalization on the normal ward | Simulation, state transition probabilities | Health (sub)state | |
| $I^d_{ICU}$ or $S_5$ | In ICU | Agents that need treatment in an intensive care unit comprising the critically severe cases | Simulation, state transition probabilities | Health (sub)state | |
| R or $S_6$ | Recovered | Agents have survived an (either asymptomatic or symptomatic) infection, are not infectious anymore, and are considered immune | Simulation, state transition probabilities | Health state | |
| D or $S_7$ | Dead | Condition of the agents that have not recovered from the infection and died | Simulation, state transition probabilities | Health state | |

| | | | | | |
|---|---|---|---|---|---|
| | Regular schedules | Regular behavior of agents. A schedule covers 7 days (168 hours). | Age, occupation, health status | Model entity | |
| | Specific schedules | Schedules that determine agents (restricted) movements under defined circumstances. This comprises diagnosed, hospital schedule, death, non-pharmaceutical intervention | Health status, non-pharmaceutical interventions, locations, non-compliant probability | Model entity | |
| $x_a$ | Interactivity | Probability for any agent a to interact with any other agent | Custom | Input parameter | |
| $p^{inf}_{ab}$ | conditional probability for infection transmission | Infection transmission probability given an interaction occurred between an infected agent a and a susceptible agent b | Infectivity, infectivity distribution, time | Input parameter | |
| $I^I_a(t)$ | Infectiousness | Infectiousness of a given infected agent a at time t | Time since infection, infectivity, infectivity distribution | Emergent parameter | |
| $f^{loc}$ | Location-specific factor | Represents the infection risk at specific location type loc | Custom | Input parameter | |
| $y^{inf}$ | Specific transmission coefficient | Specific coefficients for infected agent modulating the transmission probability between interacting agents | Custom | Input parameter | |
| $y^s$ | Specific transmission coefficient | Specific coefficients for susceptible agent modulating the transmission probability between interacting agents | Custom | Input parameter | |
| $P^{S}_{iT}$ | Remaining in state probability | Probability for being in state Si until time T | Probabilities to leave the state $S_i$ before T | Emergent parameter | |
| $P^t_{ij}$ | Remaining in state probability | Probability for staying in state $S_i$ at time t | | | |
| $N_i$ | age-resolved case numbers | number of cases in an age group | | Input Data | RKI Factsheet |
| $\mu^t_{ij}$ | State transition mean time | The probability for each state transition $X_{ij}$ is Gaussian distributed with mean at the reported or assumed average time t. | | Input parameter | an der Heiden & Buchholz 2020 |
| $\sigma^t_{ij}$ | State transition standard deviation time | The standard deviation of Gaussian distribution equals its mean | | Input parameter | |
| PMF | Probability mass function | Function that gives the probability that a discrete random variable is exactly equal to some value. | | Mathematical function | |
| $p^{ncomp}$ | Non-compliance probability | Reflects (sub)population fraction not complying with non-pharmaceutical interventions | Custom | Input parameter | |
| $t^{trace}$ | Tracing time | Time frame in which retrospective contact tracing takes place | Custom | Input parameter | |

**Table S1: Symbols and abbreviations reference table.** Description and dependencies for the different model entities and calculations of this document.

### 2 Model Design

To investigate the infection dynamics in a real-world community, e.g. Gangelt in Germany, a simulation environment is required that reflects local peculiarities, like amount and position of buildings and outdoor locations or size and demographic composition of the population. Furthermore, it is important to account for individual behavior of the members of the modeled population, like household interactions or (age-dependent) daily occupations. Finally, specifically in the context of an acute pandemic, the possibility to evaluate the effectiveness of non-pharmaceutical intervention strategies is required. Here, we provide a detailed documentation of the design and computational setup of GERDA, including a description of how we made use of external data and of the pipelines used to convert data into model input. Below, the design principles of GERDA are summarized, followed by sections about the parametrization and the computational analysis.

The GERDA model has been designed with three main goals: i) Simulation and analysis of transmission events, e.g. SARS-CoV-2 virus transmissions; ii) Applicability to different communities/Consideration of local peculiarities for simulations; iii) Creation of a test environment for the effect of (non-pharmaceutical) interventions. Those goals directly translate into three major design principles: first, the agent-based modeling approach, second, easily exchangeable external data input files, and third, a strict separation of a) model initialization, b) model parameterization, and c) model simulations. Those principles have been consistently applied throughout the entire development process.

The major object of GERDA is a so-called modeled world (MW) that provides the basic structure required to simulate virus propagation throughout the community. The MW is composed of locations (reflecting real-world buildings and outdoor spaces) and inhabited by agents (representing human individuals). To assure a realistic representation, locations and agents are initialized based on external input data and connected via households at model initialization. According to their schedules, agents visit locations at which they can interact with each other during a simulation. Upon interaction, an infection is potentially transmitted between individual agents. Infected agents can undergo various health state transitions reflecting disease progression until they either recover or die.

#### 2.1 Locations

In the MW and throughout simulations, locations serve as containers for agents and, thereby, define the interaction topology in which agent-agent interactions and virus transmission take place. Hence, locations represent the spatial structure of the model. Every location is assigned to a predefined location type and has its spatial coordinates stored to allow distance-related distinction of locations for agents. Sets of locations in spatial proximity can be clustered into neighbourhoods to further structure the MW. An outstanding feature of GERDA is the possibility to generate locations on the basis of geographical data (Sec. 3.1) to reflect the interaction and infection dynamics of real world communities. Alternatively, locations can be generated according to alternative principles or arbitrarily for comparison with a randomized case.

The assignment of locations into predefined location types allows agents to visit appropriate locations according to their specific daily or weekly routines. Furthermore, it provides the possibility to adjust particular model parameters for a specific location type, for example when testing non-pharmaceutical intervention strategies applied only to schools.

##### 2.1.1 Location Types

In the current version of GERDA, seven predefined location types are defined, i.e. workplaces, schools, public places, hospitals, morgue, homes and a mixing location, each with unique and common properties.

Workplaces are locations which are regularly visited by the agents assigned to the group of employees during their schedule-defined work hours. The work place of an agent is chosen with a likelihood inverse to the spatial distance to the home assigned to that agent. Pure workplaces represent, for example, office buildings, farms, or factories.

Public spaces can be visited throughout the day by all agents, which are not restricted otherwise (e.g. being in hospital). Additionally, they are assigned to a fraction of the agents representing employees as workplaces. The location type public space stands for a variety of different real world locations, such as parks, city halls, or restaurants.

Schools are daily visited by underage agents throughout the weekdays and are assigned to teachers as location of work. A specific location of type school is chosen for each underage agent according to the least distance to its assigned home location. In the current version of GERDA, schools comprise among others: kindergartens, elementary schools and high schools.

Hospitals are locations with two functions. On the one hand, they are assigned to medical personnel as workplaces and, on the other hand, they serve as containers for agents with severe or critical disease progression, i.e. being in the states $I_H^d$ or $I_{ICU}^d$ as explained below.

Morgue is the unique location foreseen for “deceased” agents. This location is required to prevent non-realistic interactions with agents of that status while keeping the agent number of the MW constant. At least one hospital and one morgue are required in any MW to assure functionality of the model, and they are automatically added if they are not present in the modeled community.

Home is the location type, which serves as a reference location for each modeled agent. Each agent is assigned to only one home, but one location of type home can serve as reference to more than one agent. Agents with the same home are referred to as one household.

The mixing location is an artificially introduced unique location. It is not required for real-world-related simulations, but is used to compare with simulations of a homogeneously mixed world.

#### 2.2 Agents

The population of the MW consists of agents representing human individuals. The population can not be set independently but instead results from the number of created locations of type home, i.e. the households. Based on the number of households and the demographic input data (described 3.2), the model generates a population of agents which represents realistic distributions of ages and household sizes.

##### 2.2.1 Agent Properties

Through the household initialization process, each agent obtains an age in years and an age-related occupation. Occupations resemble child/student for younger agents, employees in middle ages, and pensioners for older agents (more details below). Each agent is uniquely identifiable by an ID.

Agents have two major properties that define their potential operations: a weekly schedule and a health state. The schedule specifies the agent's time-dependent presence at locations and, thereby its potential to interact with other agents and become infected. The health state can change depending on interactions with infected individuals or during the disease progression. It modifies the potential operations of the agent leading to altered schedules. GERDA includes four main health states, namely susceptible (S), infected (I), recovered (R), and dead (D) with the default health state S (**Figure S1**).

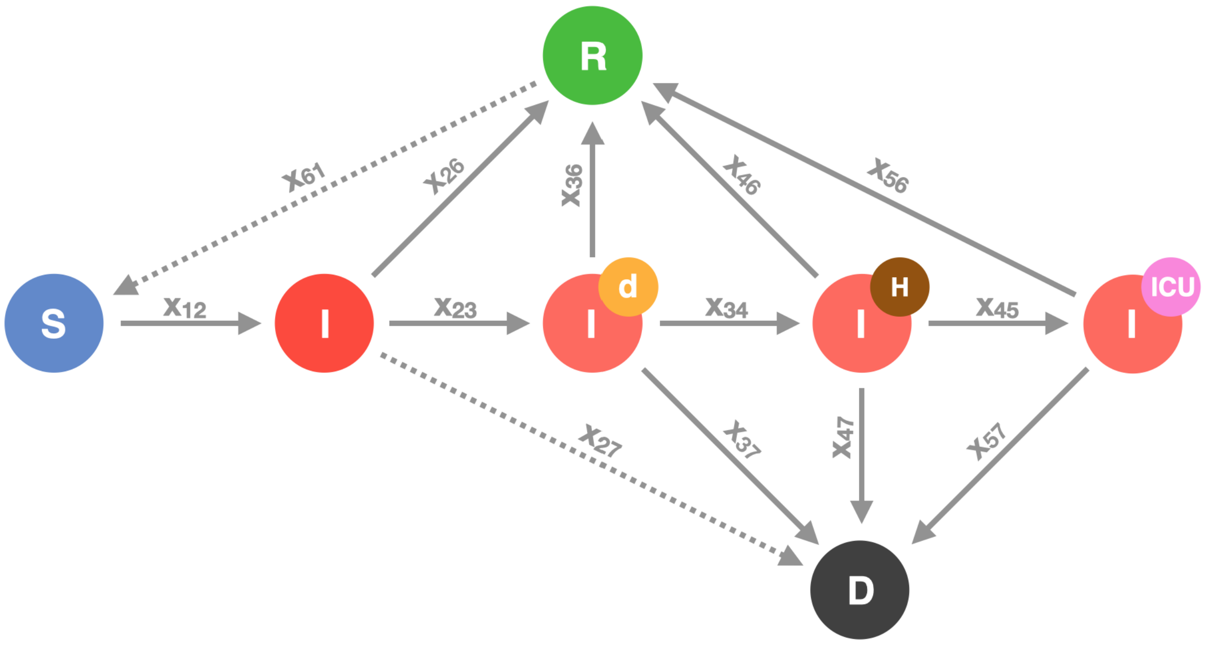


**Figure S1**: **Health states and potential transitions.** Agents can be susceptible (S), infected (I), recovered (R), or dead (D). The group of infected agents comprises the presymptomatic or asymptomatic infected (plain I), the diagnosed infected ($I^d$), the hospitalized infected ($I^d_H$) and the infected requiring intensive care unit ($I^d_{ICU}$). The transitions indicated by solid arrows are implemented, the transitions indicated by dotted arrows are foreseen, but not used in the current simulations (probabilities set to zero).

Below, we describe first the schedules and the resulting potential interactions between agents. Then, we introduce the infection transmission process, which links the concept of schedules and locations with the concept of health states. Eventually, we explain the considered health states and transitions between them in detail.

#### 2.2.2 Agent Operations - Schedules and Occupation

Every agent has a weekly schedule, which is assigned based on age and a given schedule distribution for the corresponding age group during agent initialization. This schedule determines the locations visited by the agent over the period of one week in an hourly resolution. Different agent subgroups share a certain schedule type, i.e. their schedules comprise the same location types, but differ in the exact location and the exact times of movement. These subgroups are underage, working adult, teacher, medical professional, public worker and pensioner and reflect the occupation of an agent. Each subgroup consists of a fraction of people within a specific age section (0-17: underage; 18-66: working adults, teachers, medical professionals, public workers; 67+: pensioners). The schedules are designed in a way that every agent spends some time at its home (typically the nights). During the day, underaged visit schools and (working) adults visit workplaces. Teachers, medical professionals or public workers, visit schools, hospitals, and public places in a comparable regular fashion, respectively. All agents occasionally visit public places during weekdays and more extensively on weekends.

In addition to the regular schedule, all agents share the so-called specific schedules that determine their (restricted) movements under defined circumstances, in the model usually represented by defined health (sub-)states, such as a diagnosed schedule, a hospital schedule, and a death schedule (see Figure S3). These schedules constantly assign the corresponding agent to a single location, i.e. either the agent's home, a hospital or the morgue, as long as the agent's state is $I^d$ (diagnosed), $I^d_H$ (hospitalized), $I^d_{ICU}$ (in ICU) or D (dead), respectively. The only exception to this are non-compliant agents, who refuse to stay at home when they are diagnosed ($I^d$), but keep on following their regular schedule instead of a diagnosed schedule. Examples for schedules are given in section 3.3.2.

During a simulation, one schedule, by default the regular schedule, is set to active. The agents follow their active weekly schedules in the exact same way every week. The active schedule is changed to a specific schedule when an agent changes its health state. When non-pharmaceutical measures (e.g. lockdown) are requested (for all agents or a subgroup of agents), the schedules get modified accordingly for the duration of the implemented measures, i.e. such that agents stay in their respective homes, instead of visiting specified location types.

The schedules are the basis of the agents' interaction network. It is important to note that while agents visit the same locations at the same time each week, they possibly interact with different other agents on each visit. The interaction network is the basis for potential infection transmission resulting in the agent's health state transitions over time. As stated above, health state transitions can feed back into schedule activation (principles illustrated in **Figure S2**).

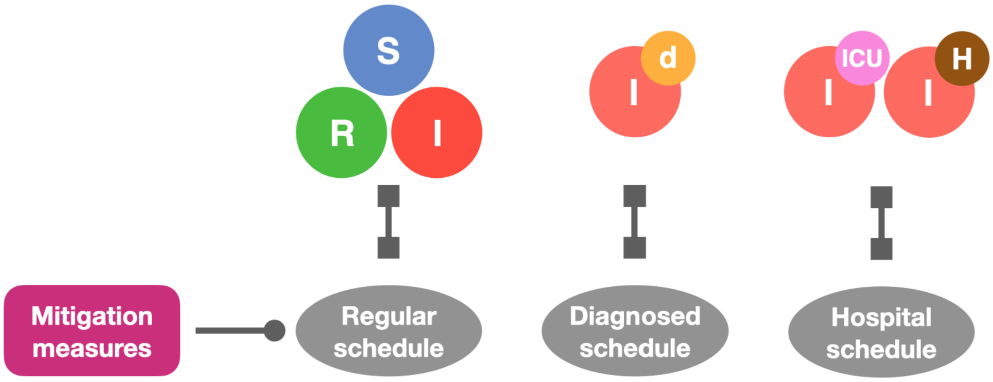


**Figure S2 - Schedule assignment and modification.** Agents have an active schedule at any time. By default, this is the regular schedule, which can be changed upon health state transition or application of non-pharmaceutical interventions. While regular schedules differ between agents, diagnosed, hospitalized, and dead schedules are specific schedules that place an agent 24/7 at home, in the hospital, or on the morgue, respectively. For non-compliant agents, the mitigation or lockdown schedules can be set equal to their regular schedule.

##### 2.2.3 Motion Between Locations According to Schedules

During a simulation, each agent sequentially visits different locations, defined by its schedule. The resolution for location changes is per hour, in consistency with (*29*) which showed that 70% of the social contacts last an hour or more . Several agents can visit the same location at the same time (**Figure S3**). This spatio-temporal overlap of agents creates their network of possible interactions and it further facilitates the network for infection spreading.

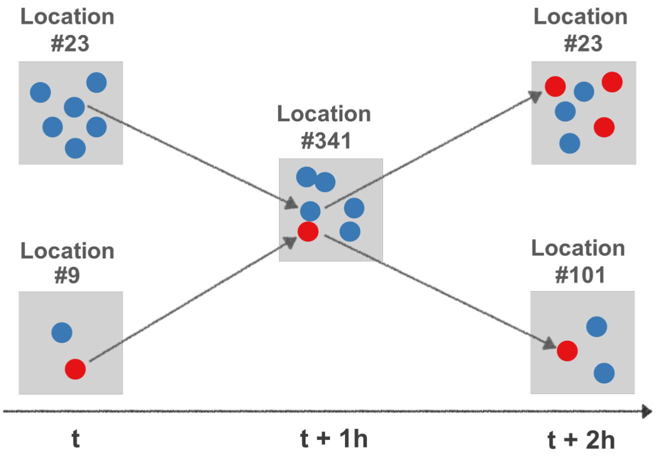


**Figure S3: Schematic representation of agents visiting locations at different times *t*.** At time *t*, six susceptible agents are at location #23 and one susceptible and one infected at location #9. In the next time step (*t+1*), one susceptible agent from location#23 and the infected agent from location #9 move to location #341. Here, they interact and transmit the infection. At time *t*+2, the newly infected agent moves back to location #23, while the first infected agent continues to location #101. Two other infected agents have moved to location #23 between times *t* and *t*+2.

##### 2.2.4 Agent-agent Interactions

During a simulation, all agents with spatio-temporal overlap, i.e. being at the same location at the same time step, have the possibility to interact with each other. Interacting pairs are determined, regardless of their health state, by a stochastic process. Each of the $n_{loc}$ agents in a location loc at time *t* has the possibility to interact with each of the $n_{loc}$-1 other agents with a given probability. We define a global interaction frequency $\mu$. Due to the stochastic nature of the interaction mechanism, some agents have more than the expected number of interactions while others do not have any.

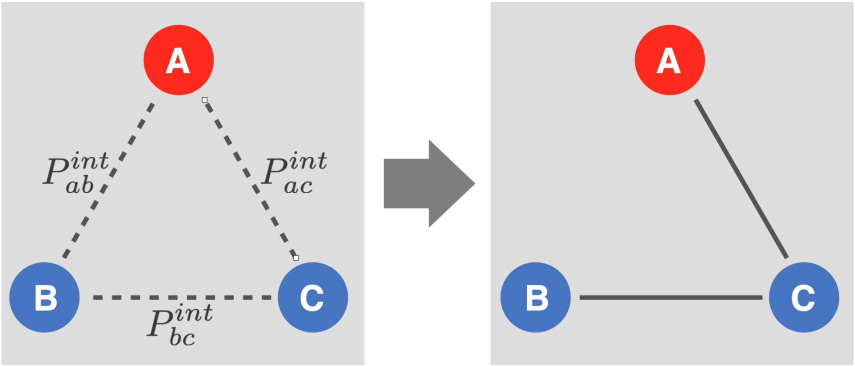


**Figure S4: Schematic representation of agent-agent interactions.** Left: Each agent has a certain probability to interact with each other agent having spatio-temporal overlap. Right: realized interactions.

The probability for any agent $a$ to interact with any other agent $b$ in the same location is dependent on their specific interactivities $x_a$ and $x_b$, the global interaction parameter $\mu$ and the number of agents per location $n_{loc} = n_{loc}(t)$ (where $n_{loc} \geq 2$ in order to enable an interaction).

Equation (1) defines the interaction probability $P_{ab}^{int}$ of two agents $a$ and $b$ per time step (at specific location):

$$P_{ab}^{int} = \frac{Minimum(n_{loc} - 1, x_a x_b \mu)}{n_{loc} - 1}$$

Equation (1)

This interaction probability is calculated for all potential pairs of agents present at the same time at the same location. The interaction is realized if a random number between 0 and 1 is smaller than the interaction probability.

Specific interactivities of agents,$x_a$, are set to 1 as default value, but can be set differently to reflect specific individual behavior (e.g. socially active or shy individuals).

The global interaction frequency $\mu$ is the expected number of interactions per time step for sufficiently occupied locations and $x_a = x_b = 1$. The realized number of interactions depends on the number of agents currently present at the specified location. Since an agent can maximally interact with $n_{loc}$- 1 other agents at any given time, the real mean of interactions may be lower than $\mu$, if $n_{loc}$- 1 is smaller than $\mu$.

#### 2.2.5 Infection Transmission

$P_{ab}^{inf}(t, loc)$is the probability for the infection transmission from agent $a$ to agent $b$ at location $loc$, where $t$ denotes the infection duration of agent $a$ in hours.

Given an interaction occurred between an infected agent $a$ and a susceptible agent $b$, the conditional probability for infection transmission is defined as:

$$p_{ab}^{inf} = p_{ab}^{inf}(t, loc) = Minimum\big(1, k_I \cdot I_a^I(t)\big)$$

Equation (2)

with $I_a^I(t)$ being the infectiousness of the infected agent and $k_I$ a globally fitted infection parameter. The approach can be extended such that parameter $k_I$is

$$k_I = k_I\big(y_a^{inf}, y_b^{inf}, f^{loc}\big)$$

Equation (3)

where $f^{loc}$ is a location-specific factor representing the infection risk at different types of locations (offices, hospital, outside areas) and $y^{inf}$ presents specific coefficients for the interacting agents modulating the transmission probability (e.g. representing protective gear).

Hence, the probability for the infection to get transmitted from agent $a$ to agent $b$ at location $loc$ and at time $t$ is the product of the conditional probability for the infection transmission and the probability for an interaction to occur,

$$P_{ab}^{inf} = p_{ab}^{inf} \cdot P_{ab}^{int}.$$

Equation (4)

Values for the infection-duration dependent infectiousness $I_a^I(t)$ are given in section 3.5 and **Table S26 (E)**.

#### 2.2.6 Health States

As mentioned above, agents can assume different health states, namely susceptible (S), infected (I), recovered (R), and dead (D) with the default health state S. In the previous section, we described the infection transmission, i.e. the transition from state susceptible to state infected (see **Figure S1**). Here, we provide more details about the infected substates and about transitions between all health states.

The four major health states (S,I,R,D) specify the epidemiological conditions of the agents. They are the basis for simulating virus infection progression in the modeled population and to analyze transmission chains on individual level. In order to cover disease progression for individual agents more precisely, the infected state is divided into four sub-states that can be associated with severity of progression and are described in detail below. In summary, this gives us seven distinguishable health states (see **Figure S1**), of which the four representing different infected states are transient, i.e., no agent can remain infected infinitely. Agent behavior is influenced by the health state: Being susceptible or infected without diagnosis, an agent follows its regular schedule while upon infection diagnosis the diagnosis schedule gets activated.

Susceptible agents have never been subject to the infection and neither are immune to it. They can become infected based on an interaction with an infected agent. They follow their regular schedule.

Recovered agents have survived an (either asymptomatic or symptomatic) infection, are not infectious anymore, and are considered immune. The model allows for loss of immunity by a transition from state R to state S, in order to account for the latest discussions about potential SARS-Cov2 antibody loss. However, this transition is disabled in the presented study (by parameterization with zero), due to lack of sufficient evidence.

Infected agents comprise presymptomatic, asymptomatic, and symptomatic infected individuals. In order to specify and distinguish between different types of severity of the infection, various attributes are associated with the different substates of the infection. Those attributes affect agent behavior, e.g. by activating the diagnosed schedule, and ultimately reflect the information available during a real-world pandemic. The plain infected state (I) represents the unrecognized cases, i.e. the vast majority of presymptomatic or asymptomatic cases. The diagnosed state ($I^d$) represents the recognized infections including cases with or without symptoms, irrespective of how they were identified. These diagnosed cases may escalate towards two subsequent stages of severity. These are either hospitalization on the normal ward for cases of moderate severity ($I^d_H$) or the treatment in an intensive care unit (ICU) for the critically severe cases ($I^d_{ICU}$).

Due to parameterization (as explained in section 3.4), all infected states are transient.

Infected individuals have a probability to either recover or to die. Birth and death processes of the susceptible or recovered states are neglected in the current version of the model.

Dead agents are assigned to the morgue.

Throughout the manuscript and simulations, we use the following color code: susceptible – blue, recovered – green, infected – red, dead – black. In addition, for the subgroups of infected holds: diagnosed – orange, hospitalized – brown, in ICU – magenta (compare **Figure S1**).

Formally, we will refer to the states $S_i,\ i = 1,..,7$. The assignment of states to numbers is given in **Table S2**.

| **State** | S | I | $I^d$ | $I^d_H$ | $I^d_{ICU}$ | R | D |
|---|---|---|---|---|---|---|---|
| **Symbol** | $S_1$ | $S_2$ | $S_3$ | $S_4$ | $S_5$ | $S_6$ | $S_7$ |

**Table S2**: Numbering of health states for formal analyses.

#### 2.2.7 Health State Transitions

At each time step, an infected agent can either remain in its current state, progress to another infected state, recover, or die (**Figure S1** and **Table S3**). This is determined by a discrete stochastic decision process, based on the respective probabilities given in Tables **S26 (A-E)**, which is detailed in section 3.4.

| | $S_1$ | $S_2$ | $S_3$ | $S_4$ | $S_5$ | $S_6$ | $S_7$ |
|---|---|---|---|---|---|---|---|
| $S_1$ | $X_{11}$ | $X_{12}$ | 0 | 0 | 0 | 0 | 0 |
| $S_2$ | 0 | $X_{22}$ | $X_{23}$ | 0 | 0 | $X_{26}$ | $X_{27}$ |
| $S_3$ | 0 | 0 | $X_{33}$ | $X_{34}$ | 0 | $X_{36}$ | $X_{37}$ |
| $S_4$ | 0 | 0 | 0 | $X_{44}$ | $X_{45}$ | $X_{46}$ | $X_{47}$ |
| $S_5$ | 0 | 0 | 0 | 0 | $X_{55}$ | $X_{56}$ | $X_{57}$ |
| $S_6$ | $X_{61}$ | 0 | 0 | 0 | 0 | $X_{66}$ | 0 |
| $S_7$ | 0 | 0 | 0 | 0 | 0 | 0 | $X_{77}$ |

**Table S3:** Possible Transitions $X_{ij}$ in agreement with the network shown in **Figure S1**.

Formally, we will refer to the allowed state transitions by $X_{ij}$ with $i, j = 1, .., 7,$ as shown in **Table S3**. In order to determine whether a health state transition occurs, we used the following probabilities:

$P_{iT}^{S}$ is the probability for being in state $S_i$ at time $T$.

$P_{ij}^{c}$ is the cumulative probability for transition from $S_i$ to $S_j$ (total probability).

$P_{ijt}^{X}$ is the probability for transition from $S_i$ to $S_j$ (per time step).

$p_{ijt}^{X}$ is the conditional probability for transition from $S_i$ to $S_j$ (per time step).

Together with the transition from susceptible to infected, we consider in total 13 potential transitions in the model, as listed below together with the respective probabilities:

| State Transition | Symbol | transition probability |
|---|---|---|
| S → I | $X_{12}$ | $p_{12t}^{X}$ |
| I → I$^{d}$ | $X_{23}$ | $p_{23t}^{X}$ |
| I → R | $X_{26}$ | $p_{26t}^{X}$ |
| I → D | $X_{27}$ | $p_{27t}^{X}$ |
| I$^{d}$ → I$^{d}_{H}$ | $X_{34}$ | $p_{34t}^{X}$ |
| I$^{d}$ → R | $X_{36}$ | $p_{36t}^{X}$ |
| I$^{d}$ → D | $X_{37}$ | $p_{37t}^{X}$ |
| I$^{d}_{H}$ → I$^{d}_{ICU}$ | $X_{45}$ | $p_{45t}^{X}$ |
| I$^{d}_{H}$ → R | $X_{46}$ | $p_{46t}^{X}$ |
| I$^{d}_{H}$ → D | $X_{47}$ | $p_{47t}^{X}$ |

| | | |
|---|---|---|
| $I^d_{ICU} \rightarrow R$ | $X_{56}$ | $p^X_{56t}$ |
| $I^d_{ICU} \rightarrow D$ | $X_{57}$ | $p^X_{57t}$ |
| R → S | $X_{61}$ | $p^X_{61t}$ |

**Table S4**: Health state transitions and the respective cumulative transition probabilities.

Considering all possible state transitions $X_{ij}$to change from state $S_i$ to $S_j$,the probability for each transition then is given by:

$$P^X_{ijT} = P(X_{ij}, T) = P(X_{ij}, T \mid X_{ii}, t = 1, \ldots, T-1) \cdot P(X_{ii}, t = 1, \ldots, T-1),$$

Equation (5)

where the probability ($P^S_{iT}$) to remain in the state $S_i$ until time *T* is one minus the sum of the probabilities to leave the state before *T*:

$$P^S_{iT} = P(X_{ii}, t = 1, \ldots, T-1) = 1 - \sum_{t=1}^{T-1} \sum_{j=1}^{n_S} P^X_{ijt}.$$

Equation (6)

We now can rewrite the Equation (5) for the state transition at time $T$ to:

$$P^X_{ijT} = p^X_{ijT} \cdot P^S_{iT}.$$

Equation (7)

Hence the required conditional probability for a state transition at each time step used in the GERDA-model follows as .

$$p^X_{ijT} = \frac{P^X_{ijT}}{P^S_{iT}} = \frac{P^X_{ijT}}{1 - \sum_{t=1}^{T-1} \sum_{j=1}^{n_S} P^X_{ijt}},$$

Equation (8)

which is the required conditional probability for a state transition at each time step used in the GERDA-model.

The derivation of numerical values for the transition probabilities from published data is detailed in Sec 3.4.2.

### 3 Model Initialization and Data Integration

The model is initialized in three steps:

(i) Locations are initialized either from real geographical data (default) or arbitrarily (for alternative scenarios or as randomized comparison). The location type home defines the number of households.

(ii) Households are initialized based on data of German zensus assigning household types to all home locations (1:1 ratio) and defining a respective number of household members (1:n ratio). Households are only relevant for initialization; they are not stored as simulation objects.

(iii) Agents are initialized according to households and with a number of attributes that either depend on their household membership or on additional data. Agents obtain an occupation and a weekly schedule that is related to their occupation.

Underlying data and details are explained below. Details of code are described in GitLab.

#### 3.1 Location Initialization

##### 3.1.1 Principles of Location Initialization

The initial step in GERDA is the generation of an *in silico* representation of the real-world community of interest through the creation of distinct locations. These locations define the space in which the agents can interact, and, in addition, the size of the agent population as described in Sec. 2.2 and further explained below in Sec. 3.2.

In general, locations are initialized from an input data file (see Sec. 3.1.2) specifying the floor area, the geographical coordinates and, if appropriate, the neighborhood identifier of that location. The input data contains further annotations of each building in one of the following annotation categories: building, amenity, shop, leisure, sport or healthcare. According to these annotations, each building is assigned a specific location type as introduced in Sec. 2.1. For the municipalities Heinsberg and Gangelt an assignment table (**Table S5**) was created which ought to be extended, when including further geolocations.

If a building is not annotated otherwise, its location type is set to home during initialization. Further if any of the two required locations, morgue or hospital, are not assigned to at least one location, they are added as an initialized location at the border of the generated world. Alternatively to the described procedure, one location called mixing location is initialized to allow for comparison with non-localized agent based models.

| location type | resp. specific annotations | annotation category |
|---|---|---|
| workplace | industria, greenhouse, cowshed, shed, commercial, warehouse, office, farm, fire station, farm auxiliary, retail | |
| public place | public, chapel, church, parish hall, townhall, restaurant, grocery store, cafe, sports centre | leisure, sport |
| school | school, university, kindergarten | |
| hospital | | healthcare |
| morgue | - | - |
| home | residential building | |
| excluded building | garage, roof, shed, bungalow, silo, barn | |

**Table S5. Location type assignment.** All initialized buildings are assigned one location type, if applicable, according to the distinct annotation category and, otherwise, according to a specific annotation.

##### 3.1.2 Data used for Location Initialization

To generate the geospatial maps, we utilized OpenStreetMap (OSM), an editable map database built and maintained by volunteers, distributed under the Open Data Commons Open Database License, and available from https://www.openstreetmap.org. OSM data retrieval is facilitated by the OSMnx package for python (*30*) and was integrated generically. Using the OSM addresses of sufficiently annotated administrative regions, such as the community Gangelt, data on area, street network, and buildings is downloaded. Area and street network data is used for visualization purposes only. All retrieved buildings of the community of interest, whose floor area is above a predefined threshold, are considered as locations in the model. Thresholding is necessary since not all buildings in the OSM database are sufficiently large to be considered a location with respect to our model categories. The building data is cropped to those relevant to determine the building functionality and extended by a manually curated neighborhood identifier to subdivide locations in functional clusters.

#### 3.2 Household Initialization

##### 3.2.1 Principles of Household/Population Initialization

To create a realistic population, the agents of a MW are initialized based on the concept of households. By iterating over all locations of type home and assigning them a household type and size from a data-based distribution, a respective number of agents is initialized. Each agent belongs to a unique household. This way, we create a reasonable demographic as well as a family structure of the agent population. Several agent properties depend directly on the agent's household, while further properties are added consecutively. The data that was used for agent initialization and the dependencies of agent properties on that data and on each other are detailed in the following subsections.

##### 3.2.2 Data for Household Initialization

For the definition of households, we used data from the German Zensus 2011 database that is available online (https://ergebnisse.zensus2011.de). The data can be obtained either in form of predefined tables (https://ergebnisse.zensus2011.de/#StaticContent:00,,,) or by specifying the combination of desired information in the dynamic table section (https://ergebnisse.zensus2011.de/#dynTable:). If not specified otherwise, the tables in this subsection were predefined. To use the latest age distribution, the age distribution for the German population of 2020 was integrated from (https://service.destatis.de/bevoelkerungspyramide). All data described in this subsection has been released by the German government and details on data collection and processing are described at the given webpage for each table. **Tables S6 to S12** listed below are available in our separate **Supplementary Tables** file.

- **Table S6: Household type frequencies.** Absolute and relative frequencies of household/family types in Germany. The table content has been translated to English, original German content is kept in brackets and small font to allow comparison with the original source.

  https://ergebnisse.zensus2011.de/#StaticContent:00,BEG_3_1,m,table

- **Table S7: Absolute distribution of households according to type and size.** Original information obtained from German Federal Government by using the dynamic table section of the Zensus 2011 database. The table content has been translated to English, original German content is kept in brackets and small font to allow comparison with the original source.

  https://ergebnisse.zensus2011.de/#dynTable:statUnit=HAUSHALT;absRel=ANZAHL;ags=00;agsAxis=X;xAxis=HHGROESS_KLASS;yAxis=HHTYP_FAM

- **Table S8: Age distributions per household type.** Total number of people in each age range (11 groups) per household type. The table content has been translated to English, original German content is kept in brackets and small font to allow comparison with the original source. As there was no specific information provided concerning the underage members of couples without kids and they only compose a very minor fraction of the population (<0.1%), we omitted the numbers grayed out.

  https://ergebnisse.zensus2011.de/#StaticContent:00,BEV_10_25,m,table

- **Table S9: Current age distribution in Germany.** This table was extracted from the 'Bevölkerungspyramide' (i.e. 'age pyramide') and shows the complete age distribution of the German population for 2020.

  https://service.destatis.de/bevoelkerungspyramide/#!y=2020

According to our concept to initialize the modeled population based on available home locations, the household data used is based on occupied flats/houses and not on families. The GERDA model in its current version allows five household types: Single person household, couple without kids, couple with kid(s), single parent household, and multi-person household without nuclear family. Household sizes range from one to six agents, with bigger households being included in size category '6+' in the data. For sake of simplicity, a few assumptions have been made when integrating the data. Household types that compose a minor fraction of the population have been included within the five types introduced above. E.g. couples without kids can only have a household size of two, thereby omitting specific households like grandparents with children. The omitted special cases as well as multi-generation households are included in the household category multi-person households without nuclear family. An algorithm was created to combine the above described data into probability distributions from which the number of agents per household and ages of the resulting household members are drawn. It is outlined below. The distributions used for GERDA simulation are listed below:

- **Table S10: Relative household type distribution.** Calculated from Table S6.
- **Table S11: Relative household size distribution.** Household-type-dependent; calculated from Table S7. For simplicity in implementing household initialization, the household type 'couples without kids' can only be of size 2, thereby ignoring very specific family types, such as grandparents with a kid. Those ignored family types compose about 1.6% of the total population. Used as input for GERDA simulation.
- **Table S12: Relative age distribution per household type.** Calculated from Tables S8 and S9.

###### 3.2.2.1 Algorithm

For household initialization, the probabilistic relative distributions obtained from the tables introduced above are used in the following order: First, the household type is sampled (**Table S10**). Second, the household size is sampled in a household-type dependent manner (**Table S11**). Finally, the age(s) of the agents living in the household (number of agents = household size) are sampled. This last step is achieved using a probabilistic distribution based on mapping the real distribution of ages in Germany 2020 (**Table S9**) as weights to the household type age distribution (**Table S8**). This results in the calculation of the relative age-dependent frequencies for each household-type (0-99 years) (**Table S12**).

As a result, agents are initialized with a unique ID, a home location, and an age. Age assignment of all agents of a household is accomplished in a household type-dependent manner: i) If the household contains kids (couple with kids or single parent household), the algorithm starts sampling the kid(s) (number of kids = n-2 or n-1, respectively), and then the parent(s) age(s) are sampled from a shifted normal distribution depending on the age of the eldest kid. The age-offset for kids, i.e. their maximum age, is set to 30 years. The sampling of sibling(s) is made by a shifted normal distribution, assuming on average an age difference of 3 years and a standard deviation of 1.5 years. To avoid negative ages, absolute values are taken. ii) In case of a couple without kids, the first member is sampled and the generated age of this member is used to make a normal distribution with a standard deviation of 1/10 of this age. The age of the second member is obtained based on the aforementioned normal distribution. iii) In the rest of the cases the age is obtained by the corresponding probability distribution of ages per household type.

The output of the algorithm provides a household with type, size, and ages of the respective number of household members for each home location.

#### 3.3 Agent Initialization

##### 3.3.1 Principles of Agent Initialization

For each member of a household, an agent is initialized with a unique ID, the household's location as home location, a household-dependent age, and, by default, the health state susceptible. Based on its age a schedule is created for this agent that also defines the occupation. This schedule determines the agent's movement during a simulation. The details about schedule creation during model initialization are described in the following subsection.

The subsequent subsection outlines the optional possibility to set an agent's health state to infected upon initialization.

##### 3.3.2 Schedule Generation for Agents

Schedules are created individually for each agent during agent initialization. The age and the home location of an agent are required to create its regular schedule. To this end, an input table that defines the possible schedule types has to be provided. Conceptually, there is no limit to the number of possible schedules that can be defined in the input table. Instructions on how such a table can be set up are given in the GitLab repository (https://tbp-klipp.science/GERDA-model/). The predefined schedules are specific for different age categories, such that typical occupations for people of different ages can be accounted for. Within each age category, a set of possible schedules and their proportion are defined. The proportion reflects the fraction of people in the corresponding age category that should follow the respective schedule. Thereby, each schedule belongs to a type that is defined in the input table. This type does not have to be exclusive, so there can be many schedules of type medical professionals, for example, reflecting agents with the same occupation, but different shifts or free time habits.

For each agent, the regular schedule is drawn from the set of schedules within its age category. The mentioned proportions serve as probabilities for each schedule to be drawn. By definition, a schedule covers 7 days (168 hours), to allow for differences in agent movements and localizations between different days, e.g. during work days and weekends. The schedules contain a list of times (hours of the week) or time spans with a resolution of one hour and a list of location types. The times determine the hours of the week at which the agent moves from its current location to the next. If a time span is provided, the exact time for the movement is drawn from a uniform distribution within the time span. When the exact times are defined, the agent gets assigned a set of explicit locations. For each location type in the schedule an explicit location of the corresponding type is drawn with a probability linearly decreasing with their relative distance to the agent's home location. Relative distance means that all existing locations of this type are ranked by their distance to the home location, such that only the order of distances and not the real geographical distance is of importance. If there are several locations of the same type in a schedule, they are denoted with a number (e.g. public 1, public 2). For each number, a different location of this type is drawn that stays associated with this number for this agent. For example, public 1 is always the same location of type public whenever it appears in the agent's schedule. Now all the exact times and locations are set for this agent, representing its regular schedule.

During the simulation, the agent can, instead of the regular schedule, get assigned a diagnosed or hospitalized schedule, respectively. The diagnosed schedule reflects quarantine rules, i.e. the agent has to stay at home. In case of non-compliance, the agent continues to follow its regular schedule. With a hospitalized schedule, the agent stays in the hospital.

Each agent has an active schedule at each time that specifies its movements between locations over time. **Table S13** shows three typical regular schedules of different types and one diagnosed schedule. **Figure S5** shows a snapshot of the complete input table for schedules to be drawn from, which is available in the additional file for **Supplementary Tables, Table S14**. For detailed information about creation of schedules, please refer to https://tbp-klipp.science/GERDA-model/schedule.

| **schedule type** | **pensioner** | | **working adult** | | **underage** | | **diagnosed** | |
|---|---|---|---|---|---|---|---|---|
| | **time, h ( week)** | **location** | **time** | **location** | **time** | **location** | **time** | **location** |
| | 0 | home | 0 | home | 0 | home | 0 | home |
| | 10 | public 1 | 8 | work | 7 | school | 9 | home |
| | 11 | home | 16 | public 1 | 15 | public 1 | 14 | home |
| | 36 | public 2 | 18 | home | 18 | home | 19 | home |
| | 37 | home | 32 | work | 32 | school | 33 | home |
| | 55 | public 3 | 40 | public 2 | 39 | public 2 | 38 | home |
| | 56 | home | 42 | home | 44 | home | 43 | home |
| | 82 | public 1 | 56 | work | 56 | school | 57 | home |
| | 85 | home | 64 | public 3 | 62 | public 3 | 62 | home |
| | 112 | public 3 | 65 | home | 68 | home | 67 | home |
| | 113 | home | 80 | work | 80 | school | 81 | home |
| | 127 | public 2 | 88 | public 1 | 85 | public 1 | 86 | home |
| | 128 | home | 89 | home | 93 | home | 91 | home |
| | 156 | public 4 | 104 | work | 104 | school | 105 | home |
| | 157 | home | 112 | public 3 | 110 | public 3 | 110 | home |
| | | | 114 | home | 115 | home | 115 | home |
| | | | 133 | public 4 | 132 | public 4 | 129 | home |
| | | | 138 | home | 138 | home | 134 | home |

| | | | | | | | | | | |
|---|---|---|---|---|---|---|---|---|---|---|
| | | | 154 | public 5 | | 155 | public 5 | | 139 | home |
| | | | 163 | home | | 162 | home | | 153 | home |
| | | | | | | | | | 158 | home |
| | | | | | | | | | 163 | home |

**Table S13. Example schedules.** Agent-specific schedules with fixed hours, possibly ranging from 0 to 167 hours (over the course of a week) defining the agents' movements during a simulation.

| age (upper bound) | version | fraction | description | Start time | expand to range | **mo** | Start time | expand to range | **tu** | Start time | expand to range | **we** | Start time | expand to range | **th** |
|---|---|---|---|---|---|---|---|---|---|---|---|---|---|---|---|
| 18 | 1 | 1 | underage | 7 | 9 | school | 7 | 9 | school | 7 | 9 | school | 7 | 9 | school |
| | | | | 13 | 16 | public_1 | 13 | 16 | public_2 | 13 | 16 | public_3 | 13 | 16 | public_1 |
| | | | | 17 | 22 | home | 17 | 22 | home | 17 | 22 | home | 17 | 22 | home |
| 67 | 1 | 0.77 | adult | 7 | 12 | work | mo | | work | mo | | work | mo | | work |
| | | | | p7 | 9 | public_1 | mo | | public_2 | mo | | public_3 | mo | | public_1 |
| | | | | p1 | 3 | home | p1 | 3 | home | p1 | 3 | home | p1 | 3 | home |
| | 2 | 0.004 | medical_professional | 7 | 12 | hospital | mo | | hospital | mo | | hospital | mo | | hospital |
| | | | | p7 | 9 | public_1 | mo | | public_2 | mo | | public_3 | mo | | public_1 |
| | | | | p1 | 3 | home | p1 | 3 | home | p1 | 3 | home | p1 | 3 | home |
| | 3 | 0.003 | medical_professional | 10 | 16 | public_4 | 10 | 16 | public_5 | 7 | 12 | hospital | we | | hospital |
| | | | | 17 | 22 | home | 17 | 22 | home | p7 | 9 | public_3 | we | | public_1 |
| | | | | | | | | | | p1 | 3 | home | p1 | 3 | home |
| | 4 | 0.0015 | medical_professional | 6 | 7 | home | mo | | home | mo | | home | mo | | home |
| | | | | p12 | 14 | public_1 | p12 | 14 | public_2 | p12 | 14 | public_3 | p12 | 14 | public_1 |
| | | | | 22 | 23 | hospital | mo | | hospital | mo | | hospital | mo | | hospital |
| | 5 | 0.0015 | medical_professional | 6 | 7 | home | 10 | 16 | public_5 | mo | | home | mo | | home |
| | | | | p12 | 14 | public_4 | 17 | 22 | home | p12 | 14 | public_1 | p12 | 14 | public_2 |
| | | | | p2 | 3 | home | | | | 22 | 23 | hospital | we | | hospital |
| | 6 | 0.02 | teacher | 7 | 9 | school | 7 | 9 | school | 7 | 9 | school | 7 | 9 | school |
| | | | | 13 | 15 | public_1 | 13 | 15 | public_2 | 13 | 15 | public_3 | 13 | 15 | public_1 |
| | | | | 16 | 18 | home | 16 | 18 | home | 16 | 18 | home | 16 | 18 | home |
| | 7 | 0.1 | public_worker | 7 | 9 | public_1 | mo | | public_1 | mo | | public_1 | mo | | public_1 |
| | | | | p6 | 8 | public_2 | mo | | public_3 | mo | | public_4 | mo | | public_2 |
| | | | | p1 | 3 | home | p1 | 3 | home | p1 | 3 | home | p1 | 3 | home |
| | 8 | 0.1 | public_worker | 11 | 13 | public_2 | mo | | public_3 | mo | | public_2 | mo | | public_4 |
| | | | | p1 | 3 | public_1 | mo | | public_1 | mo | | public_1 | mo | | public_1 |
| | | | | p6 | 8 | home | p1 | 3 | home | p1 | 3 | home | p1 | 3 | home |

**Figure S5: Snapshot of the schedule input table.** The possible schedules are defined by an input table that determines age categories and different schedule versions within each category that reflect typical schedules for different occupations. The corresponding complete Table S14 is found in the additional Supplementary Tables file, detailed instructions on schedule creation are found at https://tbp-klipp.science/GERDA-model/schedule.

#### 3.4 Health State Transitions

##### 3.4.1 Principles

As introduced in section 2.2.6, in the GERDA-model agents can assume four main different health states: susceptible S, infected I, recovered R or dead D (**Figure S1**, **Table S2**). Once infected, an agent can successively assume four different infection sub-states: The (a-/presymptomatic) infected I can possibly become diagnosed $I^d$, subsequently hospitalized $I^d_H$, and further referred to ICU $I^d_{ICU}$. Alternatively, the infected agent - generally from all of its different sub-states - can recover (transition to state R) or die (transition to state D) (**Table S3**).

Because all state transitions are irreversible (compare unidirectional arrows in **Figure S1**), this leads to a total number of thirteen possible health state transitions. However, due to the lack of sufficient data on reinfection with SARS-CoV-2 when already having recovered from it, represented by state transition $X_{61}$, loss of immunity is not considered in the presented GERDA model. This is technically achieved by setting the respective cumulative state transition probability $P_{61}^{c}$ to zero. Furthermore, we exclude cases of dying from COVID-19 without being recognized, assuming that any deceased gets tested and a positively tested deceased is counted as COVID-19-related death. The cumulative probability $P_{72}^{c}$ of the corresponding state transition $X_{72}$ is set to zero. Transitions with probability zero are depicted by dotted arrows in **Figure S1** and omitted in the tables of the following subsections.

For the remaining eleven state transitions, simulation of GERDA requires not only cumulative state transition probabilities $P_{ij}^{c}$ (i.e. how likely is it that a diagnosed individual has to got to hospital), but hourly resolved conditional transition probabilities $p_{ijt}^{X}$ for transition from state $i$ to state $j$ at time $t$. These probabilities depend on the duration of the infection and the time being in the current state, respectively. These probabilities also depend on the age of the agent. The following subsections outline how data from different sources have been combined, complemented with educated guesses, and processed to calculate those conditional hourly probabilities $p_{ijt}^{X}$ for all state transitions and different age groups.

##### 3.4.2 Input Data for Health State Transition Rates

If referenced by RKI Factsheet, we refer to the weekly status report of the Robert-Koch-Institute (RKI) from the 14th of March 2020 (*31*). From this factsheet, we extracted information related to COVID-19 provided in **Tables S15, S16, S17** given in the separate file **Supplementary Tables**:

- **Table S15: Reported COVID-19 cases.** Source: RKI Factsheet (*31*). 201 cases have been omitted due to lacking age information. For the remaining cases we calculated the total number (age group 'all').
- **Table S16: Deceased COVID-19 cases.** Source: RKI Factsheet (*31*). Six cases were omitted due to incomplete information. For the rest we calculated the gender-independent numbers (column 'total') and the overall cases (age group 'all').
- **Table S17: Hospitalization information for COVID-19 cases.** Source: RKI Factsheet (*31*).

Age distributions not included in the RKI Factsheet have been taken from (*32*).

- **Table S18: Age group dependent information on hospitalization and referral to ICU.** Source: (*32*).

German Census data is used to determine the demography of our susceptible model population (state $S_1$) and, due to lack of age-dependent data on infection probability, also for infected agents (state $S_2$).

- **Table S19: Demography Germany 2011.** Source: German Zensus 2011 database.

To estimate the diagnosis probability ($P_{23}^{c}$), we used the homepage of the municipality Gangelt (https://www.gangelt.de/news/226-erster-corona-fall-in-nrw) and an early report about COVID-19 dynamics in Gangelt from (*33*). Gangelt, a small town in the district of Heinsberg, was one of the first 'Corona hotspots' in Germany, where a lot of residents got infected during a large carnival event ("Kappensitzung", Feb 15, 2020). Being of high interest for the understanding of SARS-CoV-2 and COVID-19, the entire population of Gangelt got tested, giving rise to the usually unknown dark number of infected individuals and, subsequently, the total diagnosis probability including not only symptomatic but also cases diagnosed based on contact information.

The usage of the above introduced data, the required assumptions, and the calculations applied to ultimately obtain age-dependent conditional probabilities $p_{ijt}^{X}$ for all possible state transitions $X_{ij}$ between states $S_i$ and $S_j$ per time step $t$ are outlined in the following subsections.

#### 3.4.3 Probability Calculations

##### 3.4.3.1 Age-independent state transition probabilities

In section 2.2.7, we explained how to calculate cumulative and time-dependent conditional probabilities for the transitions between different states. Here and below, we outline how to convert input data with different levels of detail into the input information for the state transition probabilities required for GERDA simulation.

First, we extracted the age-independent cumulative state transition probability $P_{ij}^{c}$ from the data listed in section 3.4.2. For the cases released from ICU, we know exactly how many have been discharged alive and how many dead, resulting in $P_{56}^{c}$ and $P_{57}^{c}$. For cases with hospitalization information, we know how many are hospitalized (resulting in $P_{34}^{c}$) and how many of them are referred to ICU (resulting in $P_{45}^{c}$). For specific transitions, namely, we can derive the probabilities directly from the available data as follows.

Since we know how many people died out of all reported cases (see **Table S16**) and how many are released dead from ICU ($I^{d}_{ICU}$→D, $X_{57}$) we can infer the total number of people that died either in hospital ($I^{d}_{H}$→D,$X_{47}$) or somewhere else ($I^{d}$→D,$X_{37}$). However, we do not know the exact numbers for these two state transitions separately. We assume that people with more severe symptoms are people with a more severe disease progression. Those people are more likely to be referred to hospital, and more likely to die. Consequently, it should be more likely to die from state $I^{d}_{H}$ than from state $I^{d}$. For lack of other information, we assumed that 90% of the deaths not occuring in ICU occur in hospitals. For the given numbers, this results in 1657 people who died in hospital but not in ICU and 184 people who died elsewhere. The number of reported cases serves as the reference group for the 184 people who died elsewhere and by knowing the probability to get hospitalized, we can calculate the reference group for the 1657 people who died in hospital, i.e. the number of people referred to hospital assuming hospitalization information for all reported cases. This allows to calculate the respective cumulative state transition probabilities $P_{37}^{c}$ and $P_{47}^{c}$.

The probability to get diagnosed ($P_{23}^{c}$) was inferred from two different sources, a report from (*33*) and the Gangelt homepage (https://www.gangelt.de/news/226- erster-corona-fall-in-nrw). Assuming further that nobody remains in one of the different infected substates but that everybody eventually leaves this state, we could infer the remaining cumulative state transition probabilities $P_{26}^{c}$, $P_{36}^{c}$, and $P_{46}^{c}$, i.e., the probabilities to recover from states $S_2$ (I), $S_3$ ($I^d$), and $S_4$ ($I^d{}_H$).

In contrast to the state transitions described above, which solely depend on the state the agent is in and the time spend in this state, the state transition S→I ($X_{12}$) depends on different factors namely

- the interaction with an infected agent in state I (schedules, locations)
- the infection time dependent infectivity of interaction partner in state I (simulation input parameter infectivity)

That was explained in sections 2.2.4 and 2.2.5, respectively, and results in $P_{12}^{c}$ being the product of these two probabilities (compare **Equation 8**). The determination of the infectivity is further detailed in section 3.5. The resulting cumulative state transition probabilities are summarized in **Table S20**.

| transition | symbol | value | reference | comments |
|---|---|---|---|---|
| S → I | $P_{12}^{c}$ | *variable* | - | *see section 2.2.5* |
| I → $I^d$ | $P_{23}^{c}$ | *0.3* | *(33), Gangelt homepage* | *3940566* |
| I → R | $P_{26}^{c}$ | *0.7* | - | *remaining fraction of infected* |
| I → D | $P_{27}^{c}$ | *0* | - | *COVID-19 related deaths automatically count in reported cases as well* |
| $I^d$ → $I^d{}_H$ | $P_{34}^{c}$ | *0.15* | *Table S17* | *hospitalized fraction (reference group: cases with hospitalization info)* |
| $I^d$ → R | $P_{36}^{c}$ | *0.84* | - | *remaining fraction of diagnosed* |
| $I^d$ → D | $P_{37}^{c}$ | *0.0017* | *Tables S16, S17* | *assuming 10% of deaths not in ICU from Id state* |

| | | | | |
|---|---|---|---|---|
| $I^d_H \rightarrow I^d_{ICU}$ | $P^c_{45}$ | *0.39* | *Tables S17* | *ICU fraction from hospitalized* |
| $I^d_H \rightarrow R$ | $P^c_{46}$ | *0.42* | - | *remaining fraction of hospitalized* |
| $I^d_H \rightarrow D$ | $P^c_{47}$ | *0.19* | *Tables S16, S18* | *assuming 90% of deaths not in ICU from state* $I^d_H$ |
| $I^d_{ICU} \rightarrow R$ | $P^c_{56}$ | *0.71* | *Table S18* | *fraction released alive from ICU* |
| $I^d_{ICU} \rightarrow D$ | $P^c_{57}$ | *0.29* | *Table S18* | *fraction released dead from ICU* |
| $R \rightarrow S$ | $P^c_{61}$ | *0* | - | - |

**Table S20**: State Transitions and corresponding overall (age- and time-independent) cumulative probabilities.

###### 3.4.3.2 Age-dependent state transition probabilities

To calculate age-dependent cumulative state transition probabilities $P^c_{ij}(age)$, in addition to the age-independent transition probabilities $P^c_{ij}$ listed in **Table S20**, we require age distributions for the different states $S_i$ with $i, j = 1, .., 7$. From the input data, we obtain three consensus age groups in addition to the age-independent group 'all' (Sec. 3.4.2.1):

- 0 - 59
- 60 - 79
- 80 - 99

Combining the information from German Zensus and RKI Factsheet provides the following (age-dependent) information for those consensus age groups (**Table S21**).

| **age group** | **people in Germany** | **reported cases** | **hospitalized cases** | **cases in ICU** | **cases released from ICU** | **cases released dead from ICU** | **deceased cases** |
|---|---|---|---|---|---|---|---|
| 0-59 | 58998759 | 87375 | - | - | - | - | 134 |
| 60-79 | 17007688 | 23795 | - | - | - | - | 922 |
| 80-99 | 4213248 | 11645 | - | - | - | - | 1737 |

| | | | | | | |
|---|---|---|---|---|---|---|
| all | 80219695 | 122815 | 14554 | 5700 | 3253 | 952 | 2793 |

**Table S21**: Consensus age group dependent information combined from the different data sources described above (compare Tables S15-S19). *cases* refer to COVID-19 cases.

To calculate age distributions, we first require age-resolved case numbers $N_i$ for all states $i \in \{1,\ldots,7\}$ (**Table S22**). One exception is state R ($S_6$), which emerges based on the others. As outlined above, the infection probability $P_{12}^c$ is age-independent (compare section 3.4.3.1). Therefore, the information from **Table S21** provides the age frequencies $N_i$ for the four states S ($S_1$), I ($S_2$), $I^d$ ($S_3$), and D ($S_7$). For the remaining two states $I^d_H$ ($S_4$) and $I^d_{ICU}$ ($S_5$), we used age frequencies reported for a subset of hospitalized cases (case study size n=10021) in (*32*).

| age group | num. of people in S and I | num. of people in $I^d$ | num. of people in $I^d_H$ | num. of people in $I^d_{ICU}$ | reported COVID-19 related deaths |
|---|---|---|---|---|---|
| 0-59 | 58998759 | 87375 | 2896 | 422 | 134 |
| 60-79 | 17007688 | 23795 | 3779 | 917 | 922 |
| 80-99 | 4213248 | 11645 | 3346 | 388 | 1737 |
| all | 80219695 | 122815 | 10021 | 1727 | 2793 |

**Table S22**: Age group frequencies obtained from diverse input data. Numbers of people in S and I, in $I^d$, and the reported deaths are taken from the RKI factsheet ((*31*)), numbers of people in $I^d_H$ and in $I^d_{ICU}$ from Karagiannidis et al. ((*32*)). Please note, that as the numbers are obtained from different data sources, they cannot be related directly between each other.

By dividing the number of cases in an age group $N_i(age)$ by the number of cases in age group 'all' of the same state $N_i(all)$ the age group distribution for a state *i* was calculated (**Table S23**).

| age group | fraction in S and I | fraction in $I^d$ | fraction in $I^d_H$ | fraction in $I^d_{ICU}$ | fraction in D |
|---|---|---|---|---|---|
| 0-59 | 0.74 | 0.71 | 0.29 | 0.24 | 0.05 |
| 60-79 | 0.21 | 0.19 | 0.38 | 0.53 | 0.33 |
| 80-99 | 0.05 | 0.2 | 0.33 | 0.23 | 0.62 |
| all | 1 | 1 | 1 | 1 | 1 |

**Table S23:** Age group distributions for different health states. The age group-dependent number of people in the reference state (compare Table S22) has been divided by the number for age group 'all'.

With this information and the overall cumulative state transition probabilities from **Table S20**, we calculated age group-dependent cumulative state transition probabilities $P_{ij}^{c}(age)$. Because the numbers in **Tables S22** and **S23** cannot be related between all the different states directly, we first created an example with age frequencies consistent between the different states for age group 'all'. To this end, we applied the age-independent probabilities $P_{ij}^{c}$ (compare **Table S20**) to a fictive infected population of size *n = 1000000*, starting from state $N_2(all)$, resulting in a number of people for each state: Out of state $N_2(all) = 1000000$ 30% get diagnosed, resulting in state $N_3(all) = 300000$. Out of those 15.44% end up in hospital, i.e. state $N_4(all) = 46313$. 39.16% are referred to ICU ($N_5(all) = 18138$) from which 29.27% are released dead ($N_7(all) = 5308$). By splitting the resulting numbers of people per state into their age group fractions (compare **Table S23**), we obtain age group-dependent estimates for the expected number of people in each state for the example scenario. For each state transition, the age group-dependent transition probability $P_{ij}^{c}(age)$ equals the number of people in the end state divided by the number of people in the start state:

$$P_{ij}^{c}(age) = \frac{N_j(age)}{N_i(age)}$$

Equation (9)

with $i,j \in \{1,...,7\}$ and $age \in \{0-59, 60-79, 80-99, all\}$. State transition probabilities $P_{ij}^{c}(age) \neq 0$ are summarized in **Table S24**. Please be aware that rounding resulted in several probabilities appearing to be equal to zero while in reality they are small but non-zero.

| age group | $I \rightarrow I^d$ | $I \rightarrow R$ | $I^d \rightarrow I^d_H$ | $I^d \rightarrow R$ | $I^d \rightarrow D$ | $I^d_H \rightarrow I^d_{ICU}$ | $I^d_H \rightarrow R$ | $I^d_H \rightarrow D$ | $I^d_{ICU} \rightarrow R$ | $I^d_{ICU} \rightarrow D$ |
|---|---|---|---|---|---|---|---|---|---|---|
| 0-59 | 0.29 | 0.71 | 0.06 | 0.93 | 0.00 | 0.33 | 0.64 | 0.03 | 0.94 | 0.06 |
| 60-79 | 0.27 | 0.73 | 0.3 | 0.7 | 0.00 | 0.55 | 0.28 | 0.16 | 0.82 | 0.18 |
| 80-99 | 0.54 | 0.46 | 0.54 | 0.45 | 0.01 | 0.26 | 0.39 | 0.35 | 0.19 | 0.81 |
| all | 0.3 | 0.7 | 0.15 | 0.84 | 0.00 | 0.39 | 0.42 | 0.19 | 0.71 | 0.29 |

**Table S24:** Age group-dependent state transition probabilities $P_{ij}^{c}$ for all state transitions $X_{ij}$ with $i,j \in \{1,...,7\}$. Please note that probabilities equal to zero in this table only do so due to rounding, in Tables S26 (A-D) the same probabilities are given in %, thus avoiding the zeros.

By definition of a non-chronically disease state, this state is transient, i.e. the probability to remain in a state $S_i$ with $i,j \in \{2,...,5\}$ vanishes in the long term. That means that the sum of cumulative state transition probabilities $P_{ij}^{C}$ away from each infected (sub)state *i* must equal 1, i.e.:

$$1 = \sum_{j=1}^{7} \quad P_{ij}^{c} \ \forall i \in \{2, \dots, 5\} \, .$$

Equation (10)

##### 3.4.3.3 Time-Dependent Conditional State Transitions

As explained in section 2.2.7, each agent being in a certain state at a certain time has a probability to stay in that state or leave towards selected other states within the next time step. To this end, we calculated hourly transition probabilities that depend on how long the agent was already in the given state: $p_{ijt}^{X}$ is the required conditional state transition probability per hour for all considered state transitions $X_{ij}$. Based on the age group-dependent cumulative transition probabilities $P_{ij}^{C}$ (**Table S24**), we determined age-dependent conditional state transition probabilities for $p_{ijt}^{X}(age)$, using the following assumptions:

1. The transition occurs within a given maximum time (50 days).

2. The probability for each state transition $X_{ij}$ is Gaussian distributed with mean at the reported or assumed average time $\mu_{ij}^{t}$.

3. The standard deviation of this Gaussian distribution equals its mean, i.e. $\sigma_{ij}^{t}$ = $\mu_{ij}^{t}$

From the data, we extracted cumulative transition probabilities $P_{ij}^{C}$ of a state transition $X_{ij}$ in a defined time frame and the reported average time $\mu_{ij}^{t}$ for the transition to occur. For example, if 30 % of cases died after being transferred to ICU and half of them died during the first 10 days after being transferred to ICU, the cumulative transition probability $P_{57}^{c}$would be 0.3 and the mean time would be $\mu_{45}^{t} = 10 \; [days]$.

From assumption 2 it follows that the transition probabilities are drawn from a probability mass functions (PMF), generated from a Gaussian probability distribution and weighted by $P_{ij}^{c}$ of the respective transition.

$$P_{ijt}^{X} = P_{ij}^{c} \cdot PMF \;\; (t \colon \mu_{ij}^{t}, \sigma_{ij}^{t})$$

Equation (11)

The required weighted distributions were calculated from information about the average transition times determined by the model from (*34*), **Figure 1**). For all distributions, we assume the maximum transition time to be 50 days and the mean day was taken from this reference, re-drawn here in English for readability (**Figure SY**).

Because the underlying model of an der Heiden & Buchholz does not consider death from states other than ICU, we assume the same total average time from time of infection until death for the different states. Due to irreversibility of state transitions it is not possible to be referred back to hospital from ICU in the GERDA model. Therefore, we assume the average duration in ICU (state $I^d_{ICU}$) before recovery to be 13 days (until the average release from hospital when referring to the time of infection). In summary, we used the mean transition days given in **Table S25** to generate Gaussian distributed transition probabilities $P^X_{ijt}$:

| transition | $I \to I^d$ | $I \to R$ | $I \to D$ | $I^d \to I^d_H$ | $I^d \to R$ | $I^d \to D$ | $I^d_H \to I^d_{ICU}$ | $I^d_H \to R$ | $I^d_H \to D$ | $I^d_{ICU} \to R$ | $I^d_{ICU} \to D$ |
|---|---|---|---|---|---|---|---|---|---|---|---|
| mean day | 5 | 13 | 19 | 4 | 9 | 15 | 1 | 14 | 11 | 13 | 10 |

**Table S25:** Average day after state entering for a state transition to occur. For complete model input tables used to calculated the final hourly resolved dependent state transition probabilities $P^X_{ijt}$ compare supplementary tables file, **Tables S26 (A-D)**.

From the weighted distributions and the age group-dependent overall probability to undergo a specific state transition, we calculated the conditional transition probabilities per time step as explained in 2.2.7. An illustration of the resulting probabilities is provided in **Figure S6**.

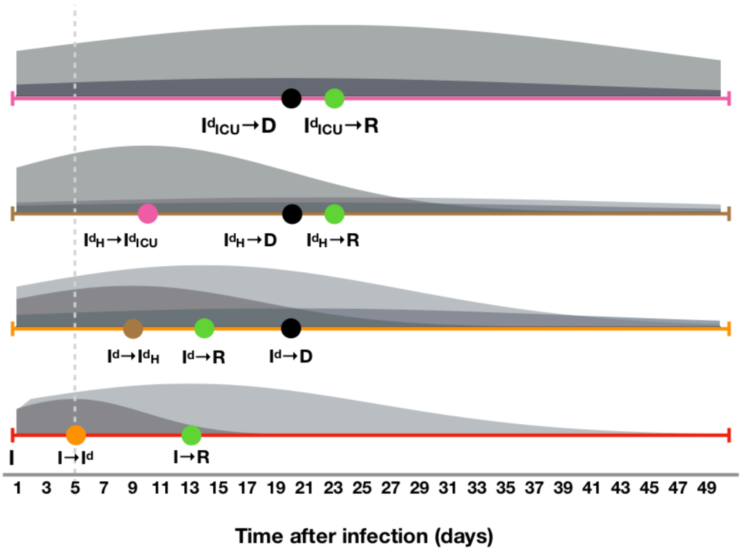


**Figure S6**: **Schematic representation of the conditional probability for transition from $S_i$ to $S_j$ (per time step, plotted against time after infection in days).** GERDA considers 11 transitions with non-zero probability. At each time step, an infected agent (states $S_2 - S_5$) can either remain in its current state, progress to another infected state or to one of the final states R ($S_6$) and D ($S_7$). Here, we show for a 64 year old agent the weighted distributions for the time-dependent probability to undergo a specific state transition. The probability for each state transition is Gaussian distributed with mean at the reported or assumed average time $t_\mu$ and standard deviation equal to mean, i.e., $t_\sigma = t_\mu$ . The dashed line represents the average time to symptoms onset and the colored circles the mean occurrence times for state transitions.

#### 3.5 Infectiousness of an infected agent

The infectiousness$I^I_a(t)$ of an infected agent $a$ is a function of time, starting with the time of infection. Background is that the reported time-dependent development of symptoms and viral load (representing the individual infectivity) do not coincide (*35*). Instead, the infectivity peaks on day 2 after infection before symptoms onset on day 3. To represent pre- and asymptomatic transmission in GERDA, we used a Gaussian probability mass function to calculate the individual infectiousness. Its mean and standard deviation have been set to day 2, similar as described for the Gaussian distributions in subsection 3.4.3 (**Table S26 (E)**). Although data on the timing of infectiousness is available (*35*), information on the intensity is still missing, therefore we weighted the obtained time-dependent probability distribution for the infectiousness by a global infection parameter $k_I$ (compare next section 3.6).

#### 3.6 Additional Parameters

As a design principle, the parameter values required for the simulation of GERDA are not hard-coded, but simulation input. They can either be calculated from input data, as explained above, or they can be specified by the user when setting up the simulation. This allows us to change the parameter set between individual simulation runs, as explained below.

In section 3.4, we have described the time-dependent probabilities for state transitions. More parameters have to be set in order to enable a simulation. These are:

- global infection parameter $k_I$ (short: infectivity; required),
- global interaction frequency $\mu$ (required),
- non-compliance probability $p^{ncomp}$ (optional).

Note that these parameters can be set for the entire population or for population subgroups only.

Furthermore, start and end times for lockdown (closure) and reopening for all or for specific location types can be defined. For the required commands to set those parameters manually, please checkout the readme on our public GERDA Gitlab repository (https://tbp-klipp.science/GERDA-model/readme).

### 4 Simulation and Analysis of GERDA

Providing a simulation origin (an initialized MW) and the required and optional input parameters, GERDA can be applied for simulation of infection transmission throughout an agent population. At each simulation time step, for each agent it is resolved i) which of its putative interactions take place, ii) which of its realized interactions bear the potential of an infection transmission, and iii) which of its realized interactions effectively result in an infection transmission (subsection 2.2.4). In addition, for infected agents it is determined how the infection proceeds based on given state transition probabilities that depend on the time of infection and the agent's age. The calculation of cumulative time and age-dependent transition probabilities is detailed in subsection 3.4.

Alternatively, the simulation can also be started from a previous simulation output. This allows for the comparison of different scenarios starting from a given time point.

The simulation can be run from GitLab. The GitLab repository (https://tbp-klipp.science/GERDA-model/) also provides ample information about code, settings, and parameters. We provide a "cookbook" for interested users explaining which data and information to use in order to apply the model to different locations. The major functionalities are described in the following.

Below, we briefly summarize the basic output of the simulation of GERDA. Of course, it is always possible to add more functionalities for further analysis, which shall be omitted here.

#### 4.1 Simulation time course and outputs

The time course of a simulation describes for each agent at each time step its

- location,
- health states (including infection sub-states),
- interaction partners,
- agent from which an infection was obtained.

Additional information, such as interaction and infection networks, frequencies and distributions of state transitions and durations for the transient infection states, or household-related transmissions can be directly inferred. This allows us to monitor infection spreading and disease progression on the individual level, thereby providing unprecedented insight into the effects of non-pharmaceutical intervention strategies, preventive measures like contact tracing, and the individual compliance.

It is possible to store complete and partial time courses. Time course simulations can be repeated (either full simulations or simulations starting from a given time point) in order to analyse the effect of stochasticity on the course of infection spreading.

##### 4.1.1 Simulation object at final time step

Together with the locations the agent population at simulation end time is combined into a simulation object that shares most of its properties with a MW object. In addition, it contains the simulation time course to allow plotting and analysis of consecutive simulation runs. It can optionally be stored (memory and time consuming).

#### 4.2 Simulation of specific scenarios

In order to investigate the effect of infection spreading, non-pharmaceutical interventions or vaccination strategies, it is possible to simulate specific scenarios.

The following scenarios are currently foreseen:

- Baseline scenario (spread of infection starting with one or few infected agents, **Figure 1F,G**)
    - Location-dependent behavior versus homogenous mixing (as control, see section **4.7**)
- Non-pharmaceutical interventions, including
    - Lockdown (temporarily or continued, **Figure 1H** and section **4.3**)
    - Selective lockdown of specific location types
    - Lockdown and full or selective reopening (section **4.3**)
    - Re-infection, test for community protection
- Non-compliance with non-pharmaceutical interventions such as lockdown (**Figure 1K**, section **4.3**)
- Combinations of the above

In the baseline scenario, we change the state of 4 random individuals from S to I and let the infections spread through the population according to the stochastic interactions that agents have when visiting locations corresponding to their schedule. No mitigation measures are applied. Default parameter values are infectivity $k_I$=0.3 and global interactivity $\mu$=2.

Lockdown describes in our model a situation, where agents stay at home instead of going to school, to workplaces or to public places. Agents, who are scheduled to go to hospital, continue so (medical personal or hospitalized agents). Selected lockdown means that only one or two location types out of {schools, workplaces, public places} are closed and individuals stay at home instead of visiting them.

Lockdown light represents closing public spaces only.

Reopening means that all agents follow their normal schedules (if in state S, I, or R) or their quarantine ($I^d$) or hospitalization ($I^d_H$, and $I^d_{ICU}$). Selective reopening indicates that the respective location can be visited again.

Non-compliance: We also investigated what happens if a number of individuals will not follow the order for closure of all locations. This reflects either non-compliance (e.g. civil disobedience) or the fact that some people in systemically relevant jobs have to go to work or need to send their children to school (or daycare) to be able to go to work themselves. Already small levels of non-compliance have severe effects (section **4.4**).

#### 4.3 Duration between agents' state transitions

In order to test whether the state transition rates, defined as input to the model, result in simulated distributions, which are similar to the empirical ones; the distributions of durations between specific state transitions in a simulated baseline scenario are presented together with the empirical distributions in **Fig. S7** below.

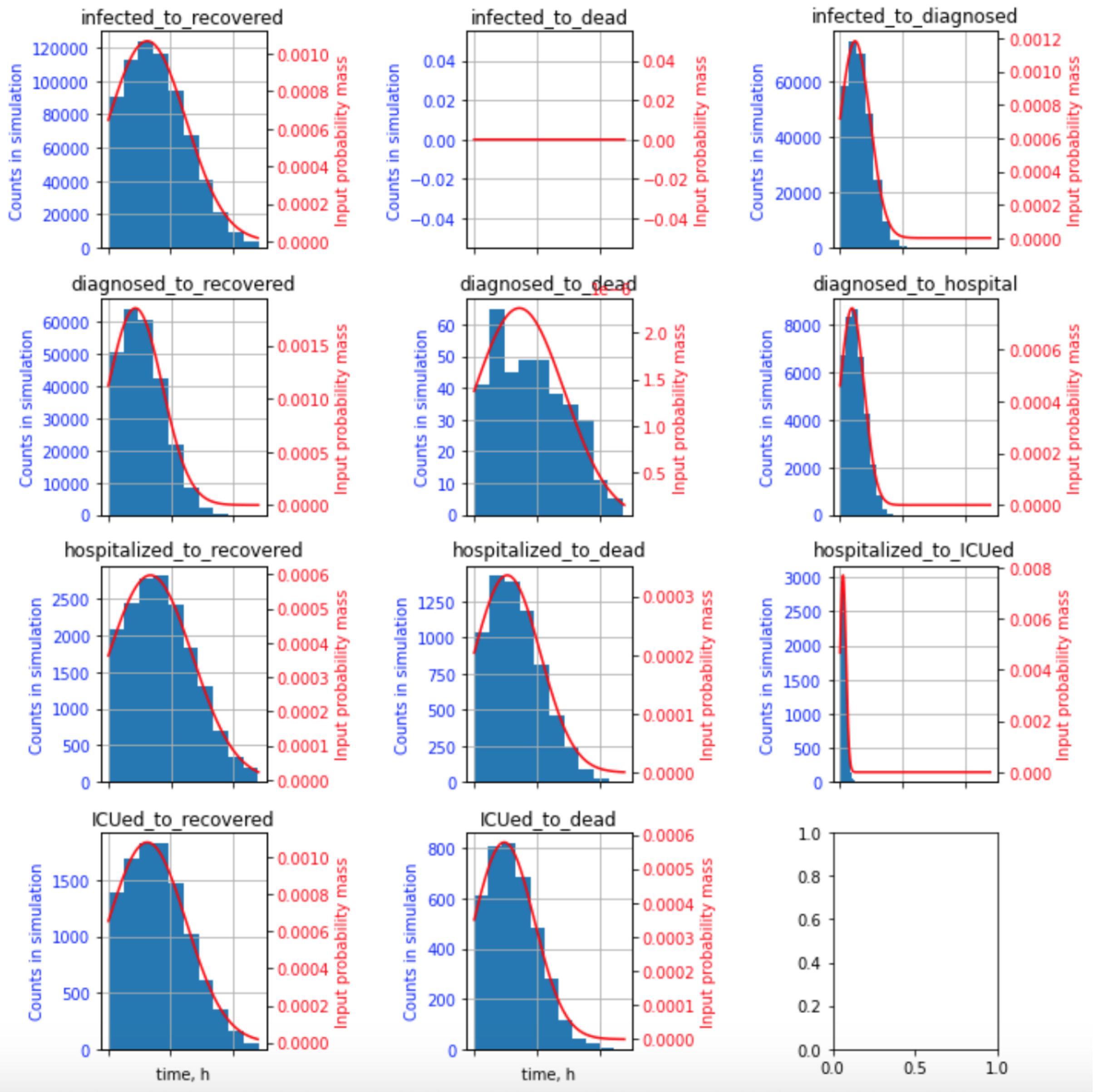


**Figure S7: Comparison of input distributions for the agents' state transitions to distributions resulting from the simulations.** Presented are the distributions of the indicated transitions.

#### 4.4 Sensitivity analysis

In order to better understand the impact of the choice of parameter values as well as the capabilities of our model, we can perform sensitivity analysis. Specifically, one can

- vary the global infection rate $k_I$ (**Fig S8**)
- vary the global interaction frequency $\mu$, (**Fig S9**)
- vary the non-compliance probability $p^{ncomp}$ (**Fig S10**)
- change start and end times of non-pharmaceutical intervention measures (**Fig S11** - variation of lockdown start time, **Fig S12** - variation of reopening times, **Fig S13** - variation of reopening times for public places only**, Fig S14** - variation of reopening times for schools only**, Fig S15** - variation of reopening times for workplaces only**, Fig S16** - variation of start times for lockdown light)
- apply specific measures to either specific age groups, groups with different occupations (e.g. pensioners), or specific locations.
- vary the fraction of non-susceptible individuals, i.e. either recovered or vaccinated individuals, at the start of simulation (section **4.6**)
- study the impact of the infection rate on the attack rate and bimodality (**Fig S17**)

Sensitivity analysis enables systematic comparison of model results with input data. It also points to specific behaviors of the model that only occur for specific parameter values.

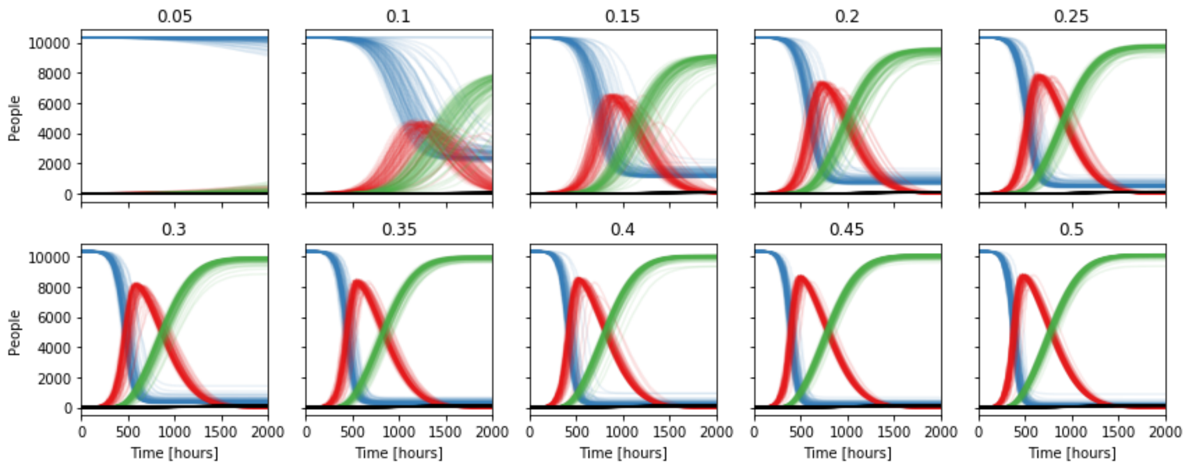


**Figure S8: Infectivity scan from 0.05 to 0.5 (Gangelt).** Resulting time courses for agent health states S (blue), I (red), R (green), and D (black) from 100 simulations for varying infectivity $k_i$.

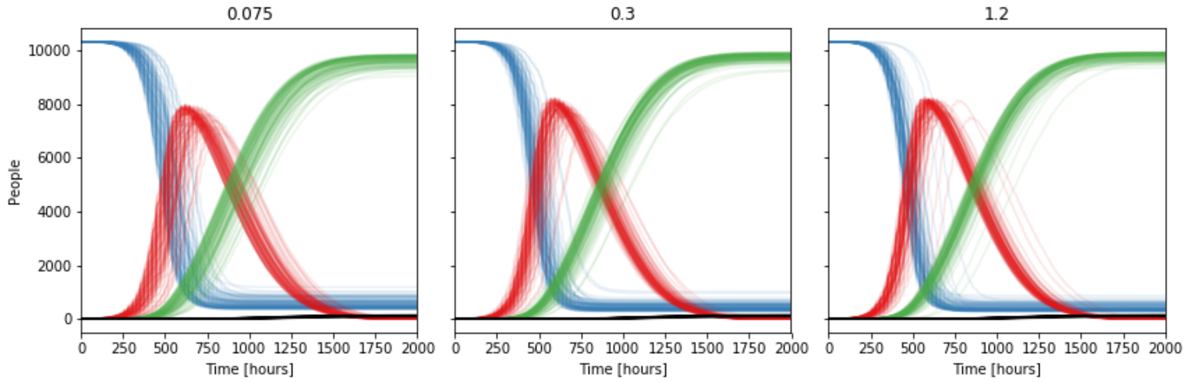


**Figure S9: Product composition scan (global infectivity $k_i$ * global mean interaction frequency $\mu$ = 0.6; varying $k_i$ from 0.075 ($\mu$=8) to 1.2 ($\mu$=0.5); Gangelt).** Resulting time courses for agent health states S (blue), I (red), R (green), and D (black) from 100 simulations for varying interaction frequency and infectivity (keeping their product constant).

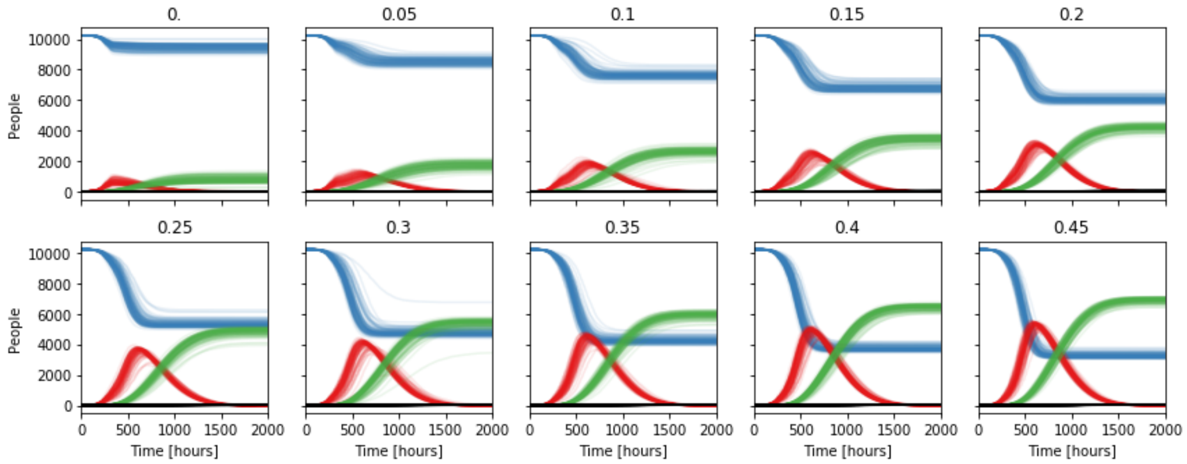


**Figure S10: Non-compliance fraction scan from 0.0 to 0.45 (Gangelt).** A scenario with lockdown starting at 200h has been simulated. The indicated fraction of individuals did not or could not obey the lockdown, but followed instead their normal schedules. Resulting time courses for agent health states S (blue), I (red), R (green), and D (black) from 100 simulations for varying fractions of non-compliant agents.

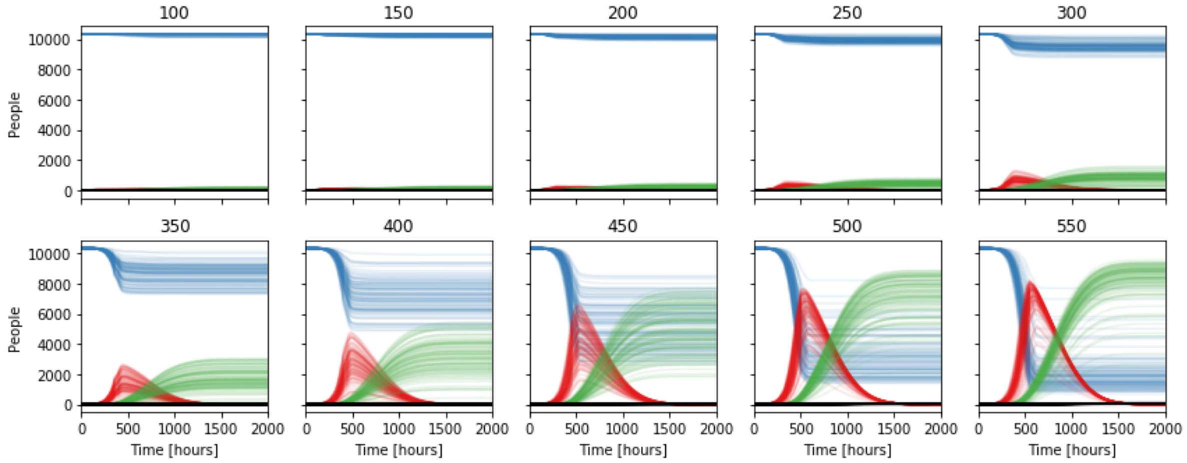


**Figure S11: Lockdown start time scan from t= 100 hours to t = 550 hours (Gangelt).** Resulting time courses for agent health states S (blue), I (red), R (green), and D (black) from 100 simulations for varying lockdown start times.

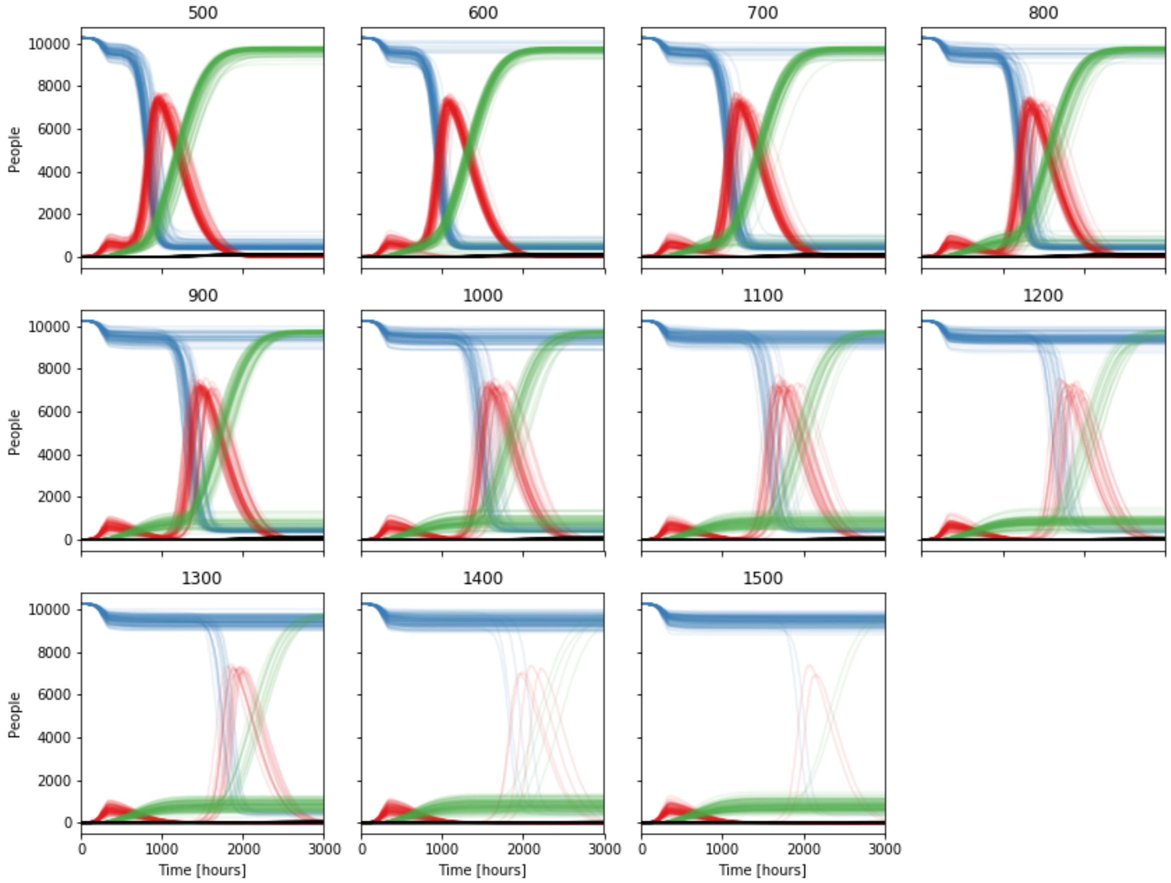


**Figure S12: Reopening times scan from t = 500 to t = 1500 (reopen all; Gangelt).** Resulting time courses for agent health states S (blue), I (red), R (green), and D (black) from 100 simulations for varying reopening times after closing all at t=200.

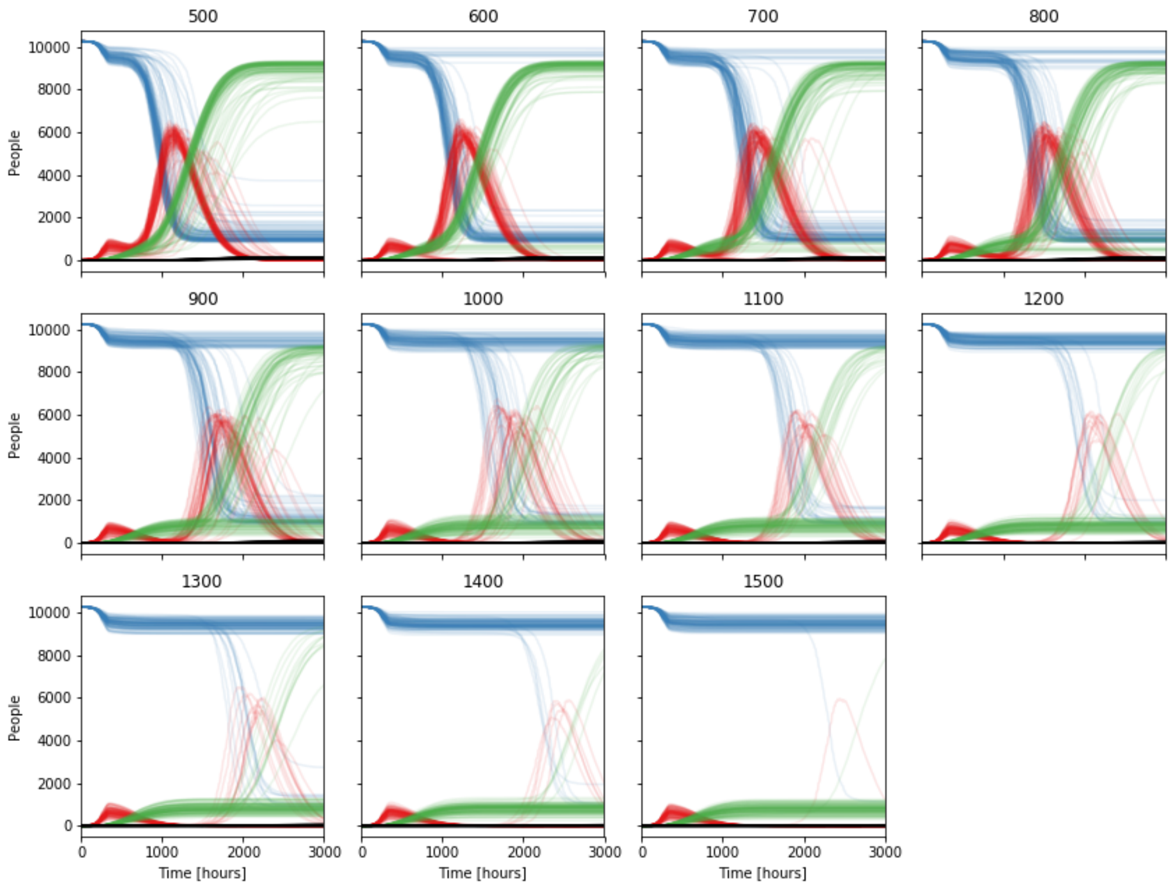


**Figure S13: Reopening times scan from 500 to 1500 (reopen public places; Gangelt).** Resulting time courses for agent health states S (blue), I (red), R (green), and D (black) from 100 simulations for varying reopening times after closing all at t=200.

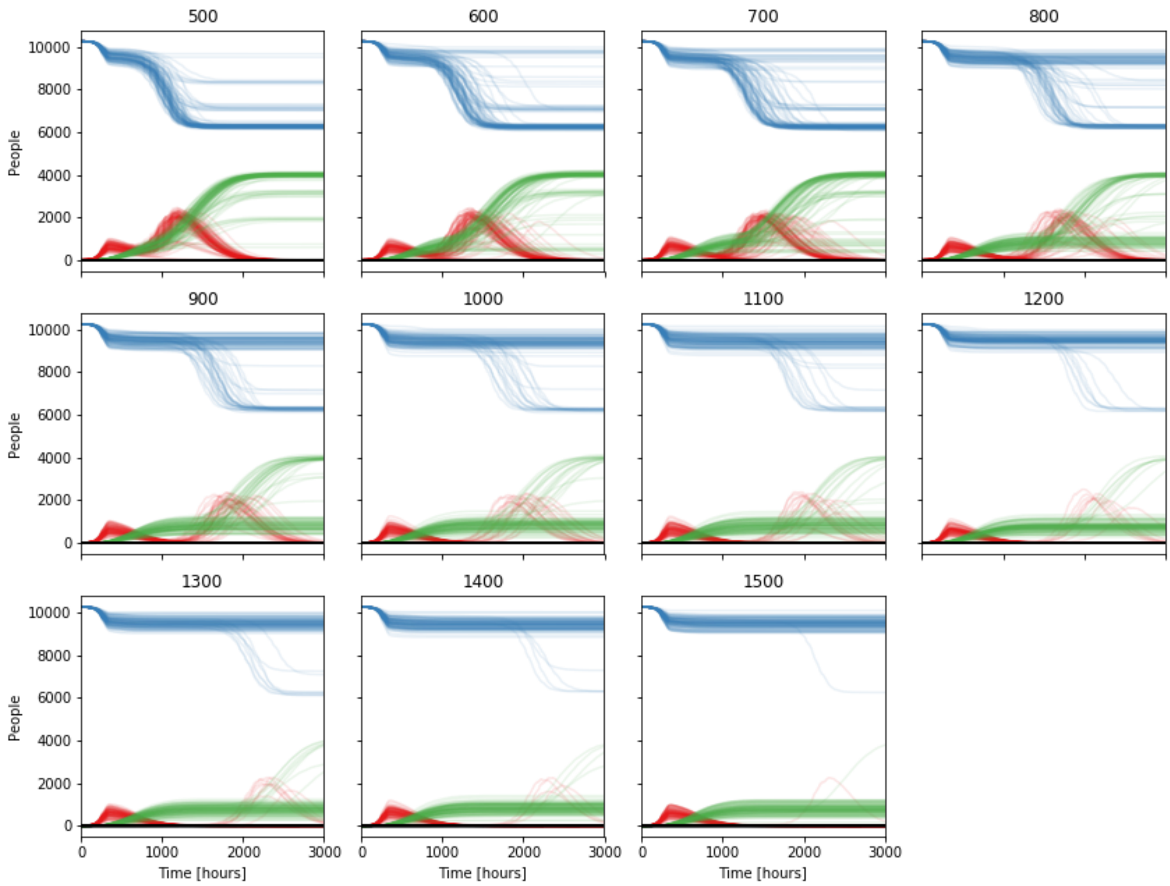


**Figure S14: Reopening times scan from 500 to 1500 (reopen schools; Gangelt).** Resulting time courses for agent health states S (blue), I (red), R (green), and D (black) from 100 simulations for varying reopening times after closing all at t=200.

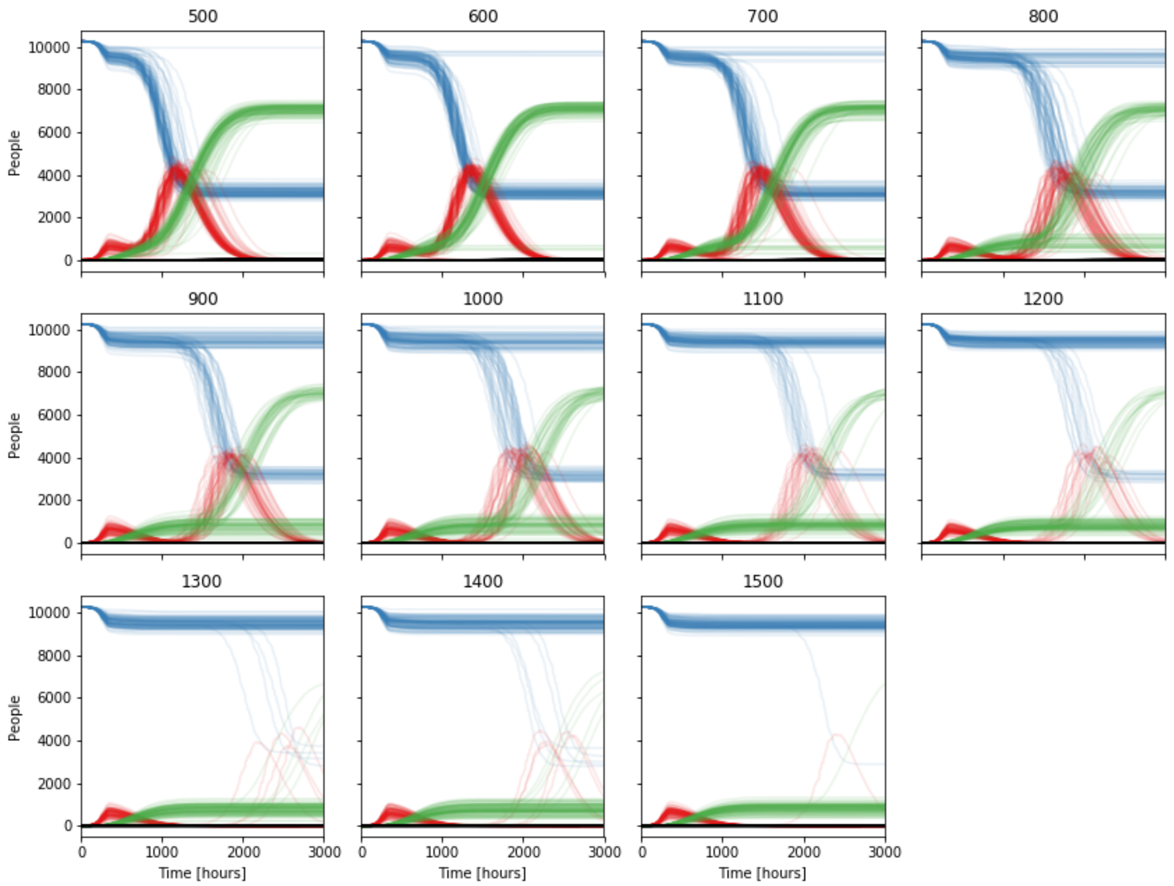


**Figure S15: Reopening times scan from 500 to 1500 (reopen work; Gangel).** Resulting time courses for agent health states S (blue), I (red), R (green), and D (black) from 100 simulations for varying reopening times after closing all at t=200.

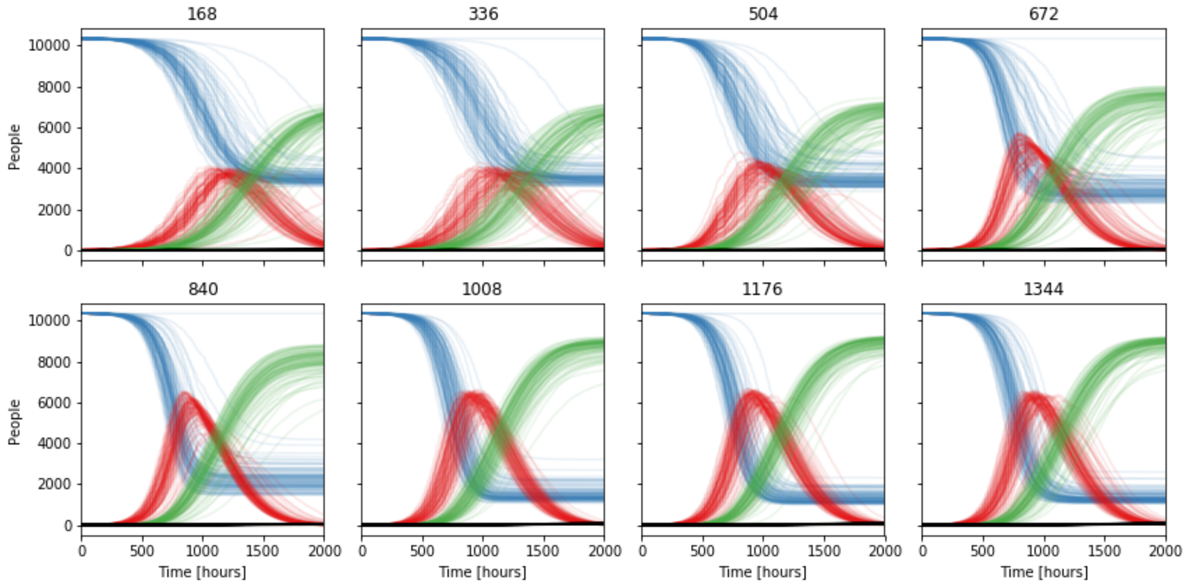


**Figure S16: Lockdown light starting times scan (Gangelt).** Resulting time courses for agent health states S (blue), I (red), R (green), and D (black) from 100 simulations for varying lockdown start times. Lockdown light is represented by closing public spaces only.

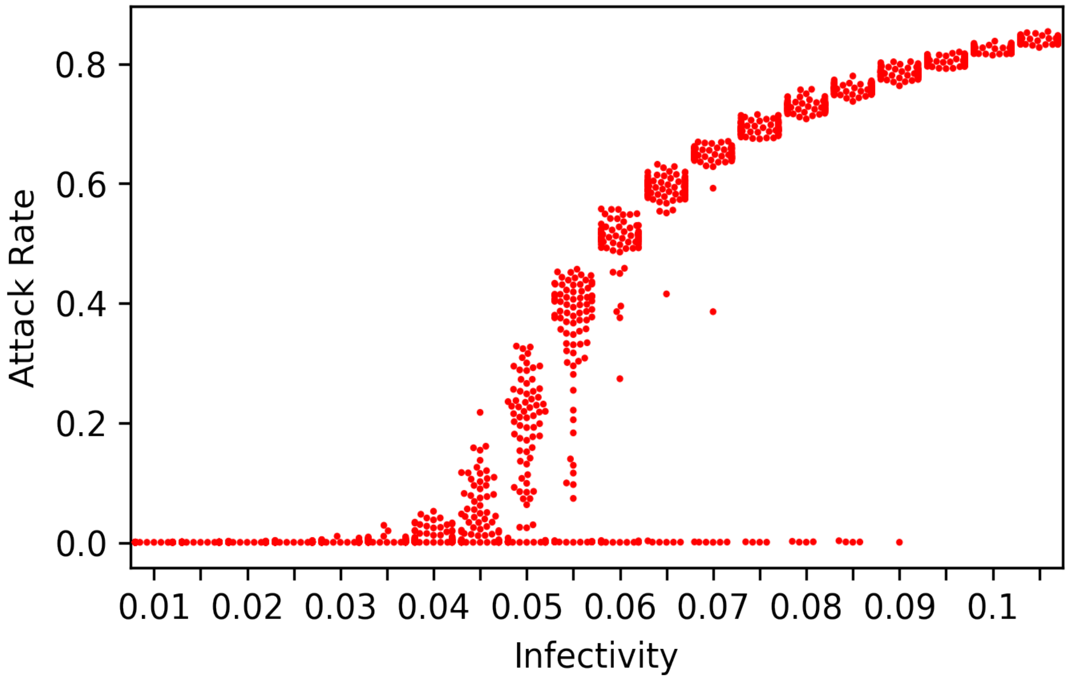


**Figure S17: Attack Rate vs. Infectivity (Gangelt).** Attack rate (total infected / initial susceptible) for 100 simulations and different $k_I$. Note bimodal outcome for the range $0.03 < k_I < 0.09$.

#### 4.5 Network analysis

##### 4.5.1 Interaction networks

The complete simulation time course allows us to reconstruct the (time-dependent) interaction network after a simulation. The complete network graph is built by adding new edges (and corresponding nodes if not yet existing) for each first appearance of a unique pairwise interaction over the entire simulation time. It is possible to define specific time frames to create and analyze subsets of interactions. Counting the number of occurrences for unique pairwise interactions provides a weighted graph.

##### 4.5.2 Infection networks

The infection network is a directed subset of the interaction network, including only those interactions during which the infection was transmitted to a new agent. This enables us to follow individual infection transmission chains. An example of the emerging network is represented in **Fig. S18**.

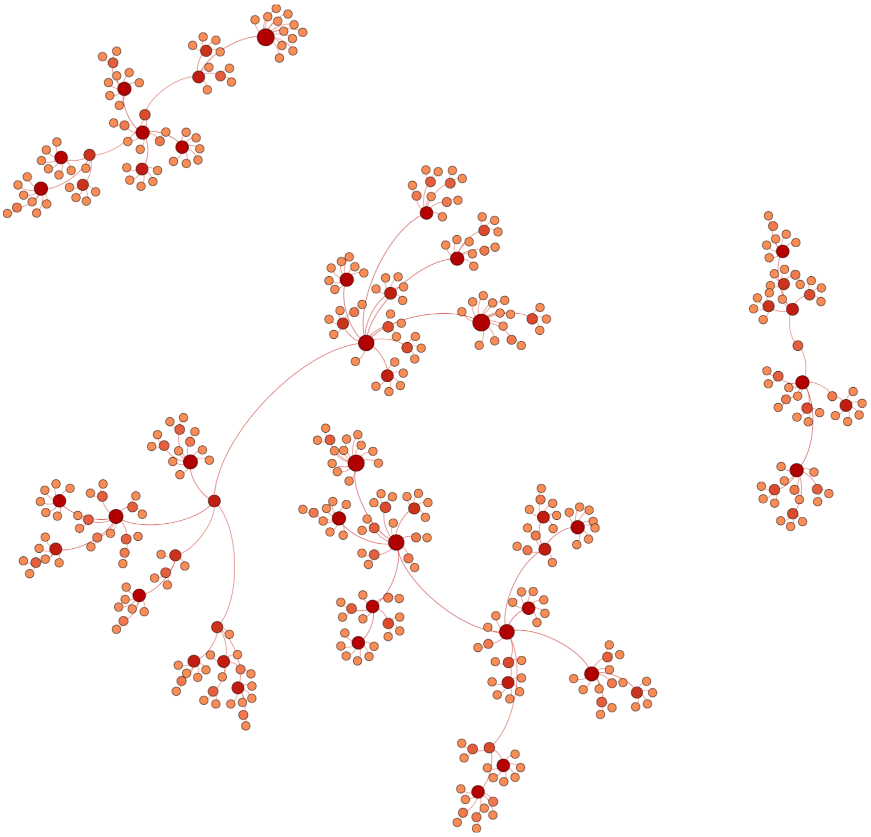

**Figure S18: Infection networks in Gangelt unfolding within 300 hours (single simulation).** Starting from four infected individuals other individuals get infected creating initially unconnected networks (which may or may not become connected later).

##### 4.5.3 Overrepresentation and underrepresentation of specific groups in the infection process

For example, comparing the distributions of agents that transmit infections and of locations at which infections are transmitted to the respective distributions of initialized agents and locations, provides a possibility to investigate the impact of specific population subgroups and location types on infection spreading. In particular, we analyzed the difference in the contribution to infection spreading between different occupations, as can be seen in the over- and underrepresentation plots (**Fig S19).** Additionally, we analyzed the average infection caused by an emitter, resolved for its occupation **(Fig. S20**).

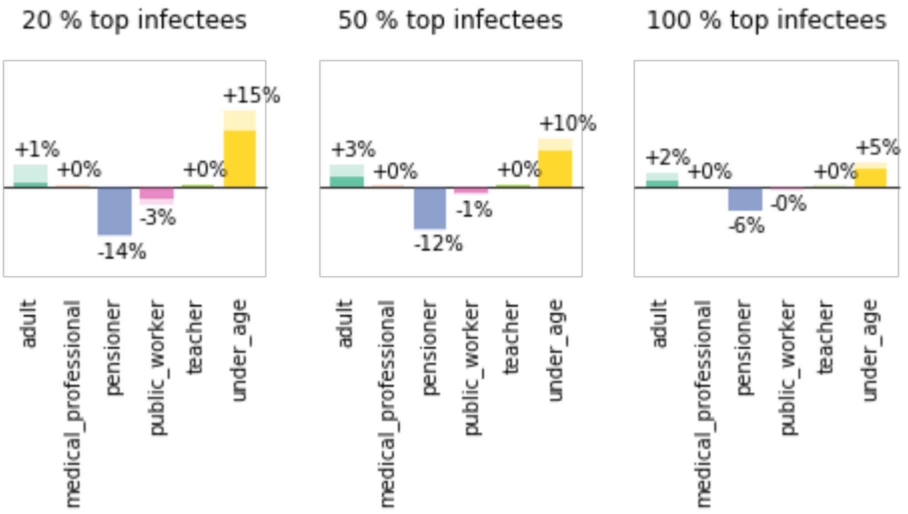


**Figure S19: Over and under representation of agents with specific schedule types in the fractions of top spreaders.** Difference in the fraction of spreaders with specific schedule type to the fraction of this type in the population.

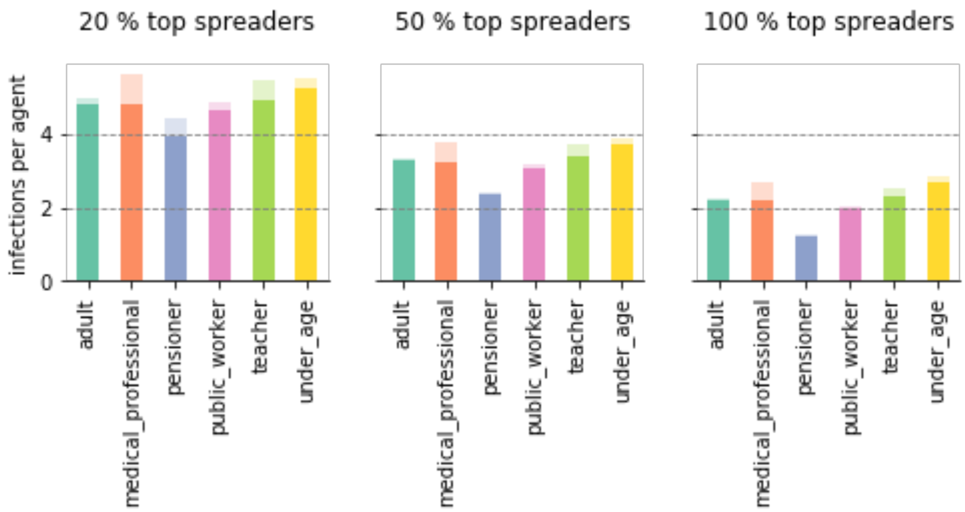


**Figure S20: Pattern in schedule type dependent infection rates.** Average infections caused by each agent resolved for schedule type and sorted for percentage of top spreaders. Shaded area corresponds to standard deviation of mean.

#### 4.6 Immunization Scans

In order to analyze the effect of immunization on the infection dynamics, we set a fraction of individuals to status R, since recovered individuals are not considered susceptible to infections anymore. Hence, the amount of initially recovered individuals can mimic the fraction of vaccinated or immunized individuals. Systematically introducing infections in such a partially vaccinated population allows us to analyze the robustness of the vaccination and the remaining burden.

All simulations are based on the same modeled world, e.g. the same locations, the same agents and the same daily routines. Prior to the simulations we defined sorted lists of individuals, reflecting different immunization strategies. The simulations were initialized with different ratios of susceptible and recovered agents, by setting individuals to health state R based on their occurrence in our predefined sorted lists. We scanned ratios of susceptible and recovered individuals ranging from 0.1 to 0.9 with a step size of 0.04 (**Figs S21-S24**). All ratios were repeatedly simulated for 3000 hours one hundred times (N=100) with four initial infection emitters chosen from the remaining pool of susceptible individuals. All simulations were analyzed with respect to appearance of an infection wave (i.e. showing more than 80 infection events during the simulation), the fraction of infections relative to the remaining susceptibles, the fraction of deceased individuals relative to the remaining susceptibles, and the number of ICU cases per 100 000 based on the whole population (the maximal number reached during the simulation).

**Sorting algorithms for the analyzed immunization strategies:**

- **forecasted**
  Before carrying out the simulations with a given percentage of individuals set to

immunized, pre-simulations have been performed to forecast potentially infected individuals. The lists are based on 100 complete infection chains from 100 preceding simulations starting always with the same initial infection emitters. Resulting infection receivers were sorted by infection time and the precise given percentage was set to R.

- **random**
  100 different lists of individuals were shuffled randomly and the given percentage set to R.

- **age**
  One list of all inhabitants of the community was used, sorted by decreasing age of the individuals.

- **interactions**
  One list was used, sorted by decreasing mean absolute contact numbers, again resulting from 100 preceding simulations, but here without infection spreading.

- **overrepresentation**
  One list was used, sorted by occupations, starting with occupations showing the strongest overrepresentation in infection (Figure 2F) to occupations with the least overrepresentation in infection. (UA, AD, TE, MP, PW, PE)

- **combined**
  One list of three consecutive blocks was used. The first block contained all individuals above age 60 with decreasing age (as in list "age"). The second block contained all individuals from age 13 to age 59 sorted for mean absolute contacts as in "interactions". The third block contained all individuals below age 12 with decreasing age.

It is important to note, that the performance of the different strategies depends on the infectivity $k_I$, i.e., while the general pattern between the different strategies is preserved, the exact numbers change for preventing an infection wave or reducing the hospitalizations or ICU beds required below health care capacities (**Fig. S25**). A ranking of vaccination strategies for infectivity $k_I = 0.3$ is shown in **Fig. S26**. **Fig. S27** analyses the effect of threshold values for deciding if the number of infections amounts to a next infection wave.

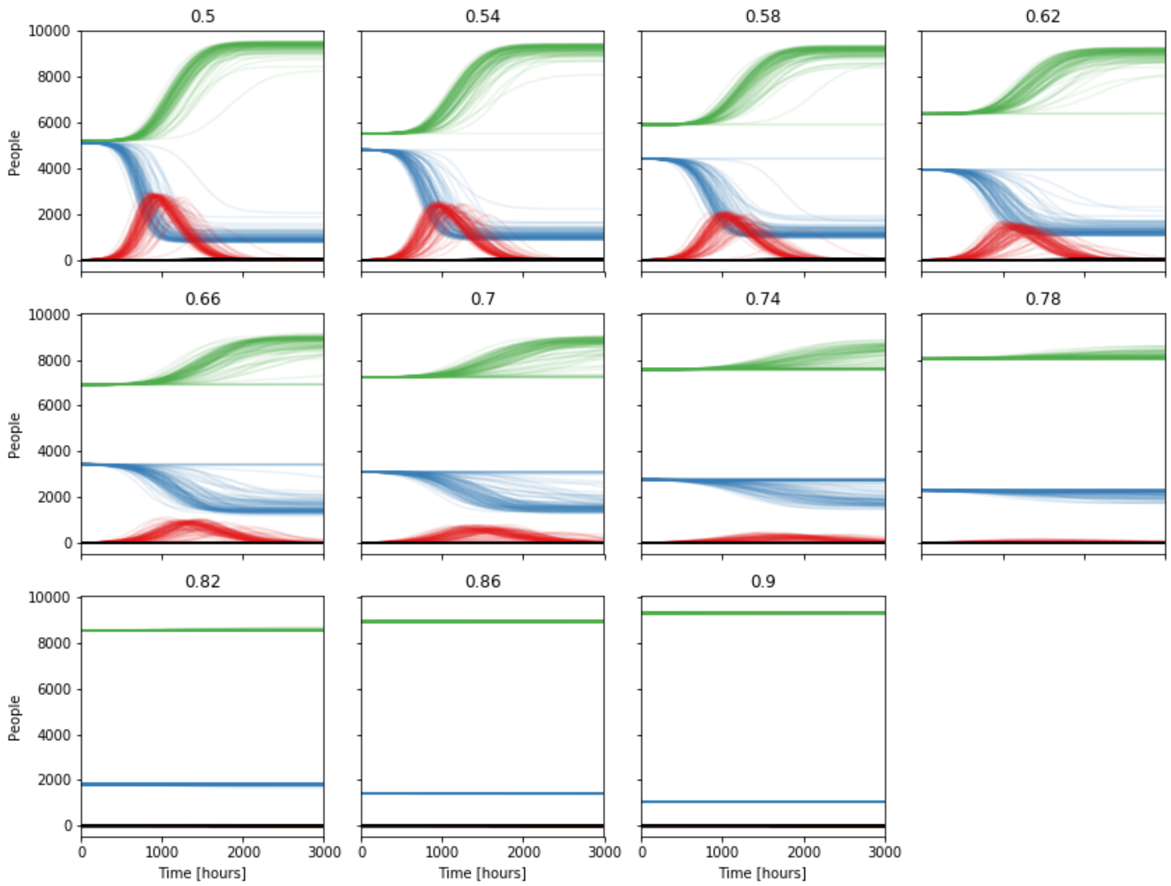


**Figure S21: Interaction-dependent recovery fraction scan for a share of 0.54 to 0.9 non-susceptible individuals (Gangelt).** Resulting time courses for agent health states S (blue), I (red), R (green), and D (black) from 100 simulations for varying fractions of recovered (vaccinated) agents.

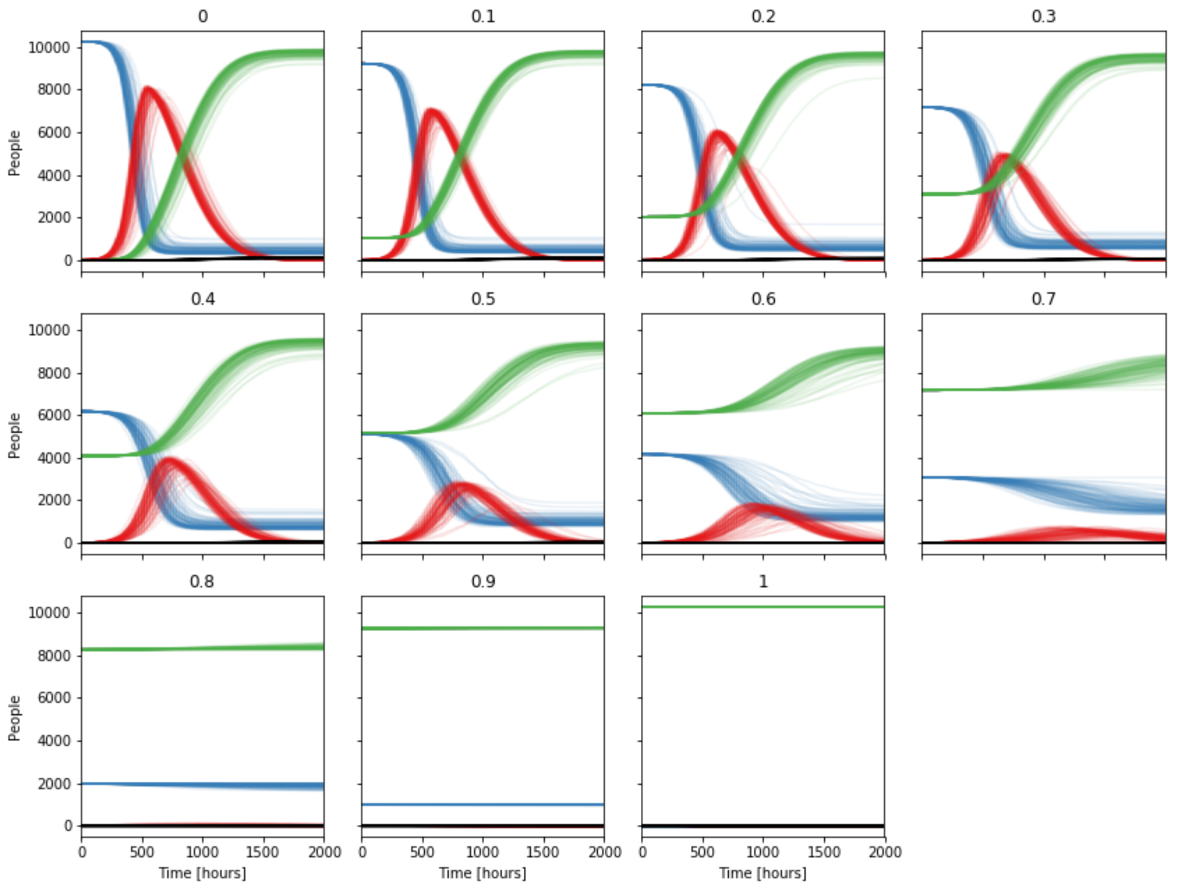


**Figure S22: Random recovery fraction scan for a share of  0 to 1 non-susceptible individuals (GangeIt).** Resulting time courses for agent health states S (blue), I (red), R (green), and D (black) from 100 simulations for varying fractions of non-susceptible  (recovered or vaccinated) agents.

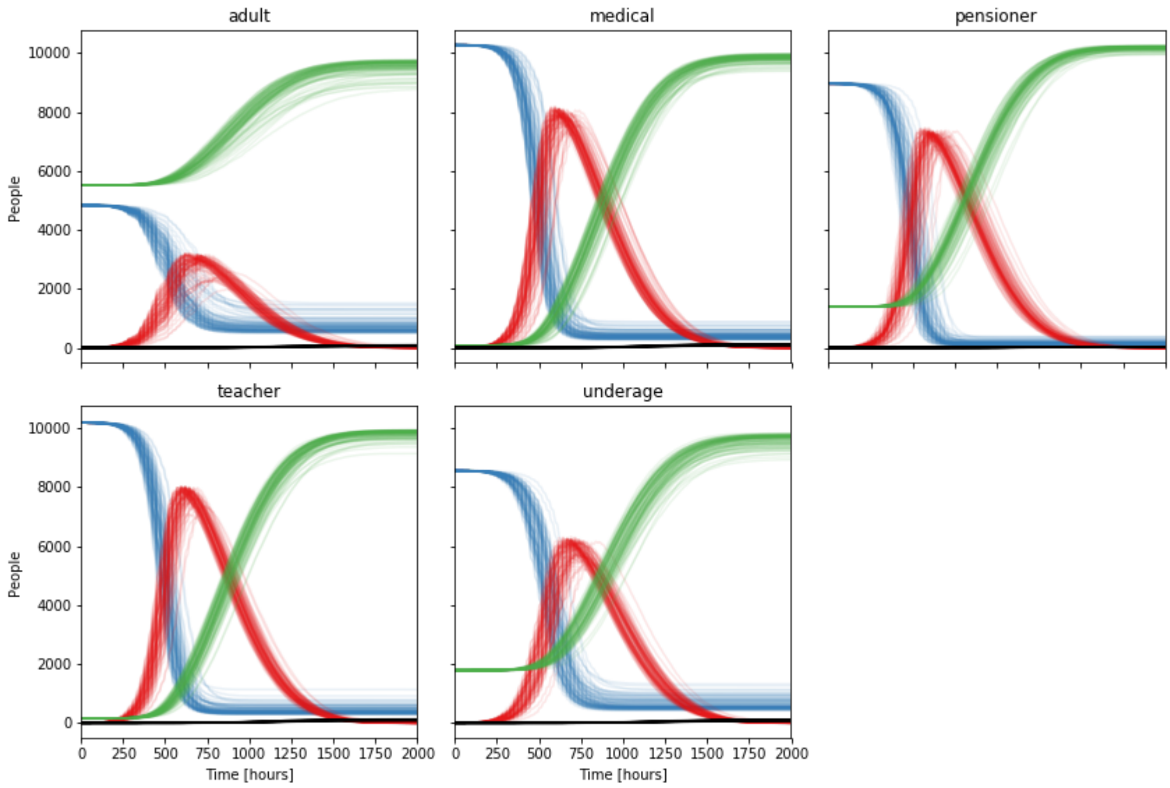

**Figure S23: Recovering (vaccination) by schedule type scan (Gangelt).** Resulting time courses for agent health states S (blue), I (red), R (green), and D (black) from 100 simulations for recovering (vaccinating) a specific schedule type.

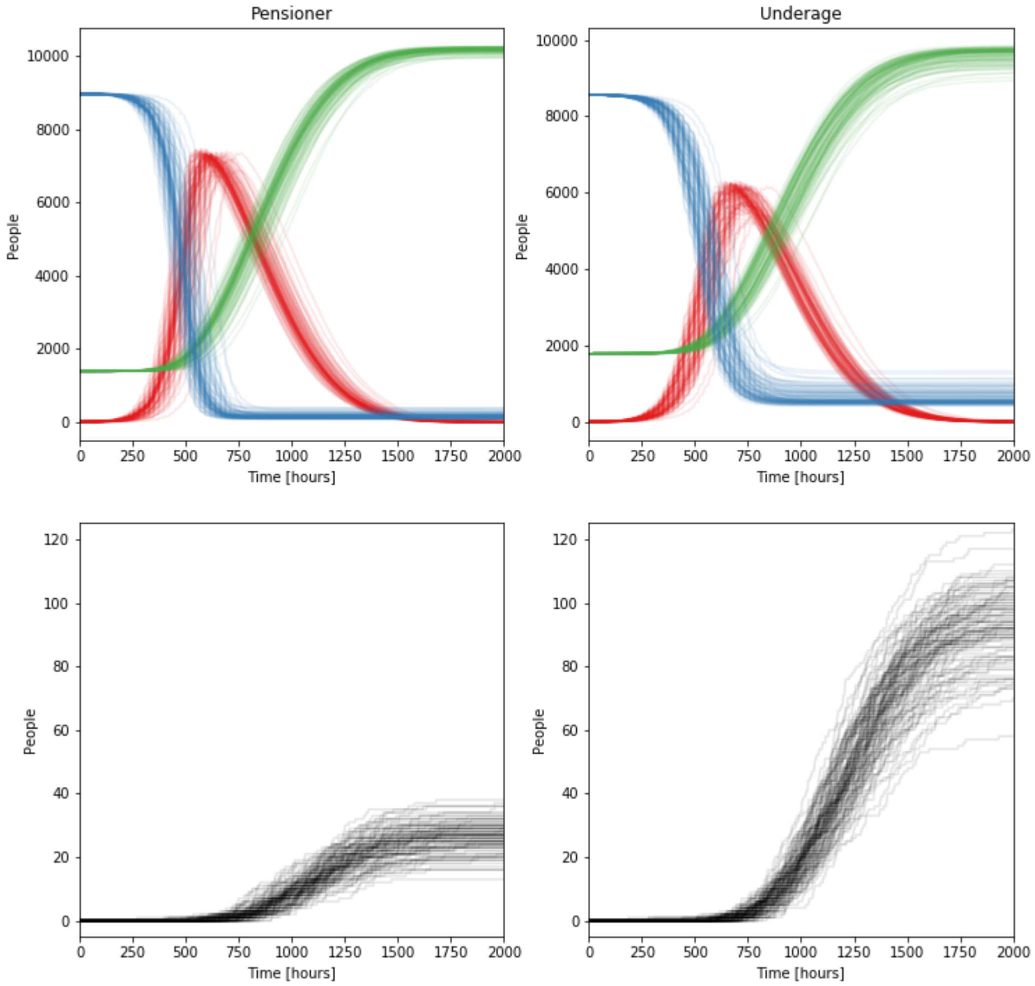


**Figure S24: Comparison of the effect of immunizing (set to R) either pensioners or underage individuals (Gangelt).** Time courses for agent health states S (blue), I (red), R (green), and D (black) from 100 simulations for recovering (vaccinating) a specific schedule type. Immunization of (the whole group of) pensioners leads to comparatively low death numbers but still significant infection dynamics, while immunization of (the whole group of) underaged lowers the infections, but essentially not the death toll.

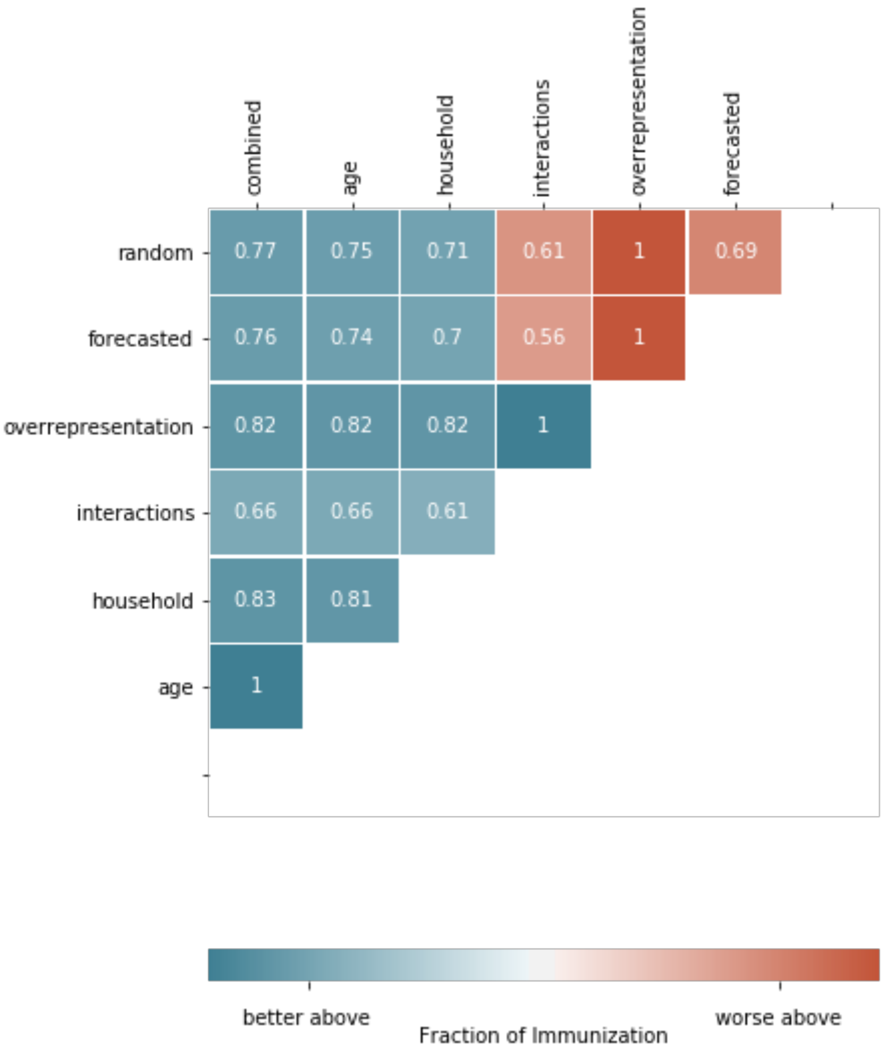


**Figure S25: Vaccination performance with respect to minimize fatalities.** Strategies (rows) perform better (blue) or worse (red) above the shown values compared to strategies shown in the columns. Values represent the non-susceptible population fraction as a measure for vaccination coverage. Parameter value $k_l$=0.3.

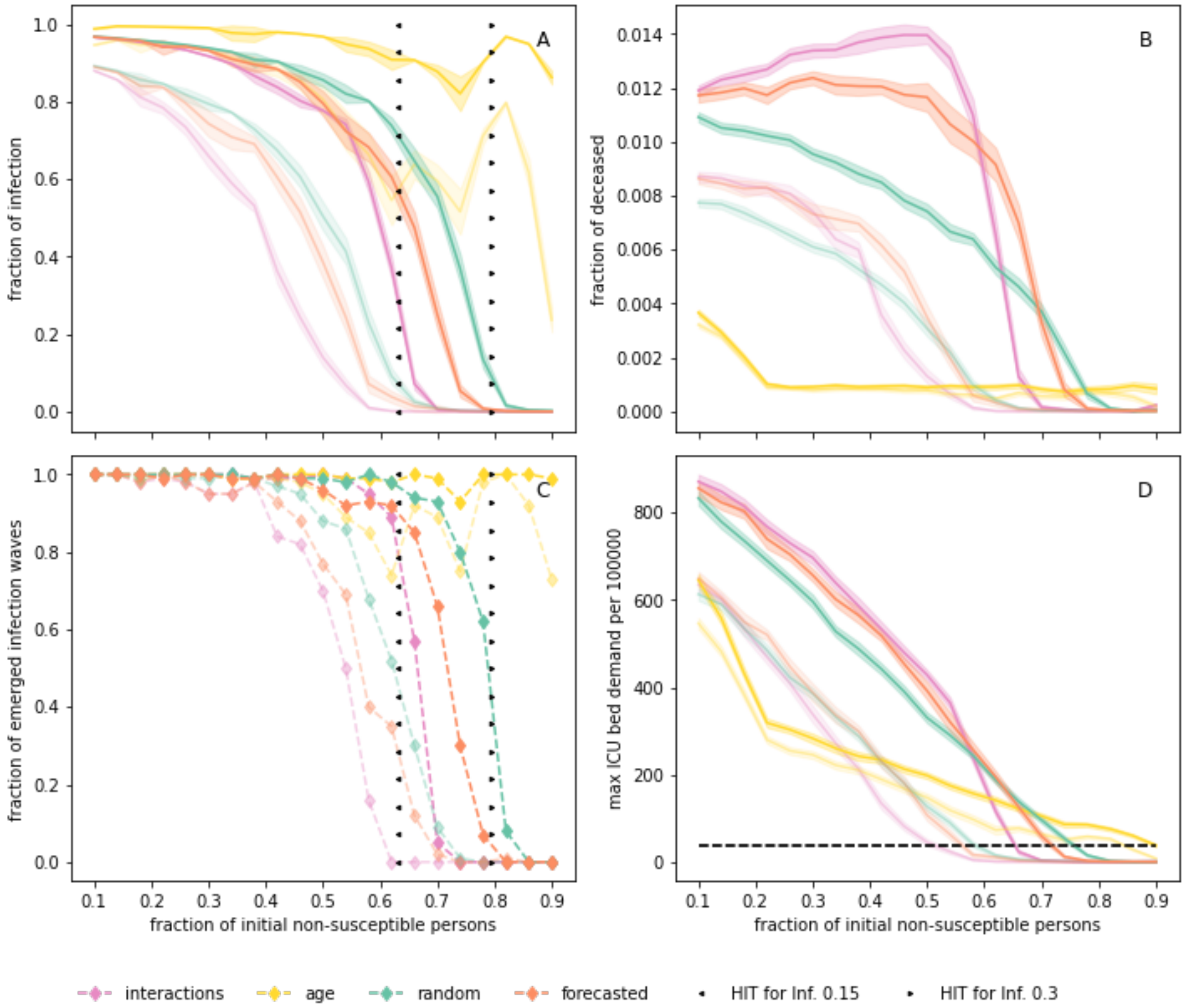


**Figure S26: Vaccination performance dependent on infectivity $k_i$.** Vaccination by interactions, age, random, and forecasted for infectivities 0.15 and 0.30, purple, yellow, green and orange, respectively. Dark colors correspond to infectivity 0.3, light colors to infectivity 0.15. **A** Fraction of infected agents with respect to remaining susceptible agents, **B** fraction of deceased agents with respect to remaining susceptible agents, **C** fraction of simulations (total N=100) with significant infection wave (>80 subsequent infections), and **D** maximal ICU demand per 100000 individuals, black dotted line represents capacity of ICU beds in Germany. **A,C** Herd immunity threshold calculated from the estimated $R_0$ of 2.71 and 4.87 for infectivities 0.15 (arrow left) and 0.30 (arrow right) are clearly consistent with the random vaccination strategy. Simulations are based on the German municipality Gangelt with 10.351 agents.

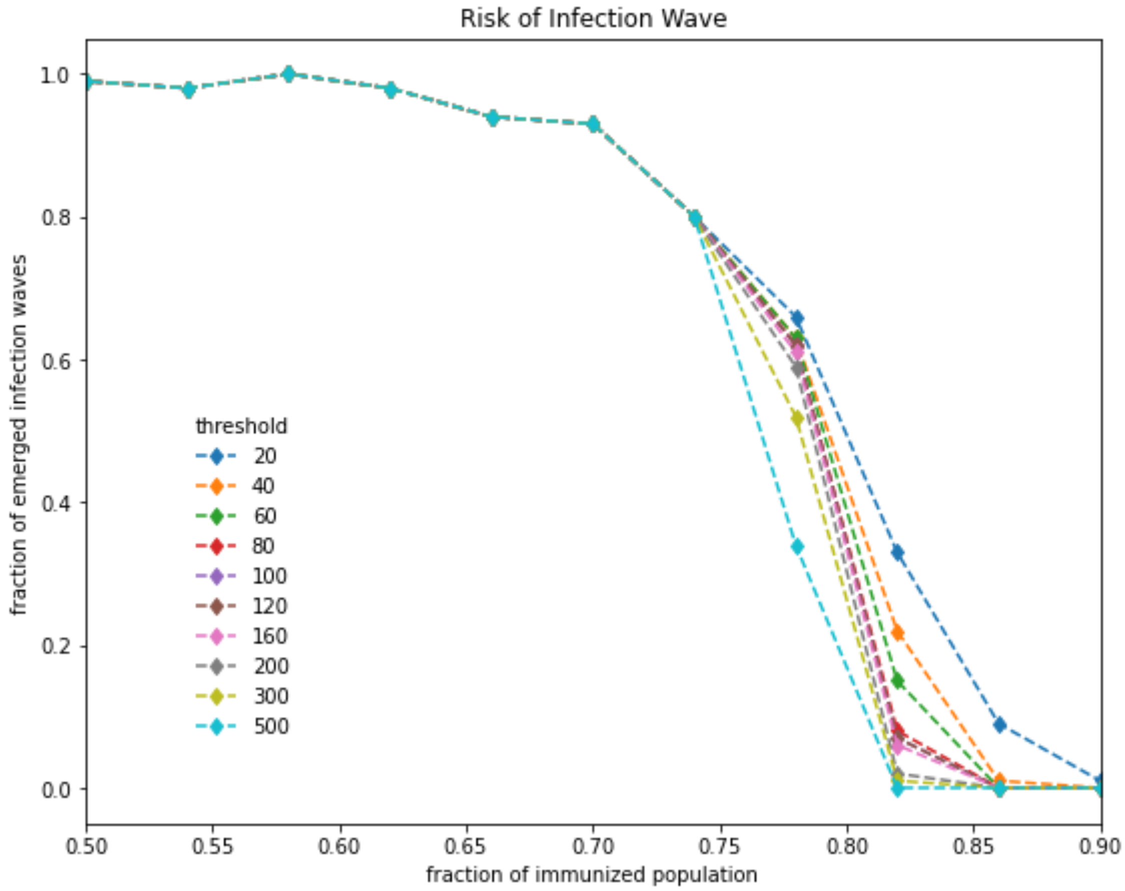


**Figure S27: Effect of threshold size on herd effect analysis (vaccination strategy: random).** Total number of infected agents after 3000 hours was counted. Outbreaks with more infected agents than the given threshold were considered as an emerged infection wave.

#### 4.7 Comparison to Homogeneous Mixing Approach

Classically, SIR modeling uses a homogeneous mixing (HM) approach, i.e. the modeled population is not distinguished by age, locations, or any other attribute that would entitle them to show distinguishable behavior. Our geospatial, demography-based agent-based approach reflects heterogeneity in the behavior of individuals, which is essentially different from the homogeneous mixing approach. This is also reflected in the outcome of various scenarios. The baseline scenario in Gerda shows greater variety in dynamics than the HM (**Fig. S28**).

Homogeneous Mixing GERDA

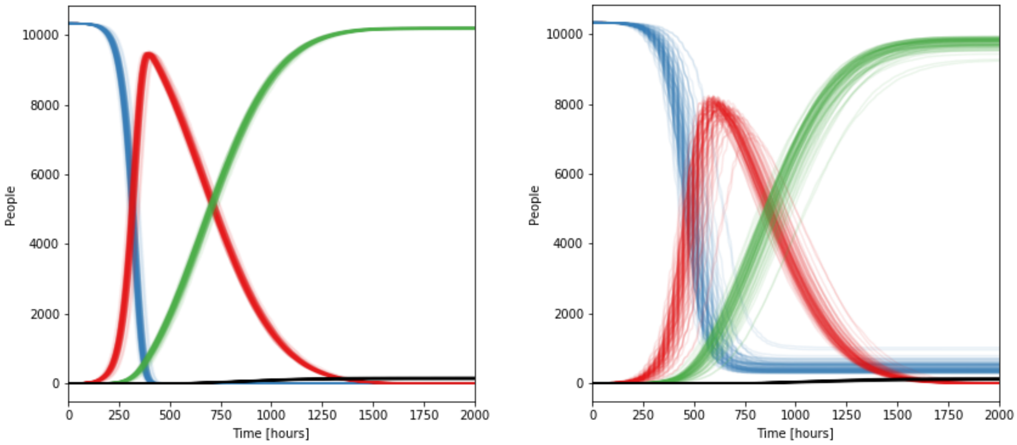


**Figure S28: Health states homogeneous mixing baseline scenario (left) and in the original Gerda baseline scenario (right) for Gangelt.** Resulting time courses for agent health states S (blue), I (red), R (green), and D (black) from 100 simulations for the homogeneous mixing baseline scenario for Gangelt.

Interactions between individuals as recorded in the HHIN as well as infections (iHHIN) also differ significantly between the two approaches. Figure **S29** shows the total and unique number of interactions as well as daily interactions between age groups and daily infections as heat maps.

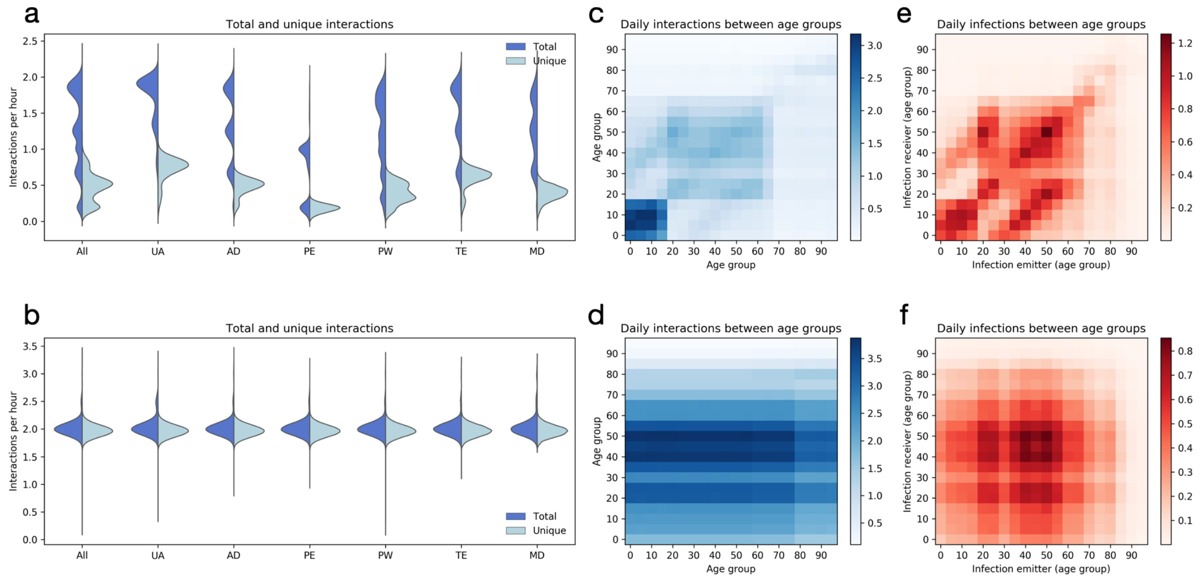


**Figure S29: Comparison of interaction- and infection patterns between homogeneous heterogeneous simulations. (a)** Distributions of average total and unique interactions among individuals of different occupations, in the heterogeneous model. **(b)** See description of (a), results obtained for the homogeneous mixing model where the age-distribution is still contained. **(c)** Mean daily unique interactions between different age-cohorts for the heterogeneous model. **(d)** See description of (c), results obtained for the homogeneous model. **(e)** Mean daily infection-transmissions between different age-cohorts for the heterogeneous model. **(f)** See description of (e), results obtained for the homogeneous model.

Also, the over- or underrepresentation of occupation groups looks different between homogeneous and heterogeneous approaches (**Fig S30**). Homogeneous approaches don't show over- or underrepresentation (number not exactly zero result from stochastic noise).

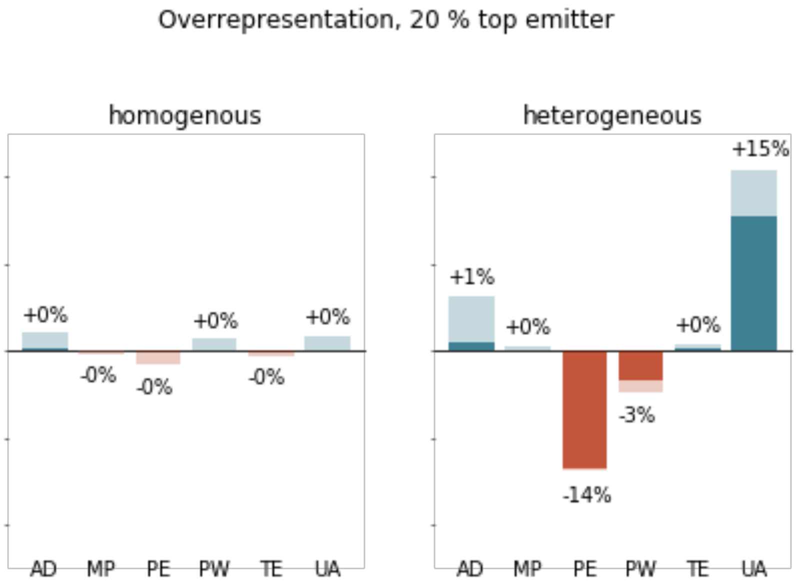


**Figure S30: Comparison between homogenous and heterogenous mixing with respect to the over and under representation of agents with specific schedule types in the fractions of top spreaders.** Difference in the fraction of spreaders with specific schedule type to the fraction of this type in the population.

#### 4.8 Exemplary application to other communities

While all major outcomes of the model approach have been presented for the municipality Gangelt, we have applied the method to other European towns, namely Heinsberg, Simbach, Linsengericht, and Oranienbaum in Germany, Vaxholm in Sweden, Epping in UK and Zhikron Ya’akov in Israel. We see differences in the precise response to infections or to NPIs which can be explained by the size, the demography and the composition of locations of each town. **Figure S31** shows exemplarily the effect of lockdown and reopening for different towns. **Figure S32** presents the analysis of the so-called November lockdown in Germany depending on the incidence value at which different communities started the lockdown.

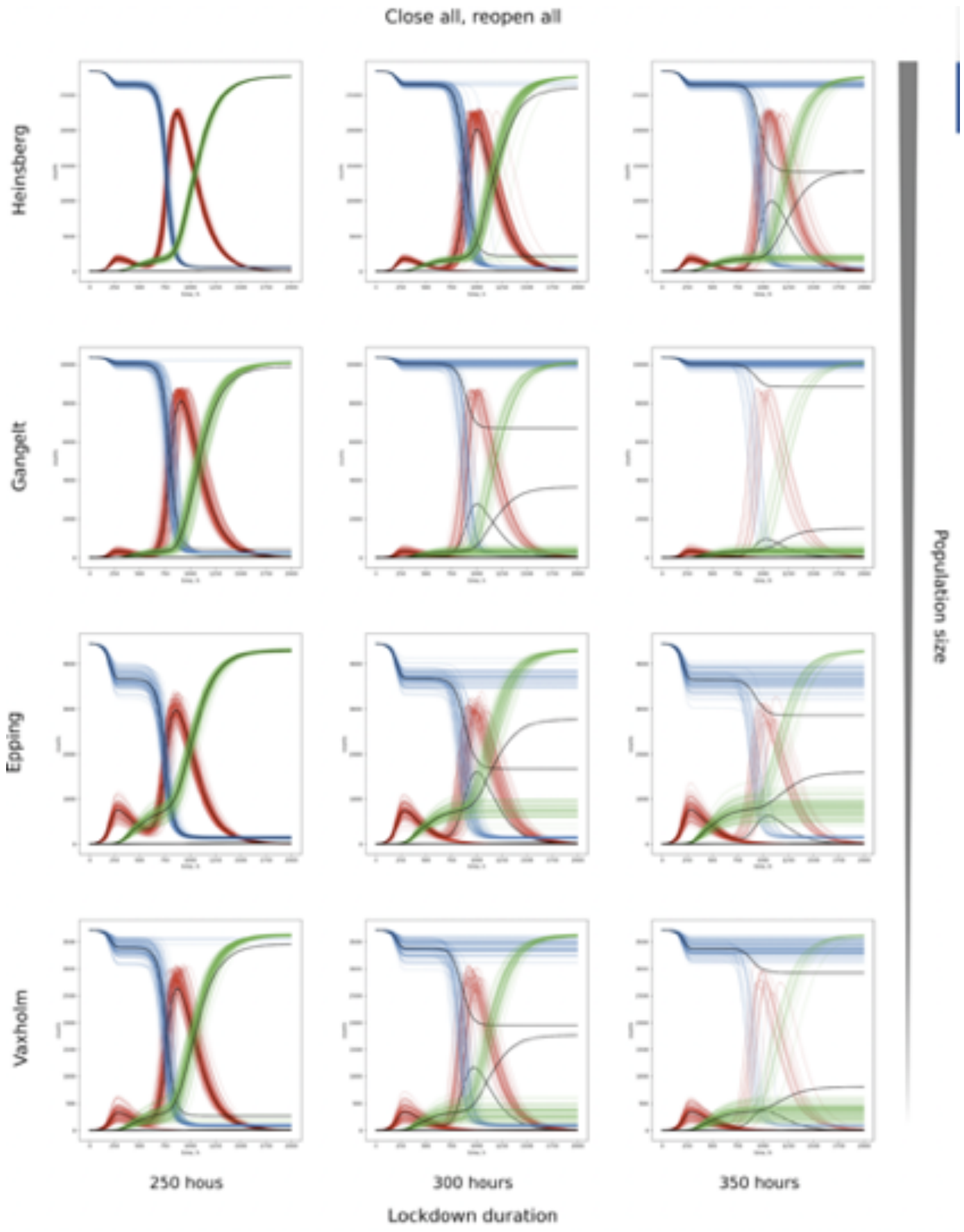


**Figure S31:** Effect of lockdown duration in different European towns. The municipalities Heinsberg (DE, ca 40.700 inhabitants), Gangelt (DE), Epping (UK, ca 11.000 inhabitants), and Vaxholm (SE, ca 4.850 inhabitants) have been considered. Lockdown starts at 200h. All parameter values are like in the baseline scenario for Gangelt.

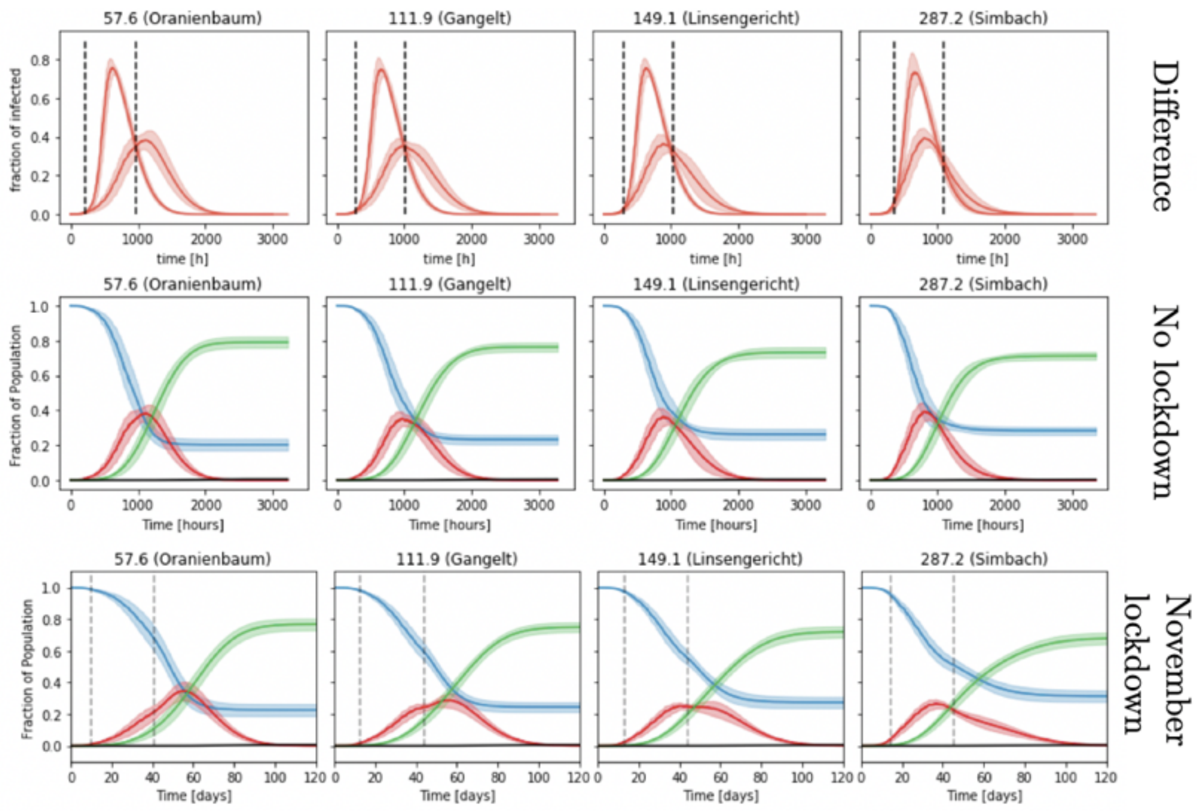


**Figure S32:** Effect of the November (2020) lockdown in different German towns. The lockdown has been simulated for about four weeks. Importantly, the towns started with different incidence values.

#### 4.9 The Movie

Movie S1 [https://www.tbp-klipp.science/GERDA-model/Movie-1.mp4] visualizes the temporal and spatial dynamics of infection spreading resulting from the simulation of the baseline scenario with a still image in **Fig. 1B**. Note that the daily rhythm shown in the movie results from individuals moving between their respective homes and workplaces. The infection starts with two infected individuals in one household at time 0 h. At times 100 h and 200 h more and more infected agents are observed, especially at the geospatial hubs (e.g., center of the town).
